# A MULTI-DIMENSIONAL MACHINE LEARNING APPROACH FOR CARDIOVASCULAR DISEASE PREDICTION IN THE UK BIOBANK STUDY

**DOI:** 10.1101/2025.11.14.25340257

**Authors:** Juan José Hernández Morante, Elena Murcia García, Carlos Martínez-Cortés, Carmen Piernas, Horacio Pérez-Sánchez

## Abstract

Cardiovascular diseases (CVD) are complex disorders involving the impaired function of blood vessels or the heart. Several risk factors contribute to CVD, including diet, body composition and physical activity. We developed an interpretable machine learning model integrating nongenetic factors from 502,134 UK Biobank participants, of whom 53,378 experienced a CVD event over eighteen years of follow-up. Age, cystatin C, and self-perceived health status emerged as the most relevant predictors of CVD. By combining these variables into a multidimensional model, we achieved excellent predictive performance, with an area under the ROC curve of 0.99 and both sensitivity and specificity above 0.97 in the validation cohort. Although cardiovascular events arise from multiple factors, our results show that data integration and machine learning can accurately predict individual risk using simple measurements. This approach could also support the prediction of other cardiovascular outcomes and aid personalized risk assessment and preventive care.

## Introduction

Cardiovascular diseases (CVD) are major causes of mortality worldwide, including ischemic coronary disease, followed by stroke, and hypertensive coronary disease, according to the latest WHO’s report^1^. Despite the vast amount of information available on factors involved in cardiovascular impairment, we still lack adequate prevention and intervention strategies, as evidenced by the fact that the prevalence and incidence of these pathologies continue increasing significantly^2^. Traditional risk indices, such as the Framingham Risk Score, are commonly used for determining CVD risk^3^. However, these indices do not take into account the large amount of information that is now available for CVD risk prediction, which is a limitation in establishing individualized risk factors for later prevention strategies.

Traditionally, cardiovascular risk involves factors such as age, sex, blood cholesterol, blood pressure, smoking, and history of diabetes^4^. Several other factors, such as nutritional factors (total energy intake^5^, fatty acid composition^6^, consumption of specific food groups^7^) as well as physical activity^8^, or sedentary behaviour^9^ have been strongly related to the incidence of CVD-related outcomes and death. In addition, certain sociodemographic characteristics, such as socioeconomic status^10^ and educational level^11^ are also associated with increased risk of CVD. Current knowledge of cardiovascular diseases has not only been extended to include these traditional risk factors, but there are new assessment tools, including various biochemical markers such as cystatin C^12^, or other physical tests such as the thickness of the intima media of the carotid artery^13^, which can help improve the diagnosis of these diseases. However, these factors are not routinely measured, since it is unknown to what extent the use of these measures improves individual risk assessment.

Given the diversity of factors to be considered and the interconnections among them, the evaluation of risk factors through classical statistical procedures like regression analysis may be inadequate or insufficient^14^. Today, modern machine learning (ML) techniques, especially those that allow individual and global interpretability of the relative influence of each factor, can help develop person-oriented predictions of CVD^15^. Such algorithms, often referred to as model-agnostic ML methods, provide greater flexibility and accuracy than traditional approaches, such as logistic regression. For instance, they allow the use of neural networks and gradient boosting, which frequently outperform conventional models while also enabling the selection of the most appropriate model for each dataset^16^. One of the main challenges when using ML, however, is the complexity of these models. When dealing with large numbers of variables, understanding how predictions are generated can be difficult. To address this issue, interpretability frameworks have been developed that align better with human reasoning, such as SHAP (SHapley Additive exPlanations), which quantifies the contribution of each feature to a given prediction^17^.

However, performing SHAP value analysis can be challenging due to several considerations. In practice, SHAP computations are often resource intensive, particularly for large datasets. The computational time also increases significantly with dataset size, especially in high-dimensional data such as those derived from the UK Biobank cohort, since the number of feature permutations grows exponentially with the number of features^18^.

Despite these limitations, the present study is among the few to apply interpretable ML analysis to a large and complex dataset for CVD prediction, effectively addressing many of the computational challenges inherent in these approaches. Using the UK Biobank cohort, we integrated sociodemographic, dietary, lifestyle, physical measures, and biochemical and metabolomic features to evaluate the prediction ability of these features on cardiovascular event prevention using a multi-dimensional ML approach. We validated the accuracy of the ML model and developed individualized CVD prediction models that enabled a more precise characterization of the key features determining CVD risk at the individual level.

## RESULTS

### Demographic and Clinical Characteristics

The primary cohort selected for the present work included 475,570 participants without evidence of a cardiovascular disease (CVD) event up to 1 year after their baseline assessment. Since information from all the variables was not available for all the participants, the sample sizes for each group of features were as follows: n=502,134 for biochemical markers; n=247,923 for metabolomics; n=126,651 for dietary features; n=446,107 for physical measures; n=213,505 for lifestyle and sociodemographic characteristics. The mean time to a CVD event was 2539 days (range 0 to 5661 days), during which 53378 participants had a CVD event (11.2% of the population): 38,067 cases of coronary heart disease, 10,937 cases of stroke, and 14,729 cases of congestive heart failure. In addition, 8873 participants had a record of two CVD events (0.9%), and 741 individuals (0.001%) had three CVD events. Regarding the baseline clinical characteristics of the study cohort (Table 1), the proportion of women was slightly higher than men (55% vs 45%), and the average body mass index (BMI) characterized the population as overweight. Systolic blood pressure values were slightly above the recommendations, although the diastolic blood pressure was within the values within the normal range. The biochemical parameters related to glucose metabolism were within the normal range, although those of lipid metabolism were borderline high, especially in those individuals that developed a CVD event.

**Table 1.** Baseline characteristics of the study cohort.

| Characteristics | All<br>(n =475570) | No CVD<br>(n =422192) | CVD<br>(n =53378) |
| --- | --- | --- | --- |
| **Demographics** |  |  |  |
| Women (n, %) | 264852 (55.7%) | 242960 (51.1%) | 21892 (4.6%) |
| Age at recruitment (y) | 56 ± 8 (37-73) | 56 ± 8 | 60 ± 7 |
| White ethnicity (n, %) | 447021 (94.1%) | 396791 (83.6%) | 50230 (10.4%) |
| **Socioeconomic status** |  |  |  |
| Townsend index | -1.3 ± 3.1 (-6.26 – 11.00) | -1.4 ± 3.1 | -1.0 ± 3.3 |
| Higher degree (college or university degree) (n, %) | 155641 (32.7%) | 142415 (29.9%) | 13226 (2.8%) |
| **Behavioral factors** |  |  |  |
| Current smoker (n, %) | 49545 (10.4%) | 42077 (8.8%) | 7468 (1.5%) |
| Current alcohol use (n, %) | 437037 (92.1%) | 389177 (82.0%) | 47860 (10.1%) |
| Physical activity (MET-min/week) | 2654 ± 2652 (0 – 19278) | 2648 ± 2637 | 2702 ± 2776 |
| **Physical measurements** |  |  |  |
| Weight (Kg) | 77.7 ± 15.8 (30.0 – 197.7) | 77.1 ± 15.6 | 82.3 ± 16.9 |
| Body mass index | 27.3 ± 4.8 (12.1 – 74.7) | 27.2 ± 4.7 | 28.7 ± 5.2 |
| Body fat (%) | 31.4 ± 8.6 (5.0 – 69.8) | 31.4 ± 8.6 | 31.6 ± 8.6 |
| Fat-free mass (kg) | 53.0 ± 11.5 (23.4-100.0) | 52.6 ± 11.4 | 56.0 ± 11.7 |
| Body water (kg) | 38.8 ± 8.4 (18.9-84.3) | 38.5 ± 8.3 | 41.0 ± 8.6 |
| SBP (mmHg) | 140 ± 20 (62 – 268) | 139 ± 19 | 146 ± 20 |
| DBP(mmHg) | 82 ± 11 (32 – 148) | 82 ± 11 | 84 ± 11 |
| **Biochemical parameters** |  |  |  |
| Fasting Glucose (mmol/L) | 5.1 ± 1.2 (1.0 – 36.8) | 5.1 ± 1.1 | 5.4 ± 1.8 |
| HbA1c (mmol/mol) | 36.1 ± 6.8 (15.0 – 515.2) | 35.6 ± 6.1 | 38.7 ± 9.3 |
| Cholesterol (mmol/L) | 5.7 ± 1.2 (0.6 – 15.5) | 5.8 ± 1.1 | 5.4 ± 1.3 |
| Triglycerides (mmol/L) | 1.8 ± 1.0 (0.2 – 11.3) | 1.71 ± 1.01 | 2.0 ± 1.1 |
| cHDL (mmol/L) | 1.5 ± 0.4 (0.2 – 4.4) | 1.47 ± 0.38 | 1.3 ± 0.4 |
| cLDL (mmol/L) | 3.6 ± 0.9 (0.3 – 9.8) | 3.6 ± 0.9 | 3.3 ± 1.0 |
| **Clinical History** |  |  |  |
| Treatment/Medications taken (n) | 2.3 ± 2.5 (0 – 36) | 2.1 ± 2.3 | 3.4 ± 3.1 |
| Diabetes (n, %) | 21672 (4.6%) | 16286 (3.4%) | 5386 (1.1%) |
| Cancer (n, %) | 36052 (7.6%) | 30893 (6.5%) | 5159 (1.1%) |
| Other serious medical conditions (n, %) | 89334 (18.8%) | 74710 (15.7%) | 14624 (3.18%) |
| Overall health rating (Excellent or Good) (n, %) | 359608 (75.8%) | 326305 (68.7%) | 33303 (7.1%) |

Values are mean, standard deviation and range (min-max) for quantitative variables and frequency and percentage for qualitative variables. DBP: diastolic blood pressure HDL: High-density lipoprotein. LDL: Low-density lipoprotein. SBP: systolic blood pressure.

### Lifestyle and sociodemographic characteristics *in association with CVD events*

We investigated sociodemographic characteristics, such as ethnicity, as well as lifestyle variables such as time spent watching TV, physical activity and work-related characteristics. The energy expenditure for all activities was higher in those with CVD, especially in moderate and walking activities. In contrast, energy expended on vigorous activities was higher in those without CVD (Table S9). Within these features, in addition to age, the number of medications (OR: 1.51, CI: 1.47-1.56) was a significant predictor. Self-perception of health was the next best predictor of CVD risk, in the sense that those with a worse perception of health had a higher risk of suffering a CVD event (OR: 1.17, CI: 1.14-1.20). On the contrary, walking pace (OR:0.92, CI: 0.80-0.95) and intake of minerals (OR:0.95, CI: 0.92-0.98) and vitamin supplements (OR:0.95, CI: 0.92-0.97) were associated with lower risk (Table S10 and Figure 5a). Figures 5b–e represent the differences and the frequency of individuals attending to the presence of a CV event or not.

After sex, overall health rating (self-perception of health) was more important than variables such as age, according to the ML model (Figure 5f), together with walking pace. Again, individual SHAP models allowed a better interpretation of the specific influence of the different variables in any participant (Figures 5g–h shows two examples of CVD risk predictions).

### Dietary features in association with CVD events

Dietary features were evaluated using the food group classification described previously^19^. Participants with a CVD event at follow up were characterized by higher total daily energy intake, beer drinking, and white bread intake, and by lower intakes of white rice/pasta, water and olive oil (Table S5); however, it is important to highlight the large variability within each group. The LR procedure identified several foods that were associated with a higher CVD risk (Table S6 and Figure 3a). Those with a higher OR were plant-based spread, both lower fat (OR: 1.03, ci: 1.02-1.03) and regular fat (OR: 1.02, CI: 1.01-1.02). In contrast, nut-based spreads (OR: 0.98, CI: 0.97-0.99) and vegetable dips (OR: 0.99, CI: 0.98-0.99) were associated with lower OR. Figures 3b–e represent the mean values of these variables depending on the presence or absence of a CVD throughout the follow-up period.

In this dataset, ML data were quite similar to the RL analysis (Figure 3f). The most influential variables were white bread, white pasta & rice, and plant-based fat spread. Also to note, the ML model revealed a U-shape relationship between the intake of these foods and CVD risk, since a lower and higher intake of these dietary-related variables was associated with a higher probability of CVD. The individual SHAP values (Figure 3g–h) highlighted the importance of an individual evaluation of the influence of dietary characteristics. For instance, the individual in figure 3g showed an increased risk by white bread, but in the individual in figure 3h, white bread intake was identified with a low risk of CVD.

### Physical measures in association with CVD events

The association between anthropometric, echocardiographic, and other physical measures and the risk of CVD were evaluated. Because these differences highly depend on sex and age, these features were included in the analyses. There were higher waist circumference and BMI in the CVD group, although both groups (CVD and no CVD) showed a mean BMI indicative of overweight (Table S7). The carotid intima-medial thickness (cIMT) was also higher in those participants with CVD, while spirometry variables indicated better pulmonary function in those without CVD events. The LR analysis showed that only a few physical variables were able to predict CVD events (Table S8). Interestingly, although cIMT (OR: 3.64, CI: 3.60-3.69) was the most relevant predictor, age (OR: 1.52, CI: 1.50-1.54) and sex (OR: 1.52, CI: 1.48-1.56) were also highly relevant. The most relevant anthropometric parameter associated with a higher CVD risk was waist circumference (OR: 1.44, CI: 1.41-1.48) (Figure 4).

The ML model also identified waist circumference as the variable with the greatest impact on the prediction (Figure 4f), followed by birth weight as the second variable in relevance. Other variables like BMI, sex, or body fat were less relevant than other functional parameters, such as arterial stiffness, respiratory capacity, or blood pressure. Several variables (birth weight, pulse rate, etc.) showed a dimorphic relationship with CVD, with both lower and higher values of these features indicative of high CVD risk. In this context, individual SHAP analysis allowed for a better individual identification of CVD risk. For instance, in the example person of Figure 4g, birth weight accounted for almost all the CVD risk, in comparison to the individual of Figure 4h, where this feature showed only a modest association.

### Biomarker features in association with CVD events

Attending to the effect size (Table S1), higher differences between those who suffered CVD and those who did not were observed regarding Cystatin C, urate, glycated haemoglobin, cHDL, and cholesterol. All the biochemical parameters showed statistically significant differences between participants with vs without a CVD event (Table S1). For the selection of biomarker variables in the logistic regression model, the stepwise selection method based on the Akaike Information Criterion (stepAIC) was used, followed by a multicollinearity analysis using the VIF test (Table S2). The top features individually associated with an increase in CVD risk were Cystatin C (OR: 1.45, CI: 1.23-1.70), glycated haemoglobin (OR: 1.36, CI: 1.18-1.56), and testosterone (OR:1.28, CI: 1.13-1.44), while IGF1 (OR:0.77, CI: 0.67-0.88), Apolipoprotein A (OR:0.80, CI: 0.69-0.92), and oestradiol (OR:0.83, CI: 0.65-1.02), were associated with a decrease risk (Figure 1a). However, in the oestradiol case, the statistical significance disappeared after adjustment for multiple variables (FDR = 0.2078). Figure 1b–e represents the mean differences between those participants who developed a cardiovascular event or not in main predictive variables.

**Figure 1.**
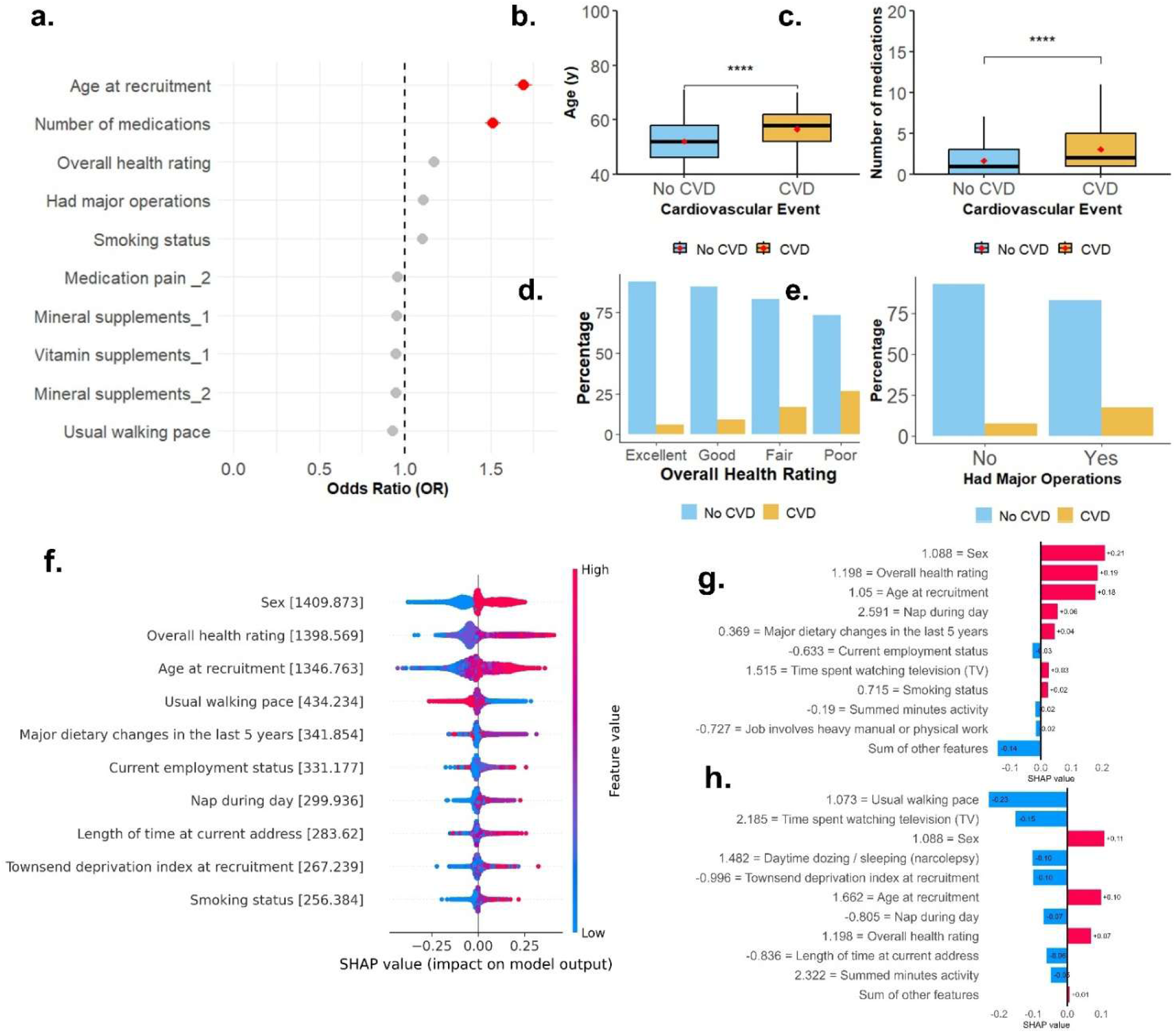
Lifestyle features monotonically associated with a cardiovascular event. **a**, Associations between biomarker parameters and CVD. The measure of the centre corresponds to the odd ratio, and the error bars represent the 95% confidence intervals; the vertical dashed line corresponds to an odd ratio of 1. Red circles highlight features with OR > 1.5; **b-c,** Box plots showing the differences between age and the number of medications attending to the presence or not of a CVD event; d-e, Bar plots showing the differences between the overall health perception and the presence of major operations attending to the presence or not of a CVD event; **f,** predictive machine learning model derived from the SHAP-values considering only lifestyle features; **g-h**, examples of a participant who suffered a cardiovascular event and one who did not.

The predictive machine learning (ML) (Figure 1f) revealed that, globally, testosterone, platelet volume, and haemoglobin concentration were the highest contributors to the ML model. Concentrations of potassium and sodium in urine were also relevant to the model. In contrast, cholesterol-related parameters seemed to have little influence on the global prediction model. Nevertheless, the relevance of the present work was the individual prediction of CVD risk. Attending to SHAP values, Figure 1g represents a participant who suffered a cardiovascular event, and Figure 1h represents a participant who did not. The influence of each variable varied from one person to another, thus justifying the need for individual risk assessment.

### Metabolomic features in association with CVD events

Almost all the metabolomic parameters were statistically and significantly different depending on the presence or absence of a CVD event (Table S3). Higher differences between groups were observed regarding cholesterol, triglycerides, or phospholipids to total lipid content of lipoproteins. Fatty acid composition revealed higher MUFA and SFA in those with a CVD event during follow-up; in contrast, PUFA were higher in those without a CVD event, mainly because of higher omega-6 content. Sphingomyelins were also higher in individuals without CVD events.

The metabolomic features individually associated with CVD events attending to the LR (Table S4 and Figure 2a) were creatinine (OR: 1.29, CI: 1.28-1.31) and the percentage of cholesterol-to-total lipids in medium LDL (OR: 0.69, CI: 0.67-0.70), considering the factor associated with greater or lesser risk, respectively. The model also revealed the importance of several amino acids, such as glutamine (OR: 1.19, CI: 1.18-1.21), tyrosine (OR: 1.15, CI: 1.13-1.17), glycine (OR: 0.83, CI: 0.82-0.84), and isoleucine (OR: 0.85, CI: 0.84-0.87). As with biomarkers, glycoprotein acetyls were also associated with a higher CVD risk.

**Figure 2.**
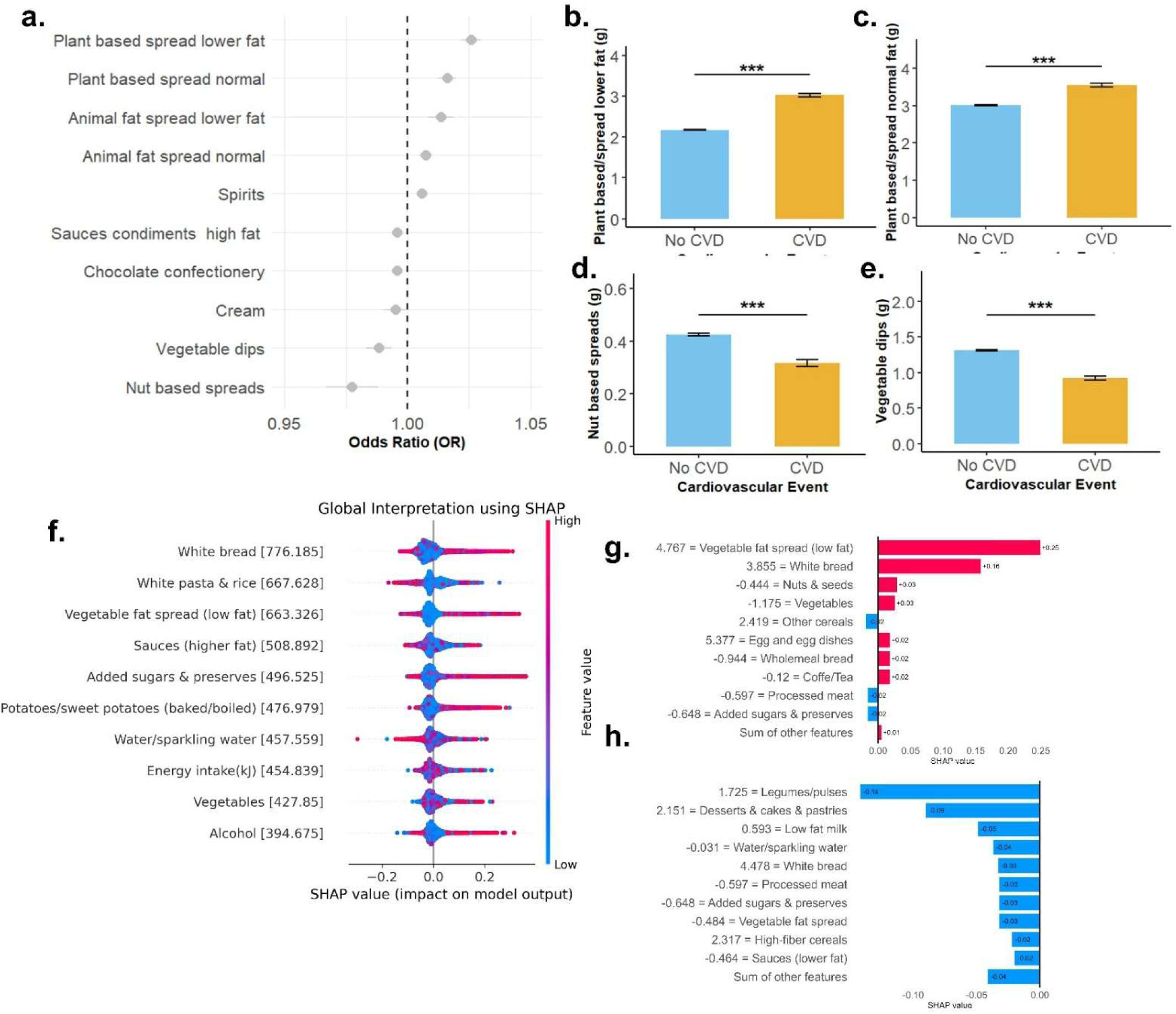
Dietary features associated with a cardiovascular event. **a**, Logistic-regression derived associations between food intake and CVD. Only top variables with higher or lower OR are shown. The measure of the centre corresponds to the odd ratio, and the error bars represent the 95% confidence intervals; the vertical dashed line corresponds to an odd ratio of 1. **b-e**, Bar plot showing the differences between plant-based spread lower fat attending to the presence or not of a CVD event. **f,** predictive machine learning model derived from the SHAP values; **g-h**, examples of a participant who suffered a cardiovascular event and one who did not.

**Figure 3.**
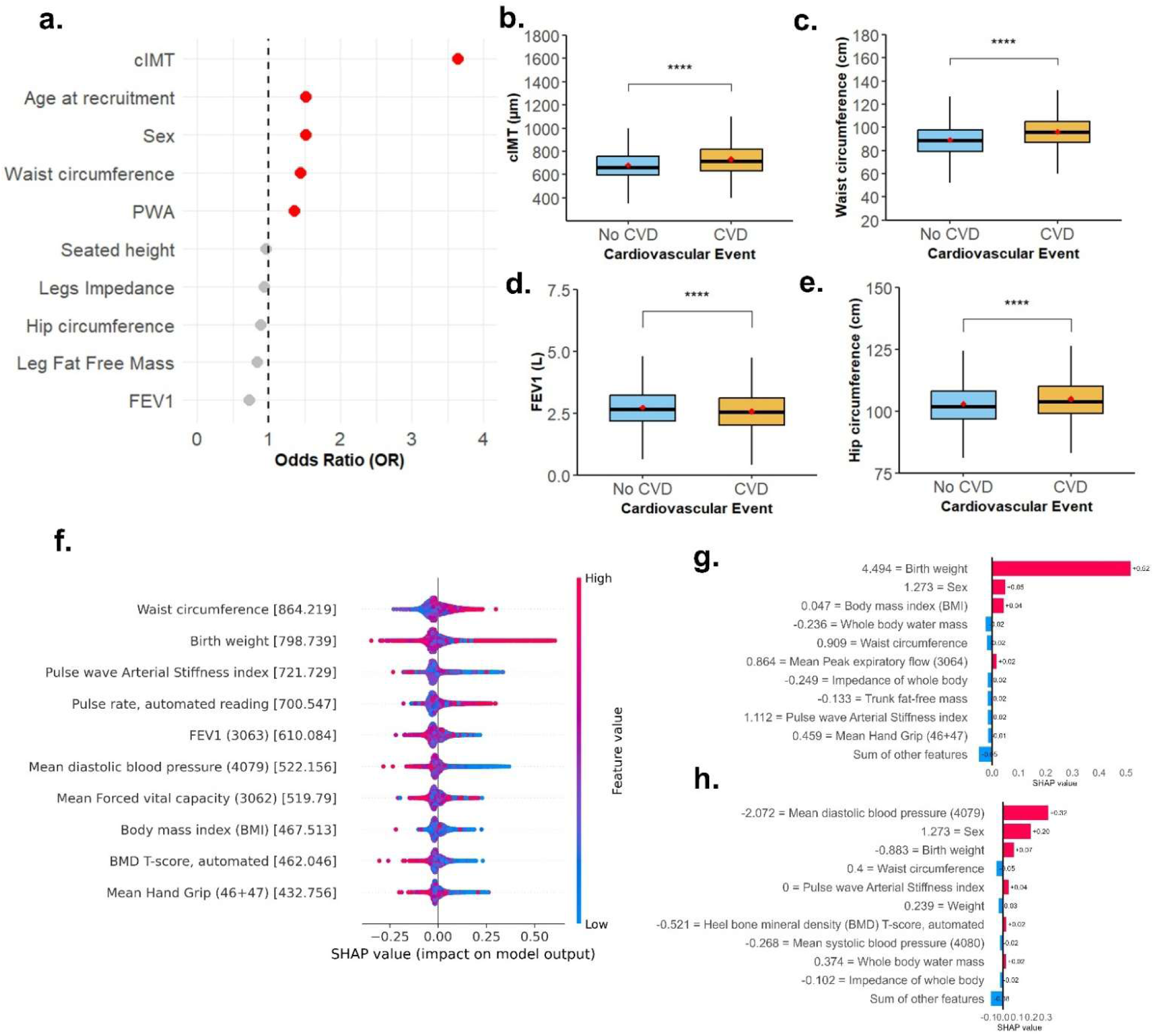
Physical measures associated with a cardiovascular event. a. The measure of the centre corresponds to the odd ratio, and the error bars represent the 95% confidence intervals; the vertical dashed line corresponds to an odd ratio of 1. b-e. Box plots showing the differences between several of the better predictors of the presence or not of a CVD event. **f,** predictive machine learning model derived physical measures according to the SHAP values; **g-h**, examples of a participant who suffered a cardiovascular event and one who did not.

**Figure 4.**
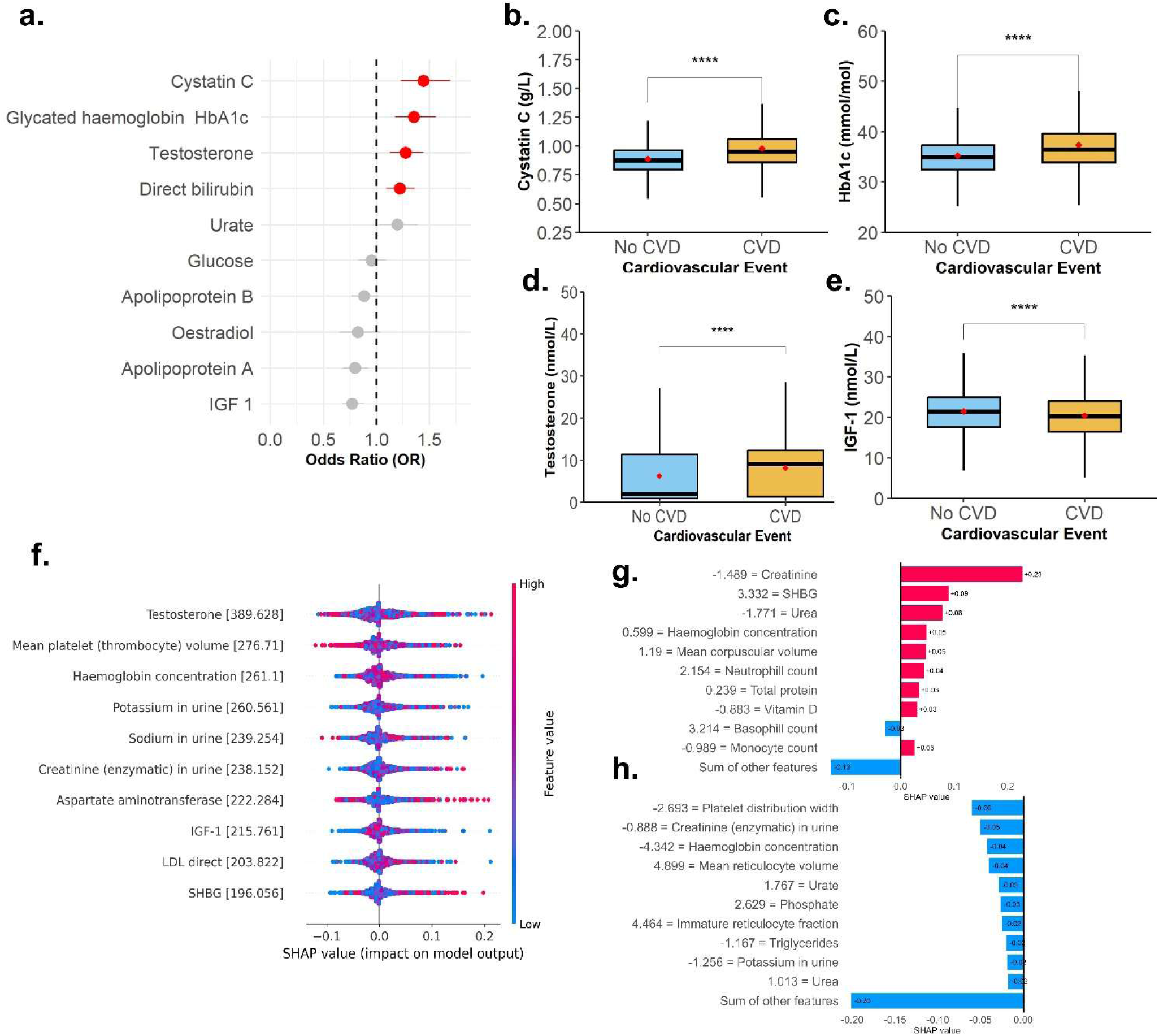
Biomarker features associated with a cardiovascular event; **a**, Top five and bottom five features associated with CVD risk. Only statistically significant associations were shown. The measure of the centre corresponds to the odd ratio, and the error bars represent the 95% confidence intervals; the vertical dashed line corresponds to an odd ratio of 1; **b-e,** Box plots showing the differences between most significant biomarker parameters attending to the presence or not of a CVD event; **f,** predictive machine learning model derived from the SHAP values; **g-h**, examples of a participant who suffered a cardiovascular event and one who did not.

The prediction of CVD with metabolomic parameters through ML yielded good performance, with an AUC with the RF algorithm of 0.998. In this model, the most influential features were creatinine and concentration of IDL particles (Figure 2f). The ML model indicated that higher values of creatinine were associated with a higher impact on the feature value, and therefore, a higher probability of CVD. The opposite was true for total cholesterol, the concentration of IDL particles, and linoleic acid (Figure 2f). The individual ML prediction (Figures 2g and 2h) showed a mixture of positive and negative features that allowed better individual prediction.

### Dataset integration for improved CVD prediction

The first step of our approach consisted of describing individual associations of the different features within biomarkers, metabolomics, diet, lifestyle and physical measures with CVD events. Although there was high accuracy and reliability of the models within each dataset, we wanted to increase our prediction ability by grouping the most relevant variables of every dataset into a single analysis by using a framework that integrated features into a new ML predictive model of CVD events. Figure 6a shows the integrative ML model, which allowed the detection of CVD with 99.7% accuracy, and allowed the identification of the specific weight of all dimensions in the prediction of CVD. Age was the most influential feature, followed by cystatin C, overall health rating, testosterone, and sex. Classic CVD factors, such as BMI or cholesterol-related parameters, were not present in the integrative ML model. Similarly, diet features were absent in this model (Figure 6a). The individual analysis of the risk of CVD reported very extreme differences depending on the individual. For instance, overall health ratings can be both promoting or protecting from a CVD event (Figures 6b and 6e), as is the case with age, which was associated with both higher and lower risk of a cardiovascular event (Figures 6c and 6d).

**Figure 5.**
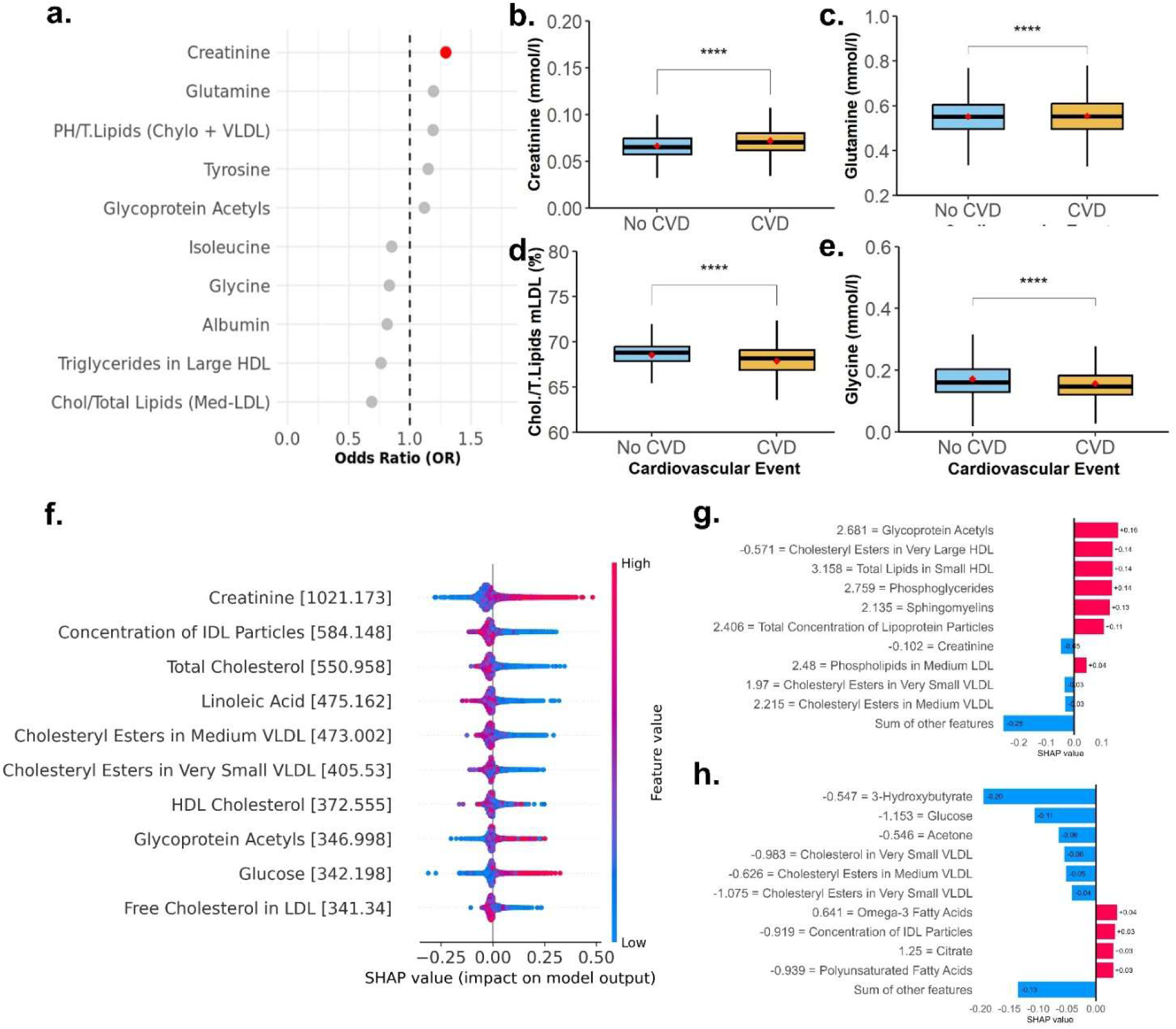
Metabolomic features associated with a cardiovascular event. **a**. Top associations between biomarker parameters and CVD. Only statistically significant associations were shown. The measure of the centre corresponds to the odd ratio, and the error bars represent the 95% confidence intervals; the vertical dashed line corresponds to an odd ratio of 1. **B–e,** Box plots showing the differences between several of the better predictors of the presence or not of a CVD event. **f,** predictive machine learning model derived from the SHAP values considering only metabolomic features; **g-h**, examples of a participant who suffered a cardiovascular event and one who did not.

**Figure 6.**
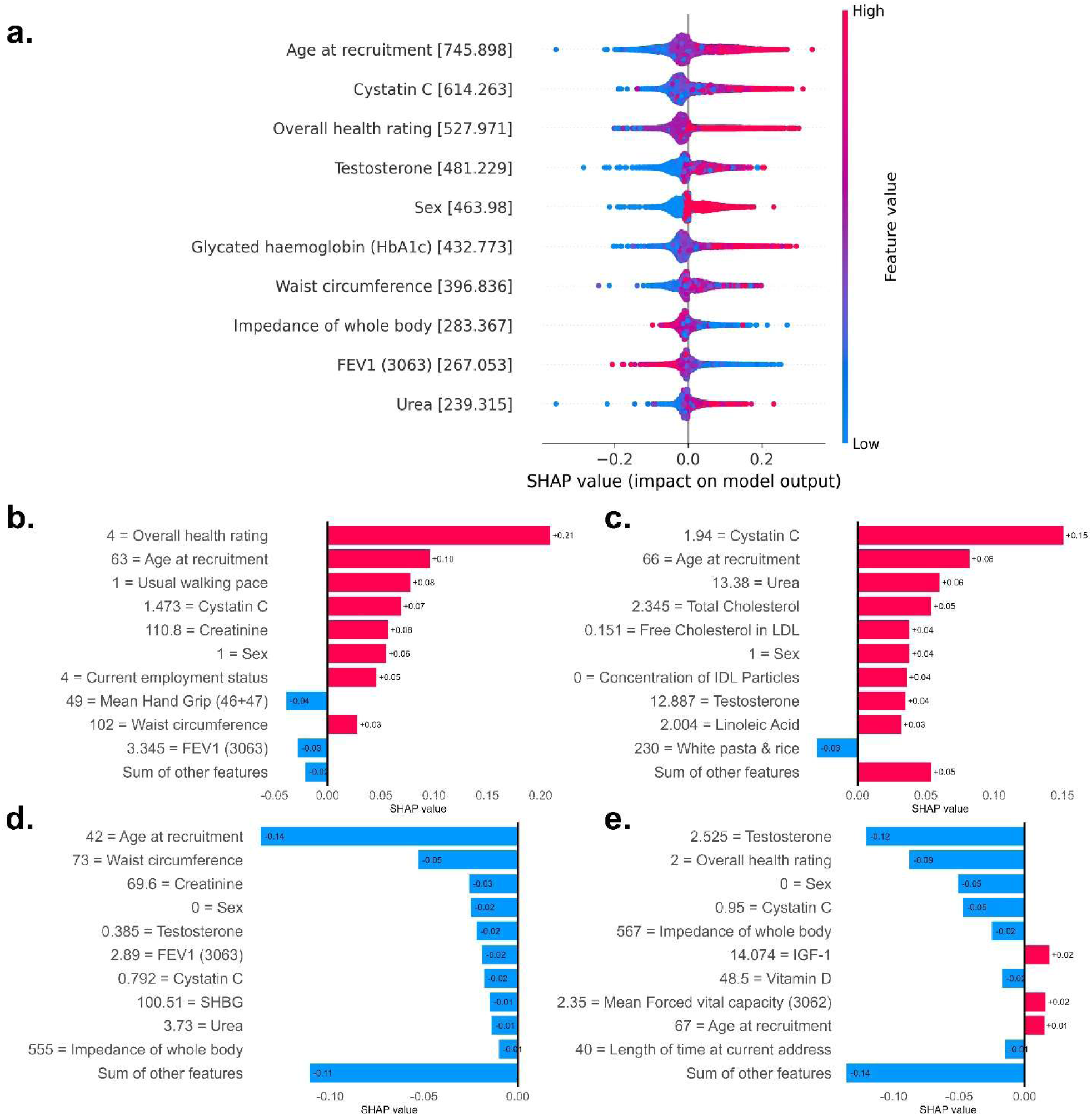
Global framework integrating biomarker, metabolomic, dietary, physical, and lifestyle features into a new ML predictive model of CVD events; a, global predictive machine learning model derived from the SHAP values including all features; **b-c**, examples of two participants who suffered a cardiovascular event; and **d-e**, examples of SHAP models for two participants who did not suffer a cardiovascular event.

## DISCUSSION

In the present study, we developed an agnostic machine learning methodology based on a multi-dimensional approach to adequately predict a cardiovascular event in the UK Biobank cohort. Through this approach, we have highlighted the role of well-known CVD-related factors, such as age, sex and cystatin, in CVD prediction. In addition, the predictive models developed in this large cohort revealed the influence of social determinants, such as the self-perceived overall health rating.

Several implications arise from these results. First, agnostic models were able to predict the presence of CVD with high accuracy, precision, and reliability, outperforming multiple logistic regression models (despite any model obtained in the different LR approaches). This is the first work in which a methodology of agnostic machine learning models has been used, through the SHAP methodology, in such a large population and with a diverse number of descriptors. The SHAP approach provided a clear interpretation of how each feature influenced the model’s predictions, which is crucial for patients in whom cardiovascular events can be fatal. By offering clear explanations, SHAP increases the confidence of healthcare professionals in the decisions made by the model, which is especially important in health ^20^. Nevertheless, this research is not the first using UK biobank-derived data with this methodology. Previous research have tried to predict different health-related issues, such as suicidal ideation ^21^, depression ^22^, liver triglyceride content, and inflammation ^23^, among others. Previous research by Atehortúa et al. predicted CVD events through exosome variables determining SHAP values ^24^, although these previous studies involved fewer individuals and variables than the present one. Nevertheless, they also described several associations that were also found to be relevant in our models, specifically in relation to current employment status^24^. Notably, previous studies also agreed with our findings regarding lifestyle-related factors, such as daytime naps; however, the relevance of these variables decreased when other predictors were included in the global model.

The present study confirmed the association of several features with cardiovascular disease, but also allowed us to describe the implications of several factors not well described. For example, cystatin c was revealed as one of the most relevant biomarkers to predict CVD, both within the biomarker dataset and in the global dataset. As an alternative to estimate glomerular filtration rate, cystatin C level associates more closely with future risk of cardiovascular disease (CVD) and mortality than eGFR, as described by Lees et al ^25^. One variable with less evidence is the platelet volume, which was associated with an increase in CVD risk according to the ML algorithms, so further studies specifically based on determining the role of the platelet volume on CVD may confirm this observation. On the other hand, the ML model already described testosterone as the most important determinant of CVD risk. There is some controversy regarding the role of testosterone in CVD risk. The relationship between testosterone and cardiovascular risk is multifaceted and has been thoroughly studied, with previous research showing both protective and detrimental effects depending on testosterone levels. Multiple large-scale studies have consistently shown that low endogenous testosterone levels are associated with increased cardiovascular mortality and morbidity ^26^. The EPIC study found an inverse relationship between endogenous testosterone concentrations and all-cause mortality and cardiovascular disease ^27^. In addition, a meta-analysis has confirmed that men with low testosterone concentrations had a significantly higher risk for all-cause mortality ^28^. However, although replacement clinical trials have shown a similar risk with placebo for major cardiovascular event ^29^. Overall, the role of testosterone in cardiovascular risk may follow a complex pattern. For instance, individuals with low testosterone levels may have an increased cardiovascular risk, but at the same time, people with supraphysiological testosterone concentrations may also show a higher CVD risk, a situation that, at the same time, may depend on other variables like haematocrit levels and kidney function ^29^.

Although blood biomarkers are well-defined variables to predict CVD, one of the advantages of the UK biobank data came from the NMR metabolomic data obtained from the same participants, capturing > 200 biomarker measures, such as lipids and amino acids ineach blood sample. The detailed metabolic profiling provides an enhanced understanding of the molecular mechanisms leading to the onset and progression of chronic diseases and may hereby identify causal biomarkers and targets for drug treatment. In this work, all metabolomic parameters were different according to CVD status. In addition, several factors, such as creatinine and glutamine, were associated with higher CVD risk. Interestingly, blood levels of several amino acids, glutamine, tyrosine, and glycine were associated with CVD risk attending to the LR. The ML models, similarly, identified creatinine as the factor with a higher impact on the feature value, and therefore, a higher probability of CVD. As with cystatin, impaired kidney function was indicative of cardiovascular risk. A growing appreciation of the pathophysiological interrelatedness of metabolic risk factors, such as obesity and diabetes, chronic kidney disease, and cardiovascular disease, has led to the conceptualization of cardiovascular-kidney-metabolic syndrome ^30,31^. The present results reinforce this association, since cystatin remained in the final model as an important feature of CVD risk. Moreover, the comprehensive evaluation of lipoproteins allowed the identification of several lipoprotein particles as protective factors. There is very little information in this regard, although a recent study revealed a positive relation between IDL particles and the 5-year progression of atherosclerosis ^32^, the most relevant parameter, after creatinine, attending to the ML model.

Analyses of dietary characteristics showed the most controversial results. The high heterogeneity between the dietary patterns of the participants may limit the power of the statistical analysis, but also dietary consumption was analysed in grams per day, so although the odds ratios may seem small, the net effect of each dietary variable may be greater than what the analysis represented. The LR analysis identified plant-based spreads as the factors with higher odds of CVD, while vegetable dips and nut-based spreads showed lower odds of CVD. The benefits of nut intake are in line with the observations of the PREDIMED study, a longitudinal study that re-evaluated the benefits of a Mediterranean diet on cardiovascular disease prevention ^33^. However, the association between dairy fat and vegetable fat is controversial. We have recently shown that a breakfast based on vegetable fat was able to reduce detrimental lipoproteins and some inflammation markers, although the effect was lower than with olive oil ^34^. The ML procedure confirmed the influence of vegetable fat on CVD but also highlighted the importance of carbohydrate-rich foods, like pasta, rice, bread, and potatoes, all of them being the most important features of the ML model. Our results described a U-shaped relationship between foods and CVD risk, since both high and low intakes of these foods increased the risk of CVD, while a moderate (median) intake was associated with a lower risk. This U-shaped relationship between fat intake and cardiovascular risk was described decades ago to hypothesize the paradoxical observations of fat intake and cardiovascular incidence in several countries ^35^. This U-shape association could be an adaptive response to excess intake or to food deprivation, which may also be involved in a constitutional variation in energy handling. More recently, it has been proposed that this U-shaped relationship could occur with other classic cardiovascular risk factors in older people ^36^. People enrolled in the UK biobank cohort were older than 40 years; consequently, further replication in younger cohorts are needed to confirm these observations.

Another classical factor that predicted CVD risk was waist circumference, a consistent observation in both LR and ML models. Of note, birth weight was the second feature of interest in the ML model—that is, both low and high birth weight—were indicative of CVD risk. Low birth weight, as an indicative of intrauterine growth retardation, has been associated with a higher risk of hypertension, type 2 diabetes mellitus (T2D), and cardiovascular disease, according to Barker’s hypothesis ^37^. Regarding the other physical measures, respiratory capacity was also an important determinant of the presence of CVD, even to a greater extent than other classical parameters like intima thickness or body fat. A 10-year prospective study conducted on an Italian cohort also showed that pulmonary function decline is associated with increased cardiovascular risk ^38^. Reduced lung function has been reportedly associated with obesity, metabolic syndrome, diabetes, and insulin resistance; conditions which are linked to increased cardiovascular risk ^39^. Therefore, in order to predict a cardiovascular event, it could be of great interest to consider the determination of these parameters, which can be easily determined with, for example, a spirometry test.

In the context of the present work, it is crucial to accentuate the importance of lifestyle factors, given their profound impact on cardiovascular outcomes. Lifestyle factors yielded better accuracy in the RL models. Concretely, we confirmed the association between age, sex, and smoking habits with a higher risk of CVD. Moreover, the self-perceived overall health rating played a crucial role in the accuracy of the prediction model. In the integrated ML model, the self-perception of health was the variable with the highest contribution only after sex. The relevance of this finding cannot be overstated, as it highlights the influence of health perception on CVD outcomes. Several hypotheses might explain this association. On the one hand, a decrease in self-perceived health has been related to inadequate health habits and lower protective behaviours ^40^. Therefore, low self-perceived health could be indicative of suboptimal lifestyle choices. On the other hand, low self-perception of health could be indicative of depressive symptoms, which are, at the same time, indicative of cardiovascular disease, as has been elucidated by Harshfield et al. in a previous study conducted in the same cohort. Another result that underscores the importance of self-perception of health is the observation that even in the global ML model, self-perception of health persists as one of the most important predictors of cardiovascular events, ranking just below age and cystatin. The inclusion of the main variables of each dataset allowed us not only to identify the most significant ones but also their interpretation.

Several limitations in this research deserve consideration. The first is the observational nature of the study, which limits the assessment of causality in the reported associations. In any case, a large body of evidence supports the relationship between several factors of the global model and cardiovascular events, such as sex, age, or other parameters such as cystatin. However, other parameters such as impedance, urea, or metabolomic parameters should be evaluated in future studies to confirm these associations. This study is based on a population from the UK, representing people over 40 years of age. Extending this model to other populations will confirm the high precision and sensitivity of our ML models. Finally, the large heterogeneity of the dietary data should be mentioned, with very wide ranges in the food intakes, ranging from no intake at all to intakes above 3 standard deviations from the mean, which probably influenced the results obtained and the absence of dietary parameters in the final model. Another possible cause is the collinearity with other metabolic parameters, so the influence of these dietary factors may underlie the final overall model.

## CONCLUSION

Machine learning (ML) models for cardiovascular disease prediction, combining biochemical, metabolomic, dietary, physical, and lifestyle data, outperform traditional models based solely on clinical variables. The accuracy, precision, and sensitivity of the ML models obtained in both the training and validation samples suggest that the models are robust and may enable future clinical decisions that consider both molecular, social and lifestyle factors. More specifically, this framework highlights the critical role of lifestyle-related variables, such as overall health perception and employment status, alongside other well-defined predictors of CVD events, including cystatin C, testosterone or glycated haemoglobin. The superior performance of ML models compared to traditional logistic approaches underscores the value of integrating diverse data sources for cardiovascular risk prediction and suggests that similar strategies could be applied to other complex health conditions.

## METHODS

### Study design

Between 2006-2010, the UK Biobank recruited 502,134 individuals aged 37 to 73 years from 22 assessment centres located across England, Scotland, and Wales (see: https://www.ukbiobank.ac.uk/ for further information). During their baseline visit at the assessment centre, participants completed a touchscreen questionnaire and underwent a face-to-face interview to collect relevant information, including self-reported sociodemographic factors, health behaviours and medical history. Medical history and comorbidities were verified during face-to-face interviews. To determine the level of deprivation, the Townsend area deprivation index was calculated using aggregated data on unemployment, car and home ownership, and household overcrowding, utilizing the participants’ residential postcode. Trained clinical staff collected various measurements, such as height, weight, blood pressure and biological samples (blood, saliva and urine). The UK Biobank study was conducted according to the Declaration of Helsinki, and ethical approval was granted by the North West Multi-Centre Research Ethics Committee (reference number 06/MRE08/65). At recruitment, all participants gave informed consent to participate and be followed up through data-linkage. UK Biobank protocols and study details can be found elsewhere (https://www.ukbiobank.ac.uk/wp-content/uploads/2025/01/Main-study-protocol.pdf) ^41^.

### Outcome definition

The main outcome included major cardiovascular disease events, defined as fatal or non-fatal angina, myocardial infarction, chronic ischaemic heart disease, atrial fibrillation, heart failure, and stroke (ICD-10 (international classification of diseases, 10th revision) codes I20, I21, I25, I48, I50, I60, I61, I63, and I64). The follow-up period started 1 year after the date they attended the baseline assessment centre and ended on the 31 December 2024. From the initial 502,134 recruited participants, 26,546 were excluded for having incident cardiovascular disease (CVD) (up to 1 year after their baseline assessment).

### Lifestyle and Sociodemographic Information

Socio-demographic variables included information about age, sex, ethnicity (classified as white ethnicity and other ethnicities), etc. This category also includes information about alcohol intake, physical activity, sexual factors, sleep, smoking, social support, sun exposure. Physical activity levels over a typical week were self-reported using an internationally validated physical activity questionnaire. The analysis employed the total metabolic equivalent of tasks (MET) per week. This study included only baseline assessments.

### Dietary Data

Towards the end of the baseline assessment period (April 2009–September 2010), UK Biobank initiated the collection of comprehensive dietary intake data using a web-based self-administered 24-hour dietary assessment tool (WebQ). Among the participants attending their baseline assessment, 70,724 individuals completed the WebQ. Furthermore, from 2011 to 2012, all participants with valid email addresses (n = 331,013) were invited to complete the WebQ on four separate occasions. These assessments took place every 3-4 months on different days to capture seasonal and day-to-day variations. Of those contacted via email, approximately 53% of participants (n = 176,012) completed at least one assessment, resulting in a total of 211,050 participants who completed at least one dietary assessment either online or during the baseline assessment^43^.The WebQ collects information regarding the consumption of foods and beverages during the previous day. Participants were presented with a list of 206 commonly consumed foods and 32 beverages in the UK and were asked to indicate the number of portions consumed for each item. This comprehensive food list was compiled using data from the UK National Diet and Nutrition Survey (NDNS), and from a pilot study ^44^.

To calculate total energy and nutrient intakes, the number of portions consumed was multiplied by the predetermined quantity of each food portion size and its corresponding nutrient composition, obtained from the UK Nutrient Databank Food Composition Tables (FCT) for survey years 6 (2012-2013 and 2013-2014)^45^. Dietary fiber content was determined using the Englyst method, which includes non-starch polysaccharides but excludes lignin and resistant starches ^46^.

In addition to macro- and micronutrient composition, in the present study, a comprehensive food group classification system for the 206 foods and 32 beverages reported in the Oxford WebQ was employed, as previously described ^47^. These items were grouped into 93 distinct categories, consisting of 79 food groups and 14 beverage groups, which belonged to 15 primary food categories (13 food and 2 beverage categories).

### Physical Measures

Height was measured with a stadiometer (Seca 202; Hamburg, Germany) to the nearest centimetre, while body weight was measured using a body composition analyser (Tanita BC-418; Tokyo, Japan) to the nearest 0.1 kg. Body mass index (BMI) was calculated as weight (kg) divided by the square of height (m^2).

Other variables included in the present study were: arterial stiffness, blood pressure, bone-densitometry of heel, carotid ultrasound, and hand grip strength, as well as spirometry parameters, were included. The pulse waveform obtained at the finger with an infrared sensor was evaluated with a PulseTrace PCA2 (CareFusion, USA). Blood pressure was determined with an Omron 705 IT electronic blood pressure monitor (OMRON Healthcare Europe B.V. Hoofddorp, NL). The mean of at least two measurements was taken as the blood pressure value. Body composition was evaluated by bioimpedance with a Tanita BC418MA body composition analyser (TANITA Europe B.V. Manchester, UK). A Sahara Heel Ultrasound device (Hologic, USA) was used to evaluate bone densitometry parameters. For carotid ultrasound analyses, images were acquired using a CardioHealth Station (Panasonic Biomedical Sales Europe BV, Leicestershire, UK), which has a linear array transducer and a frequency of 5-13 MHz. Hand grip strength was determined with a Jamar J00105 hydraulic hand dynamometer (Lafayette Instrument, US). Finally, breath spirometry was performed using a Vitalograph Pneumotrac 6800 (Vitalograph, UK).

### Clinical biomarkers

At recruitment, non-fasting blood samples were collected in EDTA collection vessels, which were processed on automation systems, and standard haematological tests were performed on fresh whole blood within 24 hours of blood collection for all the participants (https://biobank.ndph.ox.ac.uk/showcase/refer.cgi?id=5636).

We used data on features of blood count from haematological assays, including cell components like lymphocytes (basophils, eosinophils, and neutrophils), monocytes, haematocrit, erythrocytes, and platelets, as well as other biomarkers like reticulocytes, mean sphered cell volume, and mean corpuscular volume. Blood biochemistry features included 34 measures such as aminotransferases, lipid biomarkers, apolipoproteins, glucose, among others.

### Metabolomic Data

Technical details have been previously described (https://biobank.ndph.ox.ac.uk/showcase/ukb/docs/NMR_companion_phase2.pdf) ^42^

Metabolomic data were available from approximately 250,000 baseline participants. A high-throughput nuclear magnetic resonance (NMR) platform (Nightingale Health Plc, Finland) was used to quantify a total of 249 metabolic measures in EDTA plasma samples, comprising 168 measures in absolute levels (mmol/L) and 81 ratio measures. The biomarkers include comprehensive assessments of cholesterol metabolism, fatty acid compositions, and various low-molecular-weight metabolites, such as amino acids, ketones, and glycolysis metabolites. Additionally, lipid concentrations and compositions, including triglycerides, phospholipids, total cholesterol, cholesterol esters, free cholesterol, and total lipid concentration, were measured for 14 lipoprotein subclasses.

## STATISTICAL ANALYSIS

Variables are described as mean ± SD or median and interquartile range for non-normally distributed variables. Categorical variables were expressed as percentages. Comparison among subjects with or without a CVD event was done by χ2 test (for categorical variables), and by a Student’s t test or the non-parametric Mann-Whitney U test, depending on the data distribution. Multivariable logistic regression models were conducted for each type of data separately (clinical, diet, lifestyle, etc.), using CVD event as outcome variable. For variable selection in the logistic regression model, a stepwise selection method based on the Akaike Information Criterion (stepAIC) was used (library MASS). This method balances the goodness of fit with the complexity of the model, penalising those with too many variables. Nonsignificant variables were eliminated from the model if they did not contribute to improving the fit according to the AIC. The features significantly associated with CVD were described as odds ratio (OR) and 95% confidence interval (95%CI). We apply the Benjamini-Hochberg method to adjust the p-values and control the false discovery rate. All tests were two-sided, and a p < 0.050 was considered statistically significant for all analyses using R software (release 4.2.0).

## MACHINE LEARNING APPROACH

### Feature Definitions

The variables used as input features for the ML models correspond to the biochemical, metabolomic, dietary, lifestyle, physical and sociodemographic domains described previously. Each feature was preprocessed and standardized according to the procedure outlined in the Data Preprocessing subsection. No additional feature engineering was applied beyond encoding, normalization, and imputation, and all variables meeting the inclusion criteria were used for model training.

### Data Preprocessing

Data preprocessing is a critical step before training the model. Its main goal is to enhance data interpretability and ensure that the dataset is complete, consistent, and manageable within time and resource constraints. Figure 7 illustrates the entire preprocessing workflow applied to each dataset, where nominal attributes are categorized and represented as integers.

**Figure 7:**
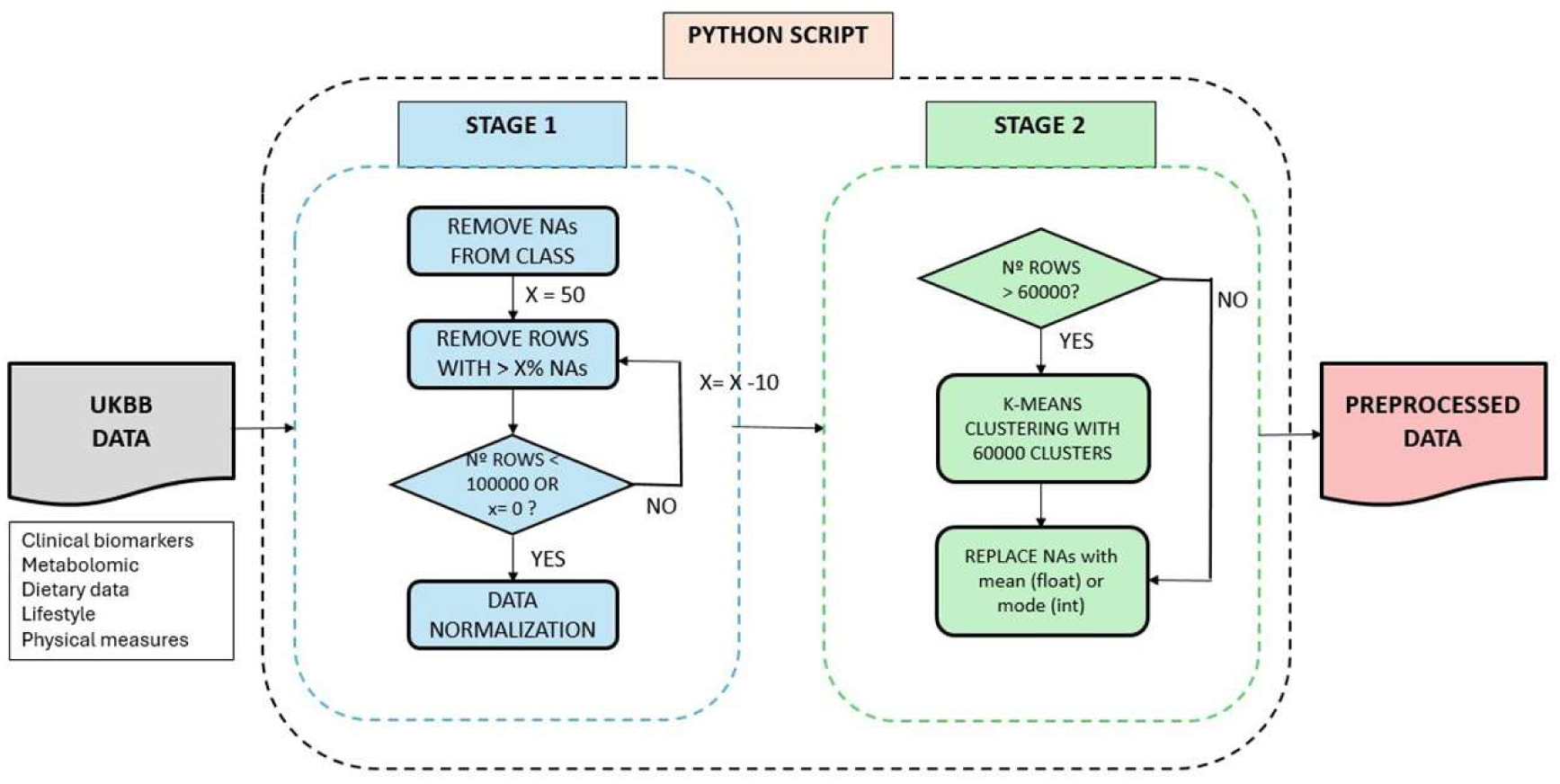
Preprocessing Workflow for UK Biobank Data. This figure shows the two-stage data preprocessing pipeline implemented using a Python script. Stage 1: Rows with unknown class attributes and >50% missing data are removed, decreasing this value to reduce to a manageable number of rows; normalization follows. Stage 2: K-means clustering (60,000 clusters) reduces dataset size, and the remaining missing values are imputed with the mean (continuous) or mode (categorical), producing a finalized dataset ready for machine learning.

The data cleaning process was carried out in two stages: initial cleaning and refinement. In the first stage, rows with an unknown class attribute were removed to retain only well-defined data points. Records with more than 50% missing values were deleted, and when the dataset still exceeded 100,000 rows, this threshold was adjusted to maintain a manageable size. The remaining data were then normalized to standardize feature scales and ensure compatibility with ML algorithms.

In the refinement stage, K-means clustering with 60,000 clusters was applied to reduce the dataset size without losing essential patterns, improving manageability for downstream analysis. Remaining missing values were imputed according to variable type: continuous features were filled with the mean, while categorical features were replaced with the mode. These combined preprocessing steps, summarized in Figure 7, effectively prepared the UK Biobank data by removing incomplete entries, standardizing scales, and imputing missing information, yielding a dataset suitable for robust and reproducible machine learning analysis.

### Model Training and Evaluation

#### Training and Testing

This study utilised an integrative approach to assess the factors contributing to cardiovascular diseases (CVDs). Recognizing CVDs as complex physiological disorders influenced by a host of genetic and non-genetic factors, such as diet, physical activity, and socio-demographic characteristics, we undertook a comprehensive investigation. We collected a multidimensional dataset encompassing clinical, socio-demographical, dietary, and physical activity profiles from half a million participants over a 15-year period. The occurrence of cardiovascular events, including heart failure, stroke and cardiac arrest, was monitored within this population cohort.

To address the challenge of personalized medicine and to make sense of this large and diverse dataset, we employed SIBILA (https://github.com/bio-hpc/sibila), an ensemble of machine learning and deep learning models. This state-of-the-art tool incorporates a variety of interpretability algorithms to identify and assign relevance to input features contributing to cardiovascular outcomes. A consensus stage within SIBILA ensures a balanced estimation of the global attribution of each variable to the predictions. Despite being a sophisticated tool, it has been containerized for compatibility with any high-performance computing platform and is accessible via a user-friendly web server, allowing users with minimal technological skills to leverage its capabilities.

SIBILA’’s strength lies not only in its ability to classify and predict but also in its transparency. Its unique architecture offers insights into the internal workings of the model, thus facilitating a deeper understanding of the associations between various factors and cardiovascular outcomes.

In the current study, SIBILA was applied with the aim of determining its viability as a robust decision-making tool for clinicians, elucidating the intricate web of factors contributing to CVDs. The results from this study not only enhance our understanding of the etiology of CVDs, but also highlight the potential for leveraging machine learning and deep learning tools like SIBILA in the pursuit of personalized medicine.

#### Machine Learning Methods Used

As shown in the preprocessing workflow (Figure 7), the analysis followed a two-stage pipeline. First, records with undefined outcomes or with more than 50% missing data were excluded. The remaining features were normalised, categorical variables encoded and missing values imputed using the mean (for continuous data) or mode (for categorical data). To further reduce dimensionality while preserving structure, k-means clustering was applied, and redundant or highly correlated variables were removed.

Model training was carried out using the SIBILA framework, which allows parallel evaluation of multiple algorithms under a unified pipeline. In line with our previous studies, we trained artificial neural networks, random forests, and gradient boosting models, comparing their performance on an 80/20 train–validation split. Model accuracy was primarily assessed through ROC-AUC, with sensitivity, specificity, precision and overall accuracy reported as secondary metrics. Finally, the most predictive features from each domain (biomarkers, metabolomics, diet, lifestyle, physical measures and sociodemographics) were integrated into a single multidimensional model, which consistently outperformed domain-specific approaches.

#### Interpretability techniques applied

Model interpretability was addressed using Shapley additive explanations (SHAP), applied in accordance with the workflow illustrated in Figure 8. Global SHAP analyses revealed the ranked importance of features across the cohort, uncovering both monotonic and non-linear associations, while individual SHAP profiles clarified how particular variables contributed to the risk estimate of a given participant. These models will be used to make predictions at the individual level (for each participant), as shown in the examples in the figures in the results section. The consensus stage of SIBILA ensured that these explanations remained stable across competing models, providing clinicians and researchers with transparent and reproducible insights into the predictions.

## STATEMENTS

### Financial Support

The UK Biobank was supported by the Wellcome Trust, Medical Research Council, Department of Health, Scottish Government, the Northwest Regional Development Agency, the Welsh Assembly Government and the British Heart Foundation. This research has been conducted using the UK Biobank resource under application number 124330. H. P.S. and C. M.-C. acknowledge funding by the European Project Horizon 2020 SC1-BHC-02-2019 [RE-VERT, ID: 848098]. C.P. is funded by RYC2020-028818-I (MCIN/AEI/10.13039/501100011033 and “ESF Investing in your future”, Ministry of Science and Innovation, Spain), which also paid for the access to the data. This research was designed, conducted, analysed, and interpreted by the authors entirely independently of the funding sources.

### Author Contributions

J. J. H. M. and E. M. G. performed data curation and integration, statistical modelling, analysing the results, and contributed to writing the manuscript. H. P. S. and C.M.C. contributed to data query and programming, to the development of machine learning models, and writing the manuscript. C. P. contributed to the conceptual design of the study, data access and writing the manuscript.

### Competing interests

The authors declare no competing interests.

### Materials & Correspondence

Correspondence and requests for materials should be addressed to the H.P.S.

### Data Availability

The authors declare that all data supporting the findings of this study are available upon request from UK Biobank (https://www.ukbiobank.ac.uk/). The raw UK Biobank data are made available to researchers from universities and other research institutions with research inquiries following IRB and UK Biobank approval (https://www.ukbiobank.ac.uk/enable-your-research/apply-for-access).

### Code Availability

The Shell and Python source code used to run the analysis described in the article and to generate all figures is available at: https://github.com/bio-hpc/sibila

## Notes

### Competing Interest Statement

The authors have declared no competing interest.

