## Supplementary Information for "A MULTI-DIMENSIONAL MACHINE LEARNING APPROACH FOR CARDIOVASCULAR DISEASE PREDICTION IN THE UK BIOBANK STUDY"

***Multi-dimensional machine learning approach to predict cardiovascular events in the UK Biobank cohort***

Hernández-Morante et al.

**Supplementary Table S1.** Biomarkers data attending to the presence or not of a cardiovascular event during the study period.

| Variable | No CVD (n=422192) | | CVD (n=79942) | | t | p value | Cohen d | FDR |
|---|---|---|---|---|---|---|---|---|
| | Mean | sd | Mean | sd | | | | |
| Albumin | 45.28 | 2.61 | 44.84 | 2.71 | 39.44 | < 0.0001 | -0.17 | < 0.0001 |
| Alkaline phosphatase | 82.87 | 25.84 | 87.95 | 29.17 | -44.35 | < 0.0001 | 0.19 | < 0.0001 |
| ALT | 23.20 | 14.01 | 25.40 | 14.91 | -37.21 | < 0.0001 | 0.16 | < 0.0001 |
| Apolipoprotein A | 1.55 | 0.27 | 1.47 | 0.26 | 77.97 | < 0.0001 | -0.32 | < 0.0001 |
| Apolipoprotein B | 1.04 | 0.23 | 0.99 | 0.26 | 46.74 | < 0.0001 | -0.20 | < 0.0001 |
| AST | 25.96 | 10.47 | 27.66 | 11.54 | -37.22 | < 0.0001 | 0.16 | < 0.0001 |
| Direct bilirubin | 1.81 | 0.83 | 1.94 | 0.95 | -31.40 | < 0.0001 | 0.15 | < 0.0001 |
| Urea | 5.33 | 1.30 | 5.80 | 1.79 | -67.82 | < 0.0001 | 0.33 | < 0.0001 |
| Calcium | 2.38 | 0.09 | 2.38 | 0.10 | 8.05 | < 0.0001 | -0.03 | < 0.0001 |
| Cholesterol | 5.76 | 1.11 | 5.34 | 1.27 | 85.17 | < 0.0001 | -0.37 | < 0.0001 |
| Creatinine | 71.27 | 15.56 | 77.83 | 29.15 | -59.87 | < 0.0001 | 0.36 | < 0.0001 |
| CRP | 2.47 | 4.17 | 3.28 | 5.21 | -39.71 | < 0.0001 | 0.18 | < 0.0001 |
| Cystatin C | 0.89 | 0.15 | 0.99 | 0.26 | -103.10 | < 0.0001 | 0.58 | < 0.0001 |
| GGT | 35.79 | 39.39 | 45.88 | 53.40 | -49.07 | < 0.0001 | 0.24 | < 0.0001 |
| Glucose | 5.07 | 1.11 | 5.40 | 1.75 | -47.70 | < 0.0001 | 0.27 | < 0.0001 |
| HbA1c | 35.64 | 6.05 | 38.72 | 9.34 | -86.24 | < 0.0001 | 0.46 | < 0.0001 |
| HDL cholesterol | 1.47 | 0.38 | 1.32 | 0.36 | 98.67 | < 0.0001 | -0.40 | < 0.0001 |
| IGF 1 | 21.57 | 5.66 | 20.51 | 5.83 | 45.60 | < 0.0001 | -0.19 | < 0.0001 |
| LDL direct | 3.60 | 0.85 | 3.33 | 0.96 | 73.16 | < 0.0001 | -0.32 | < 0.0001 |
| Lipoprotein A | 44.02 | 48.76 | 48.06 | 51.48 | -17.53 | < 0.0001 | 0.08 | < 0.0001 |
| Oestradiol | 475.64 | 441.69 | 324.03 | 278.32 | 41.54 | < 0.0001 | -0.35 | < 0.0001 |
| Phosphate | 1.16 | 0.16 | 1.15 | 0.17 | 8.88 | < 0.0001 | -0.04 | < 0.0001 |
| Rheumatoid factor | 24.37 | 19.64 | 25.48 | 20.88 | -4.17 | < 0.0001 | 0.06 | < 0.0001 |
| SHBG | 52.58 | 28.31 | 46.59 | 24.14 | 57.54 | < 0.0001 | -0.22 | < 0.0001 |
| Total bilirubin | 9.08 | 4.41 | 9.34 | 4.51 | -14.40 | < 0.0001 | 0.06 | < 0.0001 |
| Testosterone | 6.25 | 6.06 | 8.17 | 5.78 | -78.53 | < 0.0001 | 0.32 | < 0.0001 |
| Total protein | 72.54 | 4.09 | 72.38 | 4.26 | 9.04 | < 0.0001 | -0.04 | < 0.0001 |
| Triglycerides | 1.71 | 1.01 | 1.95 | 1.11 | -54.77 | < 0.0001 | 0.23 | < 0.0001 |
| Urate | 303.84 | 78.48 | 337.76 | 84.54 | -101.46 | < 0.0001 | 0.43 | < 0.0001 |
| Vitamin D | 48.78 | 21.05 | 47.73 | 21.39 | 12.08 | < 0.0001 | -0.05 | < 0.0001 |

ALT: alanine aminotransferase. AST: aspartate aminotransferase. CRP: C-reactive protein. GGT: gamma-globulin transferase. IGF-1: insulin-like growth factor-1. SHBG: sex-hormone binding globulin.

**Supplementary Table S2.** Model 4. StepAIC regression model after debugging the model for biomarkers variables with high multicollinearity (VIF >5).

| Predictor | Estimate | Std Error | Z | p | OR | Lower CI | Upper CI | FDR |
|---|---|---|---|---|---|---|---|---|
| (Intercept) | -2.11 | 0.08 | -25.75 | <0.0001 | 0.12 | 0.10 | 0.14 | <0.0001 |
| Albumin | 0.02 | 0.08 | 0.22 | 0.8236 | 1.02 | 0.88 | 1.18 | 0.9196 |
| Alkaline phosphatase | 0.01 | 0.06 | 0.14 | 0.8856 | 1.01 | 0.90 | 1.13 | 0.9196 |
| ALT | 0.04 | 0.09 | 0.43 | 0.6660 | 1.04 | 0.87 | 1.24 | 0.8991 |
| Apolipoprotein A | -0.23 | 0.07 | -3.03 | 0.0025 | 0.80 | 0.69 | 0.92 | 0.0096 |
| Apolipoprotein B | -0.12 | 0.07 | -1.72 | 0.0848 | 0.88 | 0.77 | 1.02 | 0.1909 |
| AST | -0.05 | 0.08 | -0.60 | 0.5464 | 0.95 | 0.81 | 1.10 | 0.8197 |
| C reactive protein | -0.01 | 0.05 | -0.15 | 0.8798 | 0.99 | 0.90 | 1.09 | 0.9196 |
| Calcium | -0.02 | 0.07 | -0.27 | 0.7881 | 0.98 | 0.85 | 1.13 | 0.9196 |
| Creatinine | -0.02 | 0.10 | -0.20 | 0.8437 | 0.98 | 0.81 | 1.19 | 0.9196 |
| Cystatin C | 0.37 | 0.08 | 4.52 | <0.0001 | 1.45 | 1.23 | 1.70 | 0.0001 |
| Direct bilirubin | 0.20 | 0.06 | 3.57 | 0.0004 | 1.22 | 1.09 | 1.36 | 0.0016 |
| GGT | 0.18 | 0.06 | 2.96 | 0.0031 | 1.19 | 1.07 | 1.35 | 0.0104 |
| Glucose | -0.05 | 0.07 | -0.67 | 0.5029 | 0.95 | 0.83 | 1.10 | 0.7987 |
| HbA1c | 0.30 | 0.07 | 4.24 | <0.0001 | 1.36 | 1.18 | 1.56 | 0.0002 |
| IGF 1 | -0.26 | 0.07 | -3.79 | 0.0002 | 0.77 | 0.67 | 0.88 | 0.0008 |
| Lipoprotein A | 0.12 | 0.06 | 2.17 | 0.0301 | 1.13 | 1.01 | 1.26 | 0.0740 |
| Oestradiol | -0.19 | 0.12 | -1.64 | 0.1001 | 0.83 | 0.65 | 1.02 | 0.2078 |
| Phosphate | 0.03 | 0.06 | 0.54 | 0.5871 | 1.04 | 0.91 | 1.17 | 0.8343 |
| Rheumatoid factor | 0.06 | 0.05 | 1.15 | 0.2503 | 1.06 | 0.96 | 1.18 | 0.4828 |
| SHBG | 0.00 | 0.07 | 0.04 | 0.9668 | 1.00 | 0.87 | 1.16 | 0.9668 |
| Testosterone | 0.24 | 0.06 | 3.88 | 0.0001 | 1.28 | 1.13 | 1.44 | 0.0007 |
| Total protein | -0.02 | 0.07 | -0.31 | 0.7554 | 0.98 | 0.86 | 1.12 | 0.9196 |
| Triglycerides | 0.07 | 0.08 | 0.83 | 0.4055 | 1.07 | 0.91 | 1.24 | 0.7299 |
| Urate | 0.18 | 0.08 | 2.34 | 0.0191 | 1.20 | 1.03 | 1.39 | 0.0574 |
| Urea | 0.16 | 0.07 | 2.21 | 0.0272 | 1.18 | 1.02 | 1.36 | 0.0734 |
| Vitamin D | 0.04 | 0.06 | 0.70 | 0.4843 | 1.04 | 0.93 | 1.17 | 0.7987 |

Overall model performance: $R^2_{McFadden}$: 0.191. $R^2_{Nagelkerke}$: 0.252. AIC: 2007.04. Chi-cuadrado global: 438266.3, df: 498924, p = 1.

**Supplementary Table S3.** Metabolomic data attending to the presence or not of a cardiovascular event during the study period.

| | No CVD (n=207617) | | CVD (n=40306) | | t | p value | Cohen d | FDR |
|---|---|---|---|---|---|---|---|---|
| **Variable** | **Mean** | **sd** | **Mean** | **sd** | | | | |
| Acetate | 0.02 | 0.01 | 0.02 | 0.01 | 4.36 | < 0.0001 | -0.03 | < 0.0001 |
| Acetoacetate | 0.01 | 0.01 | 0.01 | 0.01 | -14.11 | < 0.0001 | 0.08 | < 0.0001 |
| Acetone | 0.01 | 0.01 | 0.01 | 0.01 | -9.21 | < 0.0001 | 0.05 | < 0.0001 |
| Alanine | 0.30 | 0.08 | 0.31 | 0.08 | -25.14 | < 0.0001 | 0.14 | < 0.0001 |
| Albumin | 39.45 | 3.32 | 38.84 | 3.42 | 32.82 | < 0.0001 | -0.18 | < 0.0001 |
| Apolipoprotein A1 | 1.46 | 0.24 | 1.39 | 0.24 | 58.37 | < 0.0001 | -0.32 | < 0.0001 |
| Apolipoprotein B | 0.86 | 0.20 | 0.81 | 0.21 | 44.93 | < 0.0001 | -0.26 | < 0.0001 |
| Apolipoprotein B/Apolipoprotein A1 | 0.60 | 0.16 | 0.60 | 0.17 | 8.74 | < 0.0001 | -0.05 | < 0.0001 |
| Average Diameter for HDL Particles | 9.64 | 0.20 | 9.57 | 0.18 | 66.29 | < 0.0001 | -0.34 | < 0.0001 |
| Average Diameter for LDL Particles | 23.93 | 0.09 | 23.91 | 0.09 | 48.30 | < 0.0001 | -0.27 | < 0.0001 |
| Average Diameter for VLDL Particles | 38.67 | 1.19 | 38.95 | 1.22 | -42.16 | < 0.0001 | 0.23 | < 0.0001 |
| BCAA | 0.37 | 0.09 | 0.38 | 0.09 | -31.99 | < 0.0001 | 0.18 | < 0.0001 |
| Chol. in Chylomicrons | 0.06 | 0.04 | 0.06 | 0.04 | -26.90 | < 0.0001 | 0.15 | < 0.0001 |
| Chol. in IDL | 0.85 | 0.21 | 0.76 | 0.23 | 75.14 | < 0.0001 | -0.44 | < 0.0001 |
| Chol. in Large HDL | 0.30 | 0.16 | 0.24 | 0.14 | 78.12 | < 0.0001 | -0.39 | < 0.0001 |
| Chol. in Large LDL | 1.16 | 0.28 | 1.06 | 0.31 | 61.65 | < 0.0001 | -0.36 | < 0.0001 |
| Chol. in Large VLDL | 0.10 | 0.05 | 0.10 | 0.05 | -12.03 | < 0.0001 | 0.07 | < 0.0001 |
| Chol. in Medium HDL | 0.50 | 0.12 | 0.46 | 0.12 | 57.53 | < 0.0001 | -0.31 | < 0.0001 |
| Chol. in Medium LDL | 0.44 | 0.12 | 0.41 | 0.13 | 40.78 | < 0.0001 | -0.23 | < 0.0001 |
| Chol. in Medium VLDL | 0.18 | 0.06 | 0.16 | 0.07 | 54.25 | < 0.0001 | -0.31 | < 0.0001 |
| Chol. in Small HDL | 0.45 | 0.06 | 0.44 | 0.06 | 30.81 | < 0.0001 | -0.17 | < 0.0001 |
| Chol. in Small LDL | 0.18 | 0.04 | 0.17 | 0.05 | 43.76 | < 0.0001 | -0.25 | < 0.0001 |
| Chol. in Small VLDL | 0.16 | 0.05 | 0.15 | 0.05 | 22.59 | < 0.0001 | -0.13 | < 0.0001 |
| Chol. in Very Large HDL | 0.08 | 0.03 | 0.07 | 0.03 | 77.75 | < 0.0001 | -0.39 | < 0.0001 |
| Chol. in Very Large VLDL | 0.06 | 0.03 | 0.06 | 0.03 | -11.64 | < 0.0001 | 0.06 | < 0.0001 |
| Chol. in Very Small VLDL | 0.19 | 0.05 | 0.17 | 0.05 | 55.36 | < 0.0001 | -0.32 | < 0.0001 |
| Chol./T.Lipids in Chylomicrons | 30.24 | 13.11 | 28.96 | 12.19 | 19.09 | < 0.0001 | -0.10 | < 0.0001 |
| Chol./T.Lipids in IDL | 67.87 | 2.80 | 66.31 | 3.48 | 84.45 | < 0.0001 | -0.53 | < 0.0001 |
| Chol./T.Lipids in Large HDL | 44.10 | 5.20 | 41.64 | 6.45 | 72.03 | < 0.0001 | -0.45 | < 0.0001 |
| Chol./T.Lipids in Large LDL | 71.26 | 1.73 | 70.39 | 2.34 | 71.37 | < 0.0001 | -0.48 | < 0.0001 |
| Chol./T.Lipids in Large VLDL | 30.25 | 4.03 | 29.31 | 4.18 | 41.30 | < 0.0001 | -0.23 | < 0.0001 |
| Chol./T.Lipids in Medium HDL | 47.59 | 2.88 | 46.55 | 2.97 | 64.39 | < 0.0001 | -0.36 | < 0.0001 |
| Chol./T.Lipids in Medium LDL | 68.52 | 1.56 | 67.81 | 2.01 | 67.59 | < 0.0001 | -0.44 | < 0.0001 |
| Chol./T.Lipids in Medium VLDL | 30.20 | 6.46 | 27.02 | 7.21 | 82.45 | < 0.0001 | -0.48 | < 0.0001 |
| Chol./T.Lipids in Small HDL | 38.41 | 1.83 | 37.94 | 1.91 | 45.80 | < 0.0001 | -0.26 | < 0.0001 |
| Chol./T.Lipids in Small LDL | 63.37 | 1.89 | 62.58 | 2.31 | 64.30 | < 0.0001 | -0.40 | < 0.0001 |
| Chol./T.Lipids in Small VLDL | 38.16 | 4.53 | 36.26 | 4.96 | 71.07 | < 0.0001 | -0.41 | < 0.0001 |
| Chol./T.Lipids in Very Large HDL | 50.20 | 4.86 | 50.57 | 5.57 | -12.59 | < 0.0001 | 0.08 | < 0.0001 |
| Chol./T.Lipids in Very Large VLDL | 29.30 | 7.23 | 26.91 | 6.48 | 66.35 | < 0.0001 | -0.34 | < 0.0001 |
| Chol./T.Lipids in Very Small VLDL | 51.44 | 4.46 | 48.86 | 5.18 | 93.55 | < 0.0001 | -0.56 | < 0.0001 |
| CE in Chylomicrons | 0.03 | 0.02 | 0.03 | 0.02 | -23.72 | < 0.0001 | 0.13 | < 0.0001 |
| CE in HDL | 1.03 | 0.25 | 0.94 | 0.23 | 75.16 | < 0.0001 | -0.39 | < 0.0001 |
| CE in IDL | 0.63 | 0.15 | 0.56 | 0.17 | 75.76 | < 0.0001 | -0.45 | < 0.0001 |
| CE in Large HDL | 0.23 | 0.13 | 0.18 | 0.11 | 78.66 | < 0.0001 | -0.39 | < 0.0001 |
| CE in Large LDL | 0.86 | 0.21 | 0.79 | 0.23 | 57.87 | < 0.0001 | -0.33 | < 0.0001 |
| CE in Large VLDL | 0.05 | 0.02 | 0.05 | 0.02 | 0.04 | 0.9691 | 0.00 | 0.9691 |
| CE in LDL | 1.30 | 0.32 | 1.21 | 0.35 | 50.52 | < 0.0001 | -0.29 | < 0.0001 |
| CE in Medium HDL | 0.41 | 0.10 | 0.38 | 0.09 | 56.96 | < 0.0001 | -0.31 | < 0.0001 |
| CE in Medium LDL | 0.31 | 0.09 | 0.30 | 0.09 | 34.36 | < 0.0001 | -0.20 | < 0.0001 |
| CE in Medium VLDL | 0.10 | 0.04 | 0.08 | 0.04 | 65.00 | < 0.0001 | -0.38 | < 0.0001 |
| CE in Small HDL | 0.33 | 0.05 | 0.33 | 0.05 | 29.47 | < 0.0001 | -0.16 | < 0.0001 |
| CE in Small LDL | 0.13 | 0.03 | 0.13 | 0.04 | 37.58 | < 0.0001 | -0.22 | < 0.0001 |
| CE in Small VLDL | 0.10 | 0.03 | 0.10 | 0.03 | 15.22 | < 0.0001 | -0.08 | < 0.0001 |
| CE in Very Large HDL | 0.06 | 0.03 | 0.05 | 0.02 | 79.50 | < 0.0001 | -0.40 | < 0.0001 |
| CE in Very Large VLDL | 0.03 | 0.01 | 0.03 | 0.01 | -1.21 | 0.2270 | 0.01 | 0.2297 |
| CE in Very Small VLDL | 0.13 | 0.03 | 0.12 | 0.04 | 62.68 | < 0.0001 | -0.36 | < 0.0001 |
| CE in VLDL | 0.44 | 0.14 | 0.41 | 0.15 | 33.66 | < 0.0001 | -0.19 | < 0.0001 |
| CE/T.Lipids in Chylomicrons | 17.29 | 9.79 | 16.25 | 8.89 | 21.08 | < 0.0001 | -0.11 | < 0.0001 |
| CE/T.Lipids in IDL | 50.11 | 2.29 | 48.86 | 2.79 | 84.53 | < 0.0001 | -0.53 | < 0.0001 |
| CE/T.Lipids in Large HDL | 34.08 | 4.52 | 32.08 | 5.47 | 68.79 | < 0.0001 | -0.43 | < 0.0001 |
| CE/T.Lipids in Large LDL | 52.70 | 1.25 | 52.45 | 1.57 | 29.90 | < 0.0001 | -0.19 | < 0.0001 |

| | | | | | | | | |
|---|---|---|---|---|---|---|---|---|
| CE/T.Lipids in Large VLDL | 16.33 | 3.23 | 15.29 | 3.32 | 57.59 | < 0.0001 | -0.32 | < 0.0001 |
| CE/T.Lipids in Medium HDL | 39.26 | 2.52 | 38.51 | 2.64 | 51.96 | < 0.0001 | -0.29 | < 0.0001 |
| CE/T.Lipids in Medium LDL | 49.24 | 1.83 | 49.20 | 1.89 | 3.66 | 0.0003 | -0.02 | 0.0003 |
| CE/T.Lipids in Medium VLDL | 16.44 | 4.86 | 14.03 | 5.35 | 83.73 | < 0.0001 | -0.49 | < 0.0001 |
| CE/T.Lipids in Small HDL | 28.48 | 1.83 | 28.14 | 1.91 | 32.74 | < 0.0001 | -0.18 | < 0.0001 |
| CE/T.Lipids in Small LDL | 45.98 | 2.00 | 45.79 | 2.14 | 16.89 | < 0.0001 | -0.10 | < 0.0001 |
| CE/T.Lipids in Small VLDL | 23.73 | 2.69 | 22.92 | 2.92 | 51.83 | < 0.0001 | -0.30 | < 0.0001 |
| CE/T.Lipids in Very Large HDL | 35.09 | 3.32 | 34.25 | 4.06 | 39.28 | < 0.0001 | -0.24 | < 0.0001 |
| CE/T.Lipids in Very Large VLDL | 17.28 | 6.00 | 15.34 | 5.30 | 65.60 | < 0.0001 | -0.33 | < 0.0001 |
| CE/T.Lipids in Very Small VLDL | 35.38 | 4.02 | 32.98 | 4.64 | 96.61 | < 0.0001 | -0.58 | < 0.0001 |
| Citrate | 0.07 | 0.01 | 0.07 | 0.01 | -7.74 | < 0.0001 | 0.04 | < 0.0001 |
| Clinical LDL Chol. | 2.60 | 0.72 | 2.35 | 0.80 | 60.51 | < 0.0001 | -0.35 | < 0.0001 |
| Conc. of Chylomicrons Particles | 0.00 | 0.00 | 0.00 | 0.00 | -32.58 | < 0.0001 | 0.18 | < 0.0001 |
| Conc. of HDL Particles | 0.02 | 0.00 | 0.01 | 0.00 | 57.68 | < 0.0001 | -0.32 | < 0.0001 |
| Conc. of IDL Particles | 0.00 | 0.00 | 0.00 | 0.00 | 59.40 | < 0.0001 | -0.35 | < 0.0001 |
| Conc. of Large HDL Particles | 0.00 | 0.00 | 0.00 | 0.00 | 75.93 | < 0.0001 | -0.38 | < 0.0001 |
| Conc. of Large LDL Particles | 0.00 | 0.00 | 0.00 | 0.00 | 48.43 | < 0.0001 | -0.28 | < 0.0001 |
| Conc. of Large VLDL Particles | 0.00 | 0.00 | 0.00 | 0.00 | -24.99 | < 0.0001 | 0.14 | < 0.0001 |
| Conc. of LDL Particles | 0.00 | 0.00 | 0.00 | 0.00 | 44.61 | < 0.0001 | -0.26 | < 0.0001 |
| Conc. of Medium HDL Particles | 0.00 | 0.00 | 0.00 | 0.00 | 53.21 | < 0.0001 | -0.29 | < 0.0001 |
| Conc. of Medium LDL Particles | 0.00 | 0.00 | 0.00 | 0.00 | 36.38 | < 0.0001 | -0.21 | < 0.0001 |
| Conc. of Medium VLDL Particles | 0.00 | 0.00 | 0.00 | 0.00 | 23.89 | < 0.0001 | -0.13 | < 0.0001 |
| Conc. of Small HDL Particles | 0.01 | 0.00 | 0.01 | 0.00 | 26.57 | < 0.0001 | -0.15 | < 0.0001 |
| Conc. of Small LDL Particles | 0.00 | 0.00 | 0.00 | 0.00 | 35.32 | < 0.0001 | -0.20 | < 0.0001 |
| Conc. of Small VLDL Particles | 0.00 | 0.00 | 0.00 | 0.00 | -8.72 | < 0.0001 | 0.05 | < 0.0001 |
| Conc. of Very Large HDL Particles | 0.00 | 0.00 | 0.00 | 0.00 | 74.09 | < 0.0001 | -0.37 | < 0.0001 |
| Conc. of Very Large VLDL Particles | 0.00 | 0.00 | 0.00 | 0.00 | -30.52 | < 0.0001 | 0.17 | < 0.0001 |
| Conc. of Very Small VLDL Particles | 0.00 | 0.00 | 0.00 | 0.00 | 30.75 | < 0.0001 | -0.17 | < 0.0001 |
| Conc. of VLDL Particles | 0.00 | 0.00 | 0.00 | 0.00 | 8.09 | < 0.0001 | -0.05 | < 0.0001 |
| Creatinine | 0.07 | 0.01 | 0.07 | 0.02 | -55.34 | < 0.0001 | 0.39 | < 0.0001 |
| Degree of Unsaturation | 1.36 | 0.08 | 1.34 | 0.08 | 42.37 | < 0.0001 | -0.24 | < 0.0001 |
| DHA | 0.24 | 0.08 | 0.23 | 0.08 | 26.27 | < 0.0001 | -0.14 | < 0.0001 |
| DHA/Total Fatty Acids | 1.98 | 0.65 | 1.91 | 0.67 | 17.60 | < 0.0001 | -0.10 | < 0.0001 |
| Free Chol. in Chylomicrons | 0.02 | 0.02 | 0.03 | 0.02 | -30.27 | < 0.0001 | 0.17 | < 0.0001 |
| Free Chol. in HDL | 0.30 | 0.07 | 0.27 | 0.07 | 71.31 | < 0.0001 | -0.37 | < 0.0001 |
| Free Chol. in IDL | 0.22 | 0.05 | 0.20 | 0.06 | 71.89 | < 0.0001 | -0.42 | < 0.0001 |
| Free Chol. in Large HDL | 0.07 | 0.04 | 0.05 | 0.03 | 75.26 | < 0.0001 | -0.37 | < 0.0001 |
| Free Chol. in Large LDL | 0.30 | 0.08 | 0.27 | 0.08 | 70.36 | < 0.0001 | -0.41 | < 0.0001 |
| Free Chol. in Large VLDL | 0.05 | 0.02 | 0.05 | 0.02 | -22.86 | < 0.0001 | 0.12 | < 0.0001 |
| Free Chol. in LDL | 0.47 | 0.12 | 0.43 | 0.13 | 65.49 | < 0.0001 | -0.38 | < 0.0001 |
| Free Chol. in Medium HDL | 0.09 | 0.02 | 0.08 | 0.02 | 58.27 | < 0.0001 | -0.31 | < 0.0001 |
| Free Chol. in Medium LDL | 0.12 | 0.03 | 0.11 | 0.03 | 55.32 | < 0.0001 | -0.32 | < 0.0001 |
| Free Chol. in Medium VLDL | 0.08 | 0.03 | 0.08 | 0.03 | 36.09 | < 0.0001 | -0.20 | < 0.0001 |
| Free Chol. in Small HDL | 0.12 | 0.02 | 0.11 | 0.02 | 30.40 | < 0.0001 | -0.17 | < 0.0001 |
| Free Chol. in Small LDL | 0.05 | 0.01 | 0.05 | 0.01 | 55.53 | < 0.0001 | -0.32 | < 0.0001 |
| Free Chol. in Small VLDL | 0.06 | 0.02 | 0.06 | 0.02 | 34.64 | < 0.0001 | -0.20 | < 0.0001 |
| Free Chol. in Very Large HDL | 0.02 | 0.01 | 0.02 | 0.01 | 67.33 | < 0.0001 | -0.35 | < 0.0001 |
| Free Chol. in Very Large VLDL | 0.02 | 0.01 | 0.03 | 0.01 | -21.66 | < 0.0001 | 0.12 | < 0.0001 |
| Free Chol. in Very Small VLDL | 0.06 | 0.01 | 0.06 | 0.02 | 35.39 | < 0.0001 | -0.20 | < 0.0001 |
| Free Chol. in VLDL | 0.30 | 0.11 | 0.29 | 0.11 | 8.02 | < 0.0001 | -0.04 | < 0.0001 |
| Free Chol./T.Lipids in Chylomicrons | 12.95 | 4.57 | 12.71 | 4.52 | 9.92 | < 0.0001 | -0.05 | < 0.0001 |
| Free Chol./T.Lipids in IDL | 17.75 | 1.08 | 17.45 | 1.27 | 44.66 | < 0.0001 | -0.27 | < 0.0001 |
| Free Chol./T.Lipids in Large HDL | 10.02 | 1.10 | 9.56 | 1.47 | 59.67 | < 0.0001 | -0.39 | < 0.0001 |
| Free Chol./T.Lipids in Large LDL | 18.56 | 1.25 | 17.94 | 1.50 | 79.07 | < 0.0001 | -0.49 | < 0.0001 |
| Free Chol./T.Lipids in Large VLDL | 13.92 | 1.28 | 14.02 | 1.19 | -15.71 | < 0.0001 | 0.08 | < 0.0001 |
| Free Chol./T.Lipids in Medium HDL | 8.34 | 0.67 | 8.04 | 0.70 | 78.49 | < 0.0001 | -0.44 | < 0.0001 |
| Free Chol./T.Lipids in Medium LDL | 19.29 | 1.85 | 18.61 | 2.05 | 61.93 | < 0.0001 | -0.36 | < 0.0001 |
| Free Chol./T.Lipids in Medium VLDL | 13.76 | 1.67 | 12.98 | 1.91 | 76.47 | < 0.0001 | -0.46 | < 0.0001 |
| Free Chol./T.Lipids in Small HDL | 9.94 | 0.51 | 9.80 | 0.52 | 47.84 | < 0.0001 | -0.26 | < 0.0001 |
| Free Chol./T.Lipids in Small LDL | 17.39 | 1.91 | 16.79 | 2.18 | 50.94 | < 0.0001 | -0.30 | < 0.0001 |
| Free Chol./T.Lipids in Small VLDL | 14.43 | 2.13 | 13.34 | 2.26 | 88.75 | < 0.0001 | -0.50 | < 0.0001 |
| Free Chol./T.Lipids in Very Large HDL | 15.10 | 3.51 | 16.32 | 4.21 | -54.52 | < 0.0001 | 0.34 | < 0.0001 |
| Free Chol./T.Lipids in Very Large VLDL | 12.02 | 1.62 | 11.57 | 1.48 | 55.10 | < 0.0001 | -0.28 | < 0.0001 |
| Free Chol./T.Lipids in Very Small VLDL | 16.06 | 0.59 | 15.87 | 0.69 | 52.30 | < 0.0001 | -0.32 | < 0.0001 |
| Glucose | 3.65 | 1.06 | 3.94 | 1.58 | -35.50 | < 0.0001 | 0.25 | < 0.0001 |
| Glucose lactate | 5.15 | 1.08 | 5.46 | 1.62 | -37.06 | < 0.0001 | 0.26 | < 0.0001 |
| Glutamine | 0.55 | 0.08 | 0.55 | 0.09 | -6.14 | < 0.0001 | 0.03 | < 0.0001 |

| | | | | | | | | |
|---|---|---|---|---|---|---|---|---|
| Glycine | 0.17 | 0.07 | 0.16 | 0.06 | 47.28 | < 0.0001 | -0.24 | < 0.0001 |
| Glycoprotein Acetyls | 0.81 | 0.12 | 0.84 | 0.12 | -42.75 | < 0.0001 | 0.24 | < 0.0001 |
| HDL Chol. | 1.33 | 0.32 | 1.21 | 0.30 | 74.98 | < 0.0001 | -0.39 | < 0.0001 |
| Histidine | 0.07 | 0.01 | 0.07 | 0.01 | 11.62 | < 0.0001 | -0.06 | < 0.0001 |
| Isoleucine | 0.05 | 0.02 | 0.05 | 0.02 | -27.52 | < 0.0001 | 0.15 | < 0.0001 |
| Lactate | 3.95 | 1.11 | 4.00 | 1.12 | -8.86 | < 0.0001 | 0.05 | < 0.0001 |
| LDL Chol. | 1.78 | 0.43 | 1.64 | 0.48 | 55.12 | < 0.0001 | -0.32 | < 0.0001 |
| Leucine | 0.10 | 0.03 | 0.11 | 0.03 | -29.56 | < 0.0001 | 0.16 | < 0.0001 |
| Linoleic Acid | 3.52 | 0.68 | 3.29 | 0.73 | 57.25 | < 0.0001 | -0.33 | < 0.0001 |
| Linoleic Acid/Total Fatty Acids | 29.11 | 3.34 | 27.58 | 3.58 | 79.65 | < 0.0001 | -0.45 | < 0.0001 |
| MUFA | 2.92 | 0.82 | 3.00 | 0.88 | -16.75 | < 0.0001 | 0.10 | < 0.0001 |
| MUFA/Total Fatty Acids | 23.73 | 2.55 | 24.70 | 2.72 | -65.88 | < 0.0001 | 0.37 | < 0.0001 |
| Omega 3 Fatty Acids | 0.53 | 0.22 | 0.53 | 0.22 | 5.40 | < 0.0001 | -0.03 | < 0.0001 |
| Omega 3/Total Fatty Acids | 4.35 | 1.52 | 4.37 | 1.52 | -2.38 | 0.0175 | 0.01 | 0.0181 |
| Omega 6 Fatty Acids | 4.56 | 0.68 | 4.35 | 0.73 | 51.85 | < 0.0001 | -0.30 | < 0.0001 |
| Omega 6/Omega 3 | 9.89 | 4.31 | 9.55 | 4.24 | 14.74 | < 0.0001 | -0.08 | < 0.0001 |
| Omega 6/Total Fatty Acids | 37.93 | 3.51 | 36.78 | 3.79 | 56.36 | < 0.0001 | -0.32 | < 0.0001 |
| Phenylalanine | 0.05 | 0.01 | 0.05 | 0.01 | -27.58 | < 0.0001 | 0.15 | < 0.0001 |
| Phosphatidylcholines | 2.12 | 0.37 | 2.00 | 0.39 | 56.62 | < 0.0001 | -0.32 | < 0.0001 |
| Phosphoglycerides | 2.31 | 0.39 | 2.20 | 0.42 | 49.96 | < 0.0001 | -0.28 | < 0.0001 |
| PL in Chylomicrons | 0.04 | 0.03 | 0.04 | 0.03 | -33.64 | < 0.0001 | 0.19 | < 0.0001 |
| PL in HDL | 1.56 | 0.31 | 1.47 | 0.30 | 55.88 | < 0.0001 | -0.30 | < 0.0001 |
| PL in IDL | 0.30 | 0.06 | 0.27 | 0.07 | 66.85 | < 0.0001 | -0.39 | < 0.0001 |
| PL in Large HDL | 0.32 | 0.15 | 0.28 | 0.13 | 67.99 | < 0.0001 | -0.34 | < 0.0001 |
| PL in Large LDL | 0.36 | 0.08 | 0.33 | 0.09 | 60.26 | < 0.0001 | -0.35 | < 0.0001 |
| PL in Large VLDL | 0.07 | 0.04 | 0.07 | 0.04 | -27.06 | < 0.0001 | 0.15 | < 0.0001 |
| PL in LDL | 0.62 | 0.14 | 0.58 | 0.15 | 52.01 | < 0.0001 | -0.30 | < 0.0001 |
| PL in Medium HDL | 0.49 | 0.10 | 0.47 | 0.10 | 39.36 | < 0.0001 | -0.22 | < 0.0001 |
| PL in Medium LDL | 0.17 | 0.04 | 0.16 | 0.04 | 37.76 | < 0.0001 | -0.22 | < 0.0001 |
| PL in Medium VLDL | 0.13 | 0.05 | 0.13 | 0.05 | 27.35 | < 0.0001 | -0.15 | < 0.0001 |
| PL in Small HDL | 0.67 | 0.09 | 0.66 | 0.10 | 14.04 | < 0.0001 | -0.08 | < 0.0001 |
| PL in Small LDL | 0.09 | 0.02 | 0.09 | 0.02 | 40.01 | < 0.0001 | -0.23 | < 0.0001 |
| PL in Small VLDL | 0.10 | 0.03 | 0.10 | 0.03 | 18.54 | < 0.0001 | -0.10 | < 0.0001 |
| PL in Very Large HDL | 0.08 | 0.04 | 0.06 | 0.04 | 68.76 | < 0.0001 | -0.34 | < 0.0001 |
| PL in Very Large VLDL | 0.04 | 0.03 | 0.04 | 0.03 | -25.81 | < 0.0001 | 0.14 | < 0.0001 |
| PL in Very Small VLDL | 0.11 | 0.03 | 0.10 | 0.03 | 20.77 | < 0.0001 | -0.12 | < 0.0001 |
| PL in VLDL | 0.48 | 0.18 | 0.49 | 0.18 | -1.85 | 0.0642 | 0.01 | 0.0658 |
| PL/T.Lipids in Chylomicrons | 15.13 | 4.32 | 16.19 | 3.77 | -50.29 | < 0.0001 | 0.25 | < 0.0001 |
| PL/T.Lipids in IDL | 23.83 | 0.88 | 24.04 | 0.98 | -40.23 | < 0.0001 | 0.24 | < 0.0001 |
| PL/T.Lipids in Large HDL | 50.40 | 2.86 | 51.80 | 3.60 | -73.31 | < 0.0001 | 0.46 | < 0.0001 |
| PL/T.Lipids in Large LDL | 22.51 | 0.78 | 22.50 | 0.86 | 1.92 | 0.0554 | -0.01 | 0.0570 |
| PL/T.Lipids in Large VLDL | 19.16 | 3.07 | 19.83 | 2.48 | -48.07 | < 0.0001 | 0.23 | < 0.0001 |
| PL/T.Lipids in Medium HDL | 47.07 | 1.23 | 47.57 | 1.30 | -69.99 | < 0.0001 | 0.40 | < 0.0001 |
| PL/T.Lipids in Medium LDL | 26.12 | 0.80 | 26.15 | 0.80 | -6.22 | < 0.0001 | 0.03 | < 0.0001 |
| PL/T.Lipids in Medium VLDL | 22.32 | 1.71 | 21.52 | 2.02 | 74.25 | < 0.0001 | -0.45 | < 0.0001 |
| PL/T.Lipids in Small HDL | 57.05 | 1.22 | 57.07 | 1.23 | -2.65 | 0.0081 | 0.01 | 0.0084 |
| PL/T.Lipids in Small LDL | 31.18 | 1.70 | 31.28 | 1.78 | -9.84 | < 0.0001 | 0.06 | < 0.0001 |
| PL/T.Lipids in Small VLDL | 23.76 | 1.99 | 22.78 | 2.09 | 87.17 | < 0.0001 | -0.49 | < 0.0001 |
| PL/T.Lipids in Very Large HDL | 44.59 | 6.55 | 43.09 | 7.59 | 37.27 | < 0.0001 | -0.22 | < 0.0001 |
| PL/T.Lipids in Very Large VLDL | 18.39 | 2.61 | 18.38 | 2.16 | 0.44 | 0.6627 | 0.00 | 0.6653 |
| PL/T.Lipids in Very Small VLDL | 29.21 | 0.86 | 29.59 | 0.94 | -75.15 | < 0.0001 | 0.44 | < 0.0001 |
| Polyunsaturated Fatty Acids | 5.09 | 0.80 | 4.88 | 0.85 | 45.93 | < 0.0001 | -0.26 | < 0.0001 |
| PUFA/MUFA | 1.82 | 0.33 | 1.70 | 0.33 | 63.72 | < 0.0001 | -0.35 | < 0.0001 |
| PUFA Total Fatty Acids | 42.28 | 3.64 | 41.15 | 3.95 | 53.23 | < 0.0001 | -0.31 | < 0.0001 |
| Pyruvate | 0.08 | 0.03 | 0.08 | 0.03 | -0.98 | 0.3288 | 0.01 | 0.3314 |
| Remnant Chol (Non HDL-LDL). | 1.59 | 0.41 | 1.47 | 0.45 | 52.27 | < 0.0001 | -0.30 | < 0.0001 |
| Saturated Fatty Acids | 4.15 | 0.95 | 4.12 | 1.03 | 5.16 | < 0.0001 | -0.03 | < 0.0001 |
| SFA Total Fatty Acids | 33.99 | 1.92 | 34.15 | 2.10 | -14.65 | < 0.0001 | 0.08 | < 0.0001 |
| Spectrometer corrected alanine | 0.36 | 0.07 | 0.38 | 0.08 | -31.87 | < 0.0001 | 0.18 | < 0.0001 |
| Sphingomyelins | 0.45 | 0.07 | 0.43 | 0.08 | 68.16 | < 0.0001 | -0.39 | < 0.0001 |
| Total Chol. | 4.70 | 0.92 | 4.31 | 1.03 | 70.64 | < 0.0001 | -0.41 | < 0.0001 |
| Total Chol. Minus HDL C | 3.37 | 0.82 | 3.10 | 0.91 | 54.58 | < 0.0001 | -0.32 | < 0.0001 |
| Total Cholines | 2.60 | 0.41 | 2.46 | 0.43 | 59.77 | < 0.0001 | -0.34 | < 0.0001 |
| Total Conc. of Lipoprotein Particles | 0.02 | 0.00 | 0.02 | 0.00 | 61.69 | < 0.0001 | -0.35 | < 0.0001 |
| Total Esterified Chol. | 3.41 | 0.66 | 3.13 | 0.74 | 72.72 | < 0.0001 | -0.43 | < 0.0001 |
| Total Fatty Acids | 12.16 | 2.38 | 12.01 | 2.57 | 11.51 | < 0.0001 | -0.07 | < 0.0001 |
| Total Free Chol. | 1.29 | 0.26 | 1.19 | 0.29 | 64.32 | < 0.0001 | -0.38 | < 0.0001 |

| | | | | | | | | |
|---|---|---|---|---|---|---|---|---|
| T.Lipids in Chylomicrons | 0.23 | 0.20 | 0.26 | 0.21 | -30.38 | < 0.0001 | 0.17 | < 0.0001 |
| T.Lipids in HDL | 3.04 | 0.63 | 2.83 | 0.60 | 63.35 | < 0.0001 | -0.34 | < 0.0001 |
| T.Lipids in IDL | 1.25 | 0.28 | 1.14 | 0.32 | 69.19 | < 0.0001 | -0.41 | < 0.0001 |
| T.Lipids in Large HDL | 0.65 | 0.31 | 0.54 | 0.27 | 72.82 | < 0.0001 | -0.36 | < 0.0001 |
| T.Lipids in Large LDL | 1.62 | 0.37 | 1.49 | 0.41 | 58.04 | < 0.0001 | -0.34 | < 0.0001 |
| T.Lipids in Large VLDL | 0.34 | 0.18 | 0.37 | 0.18 | -22.89 | < 0.0001 | 0.13 | < 0.0001 |
| T.Lipids in LDL | 2.54 | 0.59 | 2.37 | 0.65 | 50.92 | < 0.0001 | -0.29 | < 0.0001 |
| T.Lipids in Lipoprotein Particles | 9.00 | 1.63 | 8.54 | 1.80 | 46.93 | < 0.0001 | -0.27 | < 0.0001 |
| T.Lipids in Medium HDL | 1.05 | 0.22 | 0.99 | 0.22 | 46.24 | < 0.0001 | -0.25 | < 0.0001 |
| T.Lipids in Medium LDL | 0.63 | 0.16 | 0.60 | 0.18 | 36.87 | < 0.0001 | -0.21 | < 0.0001 |
| T.Lipids in Medium VLDL | 0.60 | 0.20 | 0.58 | 0.21 | 13.42 | < 0.0001 | -0.07 | < 0.0001 |
| T.Lipids in Small HDL | 1.17 | 0.16 | 1.16 | 0.16 | 15.15 | < 0.0001 | -0.08 | < 0.0001 |
| T.Lipids in Small LDL | 0.29 | 0.06 | 0.28 | 0.07 | 38.70 | < 0.0001 | -0.22 | < 0.0001 |
| T.Lipids in Small VLDL | 0.42 | 0.13 | 0.43 | 0.13 | -3.24 | 0.0012 | 0.02 | 0.0013 |
| T.Lipids in Very Large HDL | 0.17 | 0.08 | 0.14 | 0.07 | 72.47 | < 0.0001 | -0.36 | < 0.0001 |
| T.Lipids in Very Large VLDL | 0.21 | 0.13 | 0.23 | 0.14 | -29.01 | < 0.0001 | 0.16 | < 0.0001 |
| T.Lipids in Very Small VLDL | 0.36 | 0.09 | 0.35 | 0.09 | 29.60 | < 0.0001 | -0.17 | < 0.0001 |
| T.Lipids in VLDL | 2.16 | 0.85 | 2.21 | 0.87 | -10.67 | < 0.0001 | 0.06 | < 0.0001 |
| T.PL in Lipoprotein Particles | 2.96 | 0.46 | 2.80 | 0.51 | 58.54 | < 0.0001 | -0.34 | < 0.0001 |
| T.TG | 1.33 | 0.57 | 1.43 | 0.60 | -29.52 | < 0.0001 | 0.17 | < 0.0001 |
| TG in Chylomicrons | 0.13 | 0.13 | 0.16 | 0.14 | -30.25 | < 0.0001 | 0.17 | < 0.0001 |
| TG in HDL | 0.15 | 0.05 | 0.15 | 0.05 | -24.86 | < 0.0001 | 0.14 | < 0.0001 |
| TG in IDL | 0.10 | 0.03 | 0.10 | 0.03 | -21.21 | < 0.0001 | 0.12 | < 0.0001 |
| TG in Large HDL | 0.03 | 0.01 | 0.03 | 0.01 | 7.45 | < 0.0001 | -0.04 | < 0.0001 |
| TG in Large LDL | 0.10 | 0.03 | 0.10 | 0.03 | -21.45 | < 0.0001 | 0.12 | < 0.0001 |
| TG in Large VLDL | 0.17 | 0.09 | 0.19 | 0.10 | -25.66 | < 0.0001 | 0.14 | < 0.0001 |
| TG in LDL | 0.15 | 0.04 | 0.15 | 0.04 | -24.23 | < 0.0001 | 0.14 | < 0.0001 |
| TG in Medium HDL | 0.05 | 0.02 | 0.06 | 0.02 | -24.08 | < 0.0001 | 0.13 | < 0.0001 |
| TG in Medium LDL | 0.03 | 0.01 | 0.03 | 0.01 | -27.65 | < 0.0001 | 0.16 | < 0.0001 |
| TG in Medium VLDL | 0.28 | 0.12 | 0.30 | 0.12 | -18.95 | < 0.0001 | 0.11 | < 0.0001 |
| TG in Small HDL | 0.05 | 0.02 | 0.06 | 0.02 | -47.28 | < 0.0001 | 0.26 | < 0.0001 |
| TG in Small LDL | 0.02 | 0.01 | 0.02 | 0.01 | -28.13 | < 0.0001 | 0.16 | < 0.0001 |
| TG in Small VLDL | 0.16 | 0.06 | 0.17 | 0.06 | -35.01 | < 0.0001 | 0.19 | < 0.0001 |
| TG in Very Large HDL | 0.01 | 0.00 | 0.01 | 0.00 | -1.41 | 0.1576 | 0.01 | 0.1601 |
| TG in Very Large VLDL | 0.11 | 0.08 | 0.13 | 0.08 | -34.95 | < 0.0001 | 0.20 | < 0.0001 |
| TG in Very Small VLDL | 0.07 | 0.02 | 0.07 | 0.02 | -33.11 | < 0.0001 | 0.18 | < 0.0001 |
| TG in VLDL | 0.94 | 0.48 | 1.02 | 0.50 | -29.61 | < 0.0001 | 0.17 | < 0.0001 |
| TG/Phosphoglycerides ratio | 0.58 | 0.22 | 0.65 | 0.23 | -58.58 | < 0.0001 | 0.33 | < 0.0001 |
| TG/T.Lipids in Chylomicrons | 54.63 | 14.15 | 54.86 | 13.61 | -3.00 | 0.0027 | 0.02 | 0.0028 |
| TG/T.Lipids in IDL | 8.30 | 2.33 | 9.64 | 2.96 | -85.88 | < 0.0001 | 0.55 | < 0.0001 |
| TG/T.Lipids in Large HDL | 5.50 | 3.11 | 6.56 | 3.83 | -52.50 | < 0.0001 | 0.33 | < 0.0001 |
| TG/T.Lipids in Large LDL | 6.23 | 1.69 | 7.12 | 2.25 | -74.99 | < 0.0001 | 0.49 | < 0.0001 |
| TG/T.Lipids in Large VLDL | 50.60 | 5.42 | 50.85 | 5.17 | -9.07 | < 0.0001 | 0.05 | < 0.0001 |
| TG/T.Lipids in Medium HDL | 5.33 | 1.79 | 5.88 | 1.81 | -55.35 | < 0.0001 | 0.30 | < 0.0001 |
| TG/T.Lipids in Medium LDL | 5.36 | 1.47 | 6.05 | 1.85 | -70.48 | < 0.0001 | 0.45 | < 0.0001 |
| TG/T.Lipids in Medium VLDL | 47.48 | 8.05 | 51.46 | 9.11 | -81.69 | < 0.0001 | 0.48 | < 0.0001 |
| TG/T.Lipids in Small HDL | 4.54 | 1.28 | 4.99 | 1.29 | -64.83 | < 0.0001 | 0.35 | < 0.0001 |
| TG/T.Lipids in Small LDL | 5.45 | 1.71 | 6.14 | 2.04 | -63.85 | < 0.0001 | 0.39 | < 0.0001 |
| TG/T.Lipids in Small VLDL | 38.08 | 6.41 | 40.96 | 6.97 | -76.90 | < 0.0001 | 0.44 | < 0.0001 |
| TG/T.Lipids in Very Large HDL | 5.21 | 3.56 | 6.34 | 4.83 | -44.83 | < 0.0001 | 0.30 | < 0.0001 |
| TG/T.Lipids in Very Large VLDL | 52.32 | 7.39 | 54.71 | 6.96 | -62.44 | < 0.0001 | 0.33 | < 0.0001 |
| TG/T.Lipids in Very Small VLDL | 19.35 | 3.98 | 21.55 | 4.65 | -88.96 | < 0.0001 | 0.54 | < 0.0001 |
| Tyrosine | 0.06 | 0.01 | 0.07 | 0.02 | -27.76 | < 0.0001 | 0.16 | < 0.0001 |
| Valine | 0.21 | 0.04 | 0.22 | 0.04 | -32.96 | < 0.0001 | 0.18 | < 0.0001 |
| VLDL Chol. | 0.74 | 0.24 | 0.71 | 0.25 | 23.20 | < 0.0001 | -0.13 | < 0.0001 |
| 3-Hydroxybutyrate | 0.06 | 0.06 | 0.06 | 0.06 | -2.61 | 0.0091 | 0.01 | 0.0094 |

BCAA: branched-chain amino acids. CE: Cholesteryl esters. Chol.:cholesterol. DHA: docosahexaenoic acid. MUFA: monounsaturated fatty acids. PL: phospholipids. PUFA: polyunsaturated fatty acids. SFA: saturated fatty acids. T.: Total. TG: triglycerides.

**Supplementary Table S4.** Model 4. StepAIC regression model after debugging the model for metabolomic variables with high multicollinearity (VIF >5).

| Predictor | Estimate | Std Error | Z | p | OR | Lower CI | Upper CI | FDR |
|---|---|---|---|---|---|---|---|---|
| (Intercept) | -1.80 | 0.01 | -290.50 | <0.001 | 0.16 | 0.16 | 0.17 | <0.001 |
| Acetate | -0.02 | 0.01 | -3.32 | 0.0009 | 0.98 | 0.97 | 0.99 | 0.0010 |
| Acetoacetate | 0.03 | 0.01 | 3.16 | 0.0016 | 1.03 | 1.01 | 1.05 | 0.0017 |
| Acetone | 0.08 | 0.01 | 8.58 | <0.001 | 1.08 | 1.06 | 1.10 | <0.001 |
| Alanine | 0.03 | 0.01 | 4.68 | <0.001 | 1.03 | 1.02 | 1.04 | <0.001 |
| Albumin | -0.21 | 0.01 | -30.05 | <0.001 | 0.81 | 0.80 | 0.83 | <0.001 |
| Apolipoprotein B/Apolipoprotein A1 | 0.00 | 0.01 | 0.49 | 0.6235 | 1.00 | 0.99 | 1.02 | 0.6235 |
| Average Diameter for LDL Particles | -0.08 | 0.01 | -9.77 | <0.001 | 0.92 | 0.91 | 0.94 | <0.001 |
| Chol./T.Lipids in Medium LDL | -0.37 | 0.01 | -35.54 | <0.001 | 0.69 | 0.67 | 0.70 | <0.001 |
| Chol./T.Lipids in V.Large HDL | 0.09 | 0.02 | 5.75 | <0.001 | 1.10 | 1.06 | 1.14 | <0.001 |
| CE/T.Lipids in V.Large HDL | -0.08 | 0.01 | -6.04 | <0.001 | 0.92 | 0.90 | 0.95 | <0.001 |
| Citrate | 0.02 | 0.01 | 3.50 | 0.0005 | 1.02 | 1.01 | 1.04 | 0.0006 |
| Creatinine | 0.26 | 0.01 | 38.27 | <0.001 | 1.29 | 1.28 | 1.31 | <0.001 |
| Free Chol./T.Lipids in Large HDL | -0.08 | 0.01 | -6.36 | <0.001 | 0.92 | 0.90 | 0.95 | <0.001 |
| Free Chol./T.Lipids in V.Large VLDL | -0.04 | 0.01 | -4.19 | <0.001 | 0.96 | 0.94 | 0.98 | <0.001 |
| Glucose | 0.09 | 0.01 | 17.27 | <0.001 | 1.10 | 1.08 | 1.11 | <0.001 |
| Glutamine | 0.18 | 0.01 | 25.84 | <0.001 | 1.19 | 1.18 | 1.21 | <0.001 |
| Glycine | -0.18 | 0.01 | -25.99 | <0.001 | 0.83 | 0.82 | 0.84 | <0.001 |
| Glycoprotein Acetyls | 0.11 | 0.01 | 15.55 | <0.001 | 1.12 | 1.10 | 1.13 | <0.001 |
| Histidine | -0.11 | 0.01 | -15.07 | <0.001 | 0.90 | 0.89 | 0.91 | <0.001 |
| Isoleucine | -0.16 | 0.01 | -20.29 | <0.001 | 0.85 | 0.84 | 0.87 | <0.001 |
| Lactate | 0.03 | 0.01 | 4.03 | 0.0001 | 1.03 | 1.02 | 1.05 | 0.0001 |
| Omega 3/Total Fatty Acids | 0.05 | 0.01 | 5.16 | <0.001 | 1.05 | 1.03 | 1.07 | <0.001 |
| Omega 6/Omega 3 | -0.06 | 0.01 | -5.44 | <0.001 | 0.94 | 0.92 | 0.96 | <0.001 |
| Phenylalanine | 0.02 | 0.01 | 3.32 | 0.0009 | 1.02 | 1.01 | 1.04 | 0.0010 |
| PL/T.Lipids in Chylomicrons | 0.17 | 0.01 | 18.41 | <0.001 | 1.19 | 1.17 | 1.21 | <0.001 |
| PL/T.Lipids in Medium LDL | -0.16 | 0.01 | -14.18 | <0.001 | 0.85 | 0.83 | 0.87 | <0.001 |
| PL/T.Lipids in V.Large VLDL | -0.06 | 0.01 | -5.79 | <0.001 | 0.94 | 0.92 | 0.96 | <0.001 |
| Pyruvate | -0.03 | 0.01 | -4.42 | <0.001 | 0.97 | 0.95 | 0.98 | <0.001 |
| TG in Large HDL | -0.27 | 0.01 | -25.41 | <0.001 | 0.76 | 0.75 | 0.78 | <0.001 |
| TG/T.Lipids in Chylomicrons | -0.02 | 0.01 | -1.91 | 0.0564 | 0.98 | 0.97 | 1.00 | 0.0581 |
| TG/T.Lipids in V.Large HDL | -0.08 | 0.01 | -7.72 | <0.001 | 0.93 | 0.91 | 0.94 | <0.001 |
| Tyrosine | 0.14 | 0.01 | 18.89 | <0.001 | 1.15 | 1.13 | 1.17 | <0.001 |

Chol.: cholesterol. CE: cholesteryl esters. PL: phospsholipids. T.: Total. TG: triglycerides. V.: Very. Overall model performance: $R^2_{McFadden}$: 0.080. $R^2_{Nagelkerke}$: 0.116. AIC: 202624.18. Chi-cuadrado global:17555.16, df: 32, p = 0.

**Supplementary Table S5.** Dietary data attending to the presence or not of a cardiovascular event during the study period.

| | NO CVD (n=111175) | | CVD (n=15476) | | t | p value | Cohen d | FDR |
|---|---|---|---|---|---|---|---|---|
| **Variable** | **Mean** | **sd** | **Mean** | **sd** | | | | |
| Energy (MJ/d) | 8625.64 | 2162.87 | 8770.08 | 2224.43 | -7.59 | < 0.0001 | 0.07 | < 0.0001 |
| Added sugars and preserves | 8.47 | 12.20 | 9.92 | 13.88 | -12.40 | < 0.0001 | 0.12 | < 0.0001 |
| Allium vegetables | 11.88 | 17.31 | 11.72 | 17.50 | 1.06 | 0.2904 | -0.01 | 0.3171 |
| Animal fat spread lower fat | 0.85 | 3.24 | 1.00 | 3.57 | -4.87 | < 0.0001 | 0.05 | < 0.0001 |
| Animal fat spread normal | 5.13 | 8.19 | 5.23 | 8.81 | -1.33 | 0.1844 | 0.01 | 0.2111 |
| Apples and pears | 61.09 | 71.62 | 62.55 | 73.52 | -2.32 | 0.0203 | 0.02 | 0.0268 |
| Beef | 22.13 | 32.23 | 24.11 | 33.71 | -6.88 | < 0.0001 | 0.06 | < 0.0001 |
| Beer and cider | 135.74 | 331.60 | 175.81 | 385.89 | -12.30 | < 0.0001 | 0.12 | < 0.0001 |
| Berries | 8.78 | 15.53 | 7.96 | 14.87 | 6.40 | < 0.0001 | -0.05 | < 0.0001 |
| Biscuit cereal | 5.17 | 11.73 | 6.23 | 13.09 | -9.55 | < 0.0001 | 0.09 | < 0.0001 |
| Biscuits | 12.01 | 15.81 | 13.21 | 16.96 | -8.27 | < 0.0001 | 0.07 | < 0.0001 |
| Bran cereal | 3.99 | 10.35 | 4.07 | 10.64 | -0.82 | 0.4099 | 0.01 | 0.4425 |
| Breaded battered chicken | 3.17 | 12.86 | 3.61 | 14.28 | -3.66 | 0.0003 | 0.03 | 0.0005 |
| Breaded battered fish | 5.78 | 19.15 | 6.24 | 19.92 | -2.68 | 0.0075 | 0.02 | 0.0108 |
| Chocolate confectionery | 9.30 | 15.96 | 8.43 | 15.67 | 6.46 | < 0.0001 | -0.05 | < 0.0001 |
| Citrus | 32.75 | 48.46 | 33.97 | 50.16 | -2.85 | 0.0044 | 0.03 | 0.0065 |
| Coffee caffeinated | 240.77 | 253.20 | 245.92 | 260.88 | -2.31 | 0.0209 | 0.02 | 0.0272 |
| Coffee decaffeinated | 53.76 | 141.29 | 63.28 | 156.77 | -7.16 | < 0.0001 | 0.07 | < 0.0001 |
| Cream | 1.32 | 3.97 | 1.15 | 3.77 | 5.43 | < 0.0001 | -0.04 | < 0.0001 |
| Dried fruit | 7.43 | 17.31 | 7.19 | 17.94 | 1.52 | 0.1291 | -0.01 | 0.1496 |
| Egg and egg dishes | 21.17 | 32.88 | 22.73 | 33.91 | -5.40 | < 0.0001 | 0.05 | < 0.0001 |
| Fortified wine | 1.06 | 6.23 | 1.06 | 6.15 | 0.05 | 0.9573 | 0.00 | 0.9675 |
| Fried roast potatoes | 30.21 | 47.82 | 32.88 | 50.38 | -6.22 | < 0.0001 | 0.06 | < 0.0001 |
| Fruit juice | 107.74 | 128.55 | 107.99 | 130.85 | -0.22 | 0.8235 | 0.00 | 0.8412 |
| Full fat yogurt | 10.30 | 24.76 | 8.28 | 22.33 | 10.41 | < 0.0001 | -0.08 | < 0.0001 |
| Grain dishes added fat | 7.61 | 15.05 | 8.20 | 15.54 | -4.49 | < 0.0001 | 0.04 | < 0.0001 |
| Green leafy cabbages | 29.15 | 39.08 | 30.52 | 41.39 | -3.89 | 0.0001 | 0.03 | 0.0002 |
| High fat cheese | 14.88 | 16.50 | 13.61 | 15.98 | 9.21 | < 0.0001 | -0.08 | < 0.0001 |
| Lamb | 6.52 | 18.54 | 7.13 | 19.89 | -3.64 | 0.0003 | 0.03 | 0.0005 |
| Legumes and pulses | 13.23 | 24.51 | 13.66 | 25.12 | -2.03 | 0.0423 | 0.02 | 0.0522 |
| Low fat yogurt | 33.61 | 48.16 | 33.31 | 48.02 | 0.73 | 0.4660 | -0.01 | 0.4974 |
| Sweetened beverages | 69.01 | 175.92 | 75.59 | 188.52 | -4.10 | < 0.0001 | 0.04 | < 0.0001 |
| Mashed potatoes | 17.07 | 34.88 | 19.68 | 38.00 | -8.09 | < 0.0001 | 0.07 | < 0.0001 |
| Meat substitutes/soy | 0.66 | 5.49 | 0.45 | 4.79 | 5.16 | < 0.0001 | -0.04 | < 0.0001 |
| Meat substitutes/vegetarian | 3.67 | 16.71 | 2.70 | 14.51 | 7.62 | < 0.0001 | -0.06 | < 0.0001 |
| Medium and low-fat cheese | 2.89 | 8.31 | 2.91 | 8.21 | -0.31 | 0.7569 | 0.00 | 0.7816 |
| Milk based drinks | 32.27 | 77.81 | 35.96 | 82.35 | -5.25 | < 0.0001 | 0.05 | < 0.0001 |
| Milk dairy desserts | 23.48 | 37.56 | 26.42 | 40.98 | -8.43 | < 0.0001 | 0.08 | < 0.0001 |
| Mixed bread | 23.82 | 32.32 | 24.73 | 34.07 | -3.12 | 0.0018 | 0.03 | 0.0028 |
| Muesli | 11.40 | 21.87 | 9.84 | 20.43 | 8.83 | < 0.0001 | -0.07 | < 0.0001 |
| Nut based spreads | 0.43 | 1.85 | 0.32 | 1.61 | 7.75 | < 0.0001 | -0.06 | < 0.0001 |
| Oat cereal non sugar | 34.24 | 62.98 | 35.71 | 65.49 | -2.62 | 0.0089 | 0.02 | 0.0124 |
| Oat cereal sugar | 2.52 | 9.60 | 2.25 | 9.07 | 3.44 | 0.0006 | -0.03 | 0.0010 |
| Oily fish | 11.59 | 22.47 | 12.10 | 23.77 | -2.51 | 0.0121 | 0.02 | 0.0167 |
| Olive oil drizzling dunking | 0.32 | 1.16 | 0.26 | 1.05 | 6.47 | < 0.0001 | -0.05 | < 0.0001 |
| Other bread | 9.20 | 21.86 | 8.77 | 22.35 | 2.21 | 0.0272 | -0.02 | 0.0341 |
| Other cereal sugar | 4.65 | 9.54 | 4.99 | 9.84 | -4.09 | < 0.0001 | 0.04 | < 0.0001 |
| Other desserts and cakes | 32.49 | 34.49 | 31.50 | 34.47 | 3.35 | 0.0008 | -0.03 | 0.0013 |
| Other fruit | 92.62 | 84.05 | 93.49 | 85.54 | -1.18 | 0.2362 | 0.01 | 0.2640 |
| Other meat offal | 2.00 | 9.38 | 2.09 | 9.54 | -1.10 | 0.2701 | 0.01 | 0.2984 |
| Other sweets | 2.61 | 9.63 | 2.80 | 10.12 | -2.21 | 0.0273 | 0.02 | 0.0341 |
| Other vegetables | 58.43 | 57.34 | 55.82 | 55.84 | 5.42 | < 0.0001 | -0.05 | < 0.0001 |
| Peas and sweetcorn | 8.45 | 14.23 | 9.15 | 14.81 | -5.57 | < 0.0001 | 0.05 | < 0.0001 |
| Pizza | 11.18 | 42.60 | 8.99 | 37.63 | 6.66 | < 0.0001 | -0.05 | < 0.0001 |
| Plant based spread lower fat | 2.18 | 4.81 | 3.02 | 5.75 | -17.50 | < 0.0001 | 0.17 | < 0.0001 |
| Plant based spread normal | 3.01 | 5.58 | 3.55 | 6.08 | -10.57 | < 0.0001 | 0.10 | < 0.0001 |
| Pork | 8.29 | 20.67 | 9.67 | 22.62 | -7.15 | < 0.0001 | 0.07 | < 0.0001 |
| Potatoes sweet potatoe | 47.87 | 59.67 | 53.84 | 65.08 | -10.80 | < 0.0001 | 0.10 | < 0.0001 |
| Poultry | 30.81 | 39.67 | 29.58 | 39.67 | 3.60 | 0.0003 | -0.03 | 0.0005 |
| Processed meat | 18.00 | 24.49 | 20.72 | 26.88 | -11.92 | < 0.0001 | 0.11 | < 0.0001 |
| Raw salad | 21.02 | 26.95 | 18.63 | 25.67 | 10.75 | < 0.0001 | -0.09 | < 0.0001 |
| Red wine | 66.19 | 114.84 | 69.30 | 122.90 | -2.97 | 0.0030 | 0.03 | 0.0045 |
| Rice oat milk | 0.85 | 11.03 | 0.64 | 9.67 | 2.39 | 0.0169 | -0.02 | 0.0229 |
| Root vegetables | 20.86 | 25.39 | 21.59 | 26.09 | -3.28 | 0.0010 | 0.03 | 0.0016 |
| Salted nuts and seeds | 2.63 | 8.70 | 2.46 | 8.25 | 2.36 | 0.0183 | -0.02 | 0.0245 |
| Samosa pakora | 0.88 | 5.48 | 0.80 | 5.17 | 1.79 | 0.0742 | -0.01 | 0.0892 |
| Sauces condiments high-fat | 13.59 | 15.54 | 12.72 | 15.53 | 6.54 | < 0.0001 | -0.06 | < 0.0001 |
| Sauces condiments low-fat | 9.45 | 15.95 | 8.22 | 14.80 | 9.60 | < 0.0001 | -0.08 | < 0.0001 |
| Savoury crackers | 4.91 | 9.33 | 4.88 | 9.23 | 0.43 | 0.6676 | 0.00 | 0.6969 |

| | | | | | | | | |
|---|---|---|---|---|---|---|---|---|
| Savoury snacks | 9.08 | 14.11 | 8.69 | 14.06 | 3.20 | 0.0014 | -0.03 | 0.0022 |
| Semi skimmed milk | 138.17 | 130.12 | 141.12 | 131.48 | -2.62 | 0.0088 | 0.02 | 0.0124 |
| Shellfish | 3.27 | 10.03 | 3.03 | 10.14 | 2.77 | 0.0056 | -0.02 | 0.0082 |
| Skimmed milk | 45.95 | 96.79 | 49.05 | 100.71 | -3.60 | 0.0003 | 0.03 | 0.0005 |
| Soups | 38.36 | 62.57 | 40.98 | 66.39 | -4.64 | < 0.0001 | 0.04 | < 0.0001 |
| Soy desserts and yogurt | 0.69 | 6.85 | 0.69 | 6.78 | -0.04 | 0.9681 | 0.00 | 0.9681 |
| Soy milk | 7.93 | 38.64 | 6.65 | 35.67 | 4.14 | < 0.0001 | -0.03 | < 0.0001 |
| Spirits | 3.37 | 10.28 | 4.26 | 12.09 | -8.70 | < 0.0001 | 0.08 | < 0.0001 |
| Stewed fruit | 13.15 | 28.72 | 13.72 | 29.79 | -2.25 | 0.0247 | 0.02 | 0.0317 |
| Sugar sweetened drinks | 86.20 | 151.56 | 91.17 | 162.83 | -3.58 | 0.0003 | 0.03 | 0.0005 |
| Sushi | 1.19 | 14.04 | 0.82 | 12.30 | 3.47 | 0.0005 | -0.03 | 0.0008 |
| Tea | 420.45 | 333.87 | 432.45 | 339.90 | -4.12 | < 0.0001 | 0.04 | < 0.0001 |
| Tea decaffeinated | 83.07 | 178.51 | 68.70 | 167.76 | 9.91 | < 0.0001 | -0.08 | < 0.0001 |
| Tomatoes | 39.78 | 46.55 | 37.73 | 45.47 | 5.25 | < 0.0001 | -0.04 | < 0.0001 |
| Unsalted nuts and seeds | 4.55 | 9.82 | 4.10 | 9.37 | 5.55 | < 0.0001 | -0.05 | < 0.0001 |
| Vegetable dips | 1.31 | 4.04 | 0.92 | 3.49 | 12.60 | < 0.0001 | -0.10 | < 0.0001 |
| Vegetable side dishes | 4.58 | 14.28 | 4.38 | 14.11 | 1.68 | 0.0929 | -0.01 | 0.1103 |
| Water | 516.29 | 377.44 | 472.68 | 354.10 | 14.24 | < 0.0001 | -0.12 | < 0.0001 |
| White bread | 27.65 | 38.32 | 32.62 | 43.38 | -13.55 | < 0.0001 | 0.13 | < 0.0001 |
| White fish and tinned tuna | 12.05 | 23.02 | 11.73 | 23.36 | 1.59 | 0.1128 | -0.01 | 0.1323 |
| White pasta and rice | 44.90 | 57.76 | 35.67 | 52.16 | 20.34 | < 0.0001 | -0.16 | < 0.0001 |
| White wine | 46.19 | 99.08 | 37.84 | 92.77 | 10.39 | < 0.0001 | -0.08 | < 0.0001 |
| Whole milk | 13.31 | 55.79 | 12.71 | 54.41 | 1.28 | 0.2017 | -0.01 | 0.2281 |
| Whole meal bread | 24.79 | 35.32 | 26.11 | 37.07 | -4.19 | < 0.0001 | 0.04 | < 0.0001 |
| Whole meal pasta | 8.69 | 27.27 | 7.25 | 24.87 | 6.67 | < 0.0001 | -0.05 | < 0.0001 |

**Supplementary Table S6.** Model 4. StepAIC regression model after debugging the model for dietary variables with high multicollinearity (VIF >5) (only significant variables are shown).

| Predictor | Estimate | Std Error | Z | p | Odds Ratio | Lower CI | Upper CI | FDR |
|---|---|---|---|---|---|---|---|---|
| (Intercept) | -2.23 | 0.05 | -46.95 | <0.0001 | 0.11 | 0.10 | 0.12 | <0.0001 |
| Added sugars and preserves | 0.01 | 0.00 | 8.31 | <0.0001 | 1.01 | 1.00 | 1.01 | <0.0001 |
| Allium vegetables | 0.00 | 0.00 | 1.85 | 0.0650 | 1.00 | 1.00 | 1.00 | 0.1088 |
| Animal fat spread lower fat | 0.01 | 0.00 | 5.15 | <0.0001 | 1.01 | 1.01 | 1.02 | <0.0001 |
| Animal fat spread normal | 0.01 | 0.00 | 5.52 | <0.0001 | 1.01 | 1.00 | 1.01 | <0.0001 |
| Apples and pears | 0.00 | 0.00 | 3.15 | 0.0017 | 1.00 | 1.00 | 1.00 | 0.0035 |
| Beef | 0.00 | 0.00 | 3.67 | 0.0002 | 1.00 | 1.00 | 1.00 | 0.0006 |
| Beer and cider | 0.00 | 0.00 | 8.24 | <0.0001 | 1.00 | 1.00 | 1.00 | <0.0001 |
| Berries | 0.00 | 0.00 | -1.80 | 0.0716 | 1.00 | 1.00 | 1.00 | 0.1140 |
| Biscuit cereal | 0.01 | 0.00 | 7.60 | <0.0001 | 1.01 | 1.00 | 1.01 | <0.0001 |
| Biscuits | 0.00 | 0.00 | 4.30 | <0.0001 | 1.00 | 1.00 | 1.00 | 0.0001 |
| Bran cereal | 0.00 | 0.00 | 2.25 | 0.0245 | 1.00 | 1.00 | 1.00 | 0.0435 |
| Breaded battered chicken | 0.00 | 0.00 | 2.79 | 0.0053 | 1.00 | 1.00 | 1.00 | 0.0106 |
| Breaded battered fish | 0.00 | 0.00 | 0.30 | 0.7631 | 1.00 | 1.00 | 1.00 | 0.8341 |
| Chocolate confectionery | 0.00 | 0.00 | -7.11 | <0.0001 | 1.00 | 0.99 | 1.00 | <0.0001 |
| Citrus | 0.00 | 0.00 | 2.78 | 0.0054 | 1.00 | 1.00 | 1.00 | 0.0106 |
| Coffee caffeinated | 0.00 | 0.00 | 0.41 | 0.6782 | 1.00 | 1.00 | 1.00 | 0.7657 |
| Coffee decaffeinated | 0.00 | 0.00 | 6.85 | <0.0001 | 1.00 | 1.00 | 1.00 | <0.0001 |
| Cream | 0.00 | 0.00 | -2.06 | 0.0396 | 1.00 | 0.99 | 1.00 | 0.0677 |
| Dried fruit | 0.00 | 0.00 | 1.27 | 0.2045 | 1.00 | 1.00 | 1.00 | 0.2826 |
| Egg and egg dishes | 0.00 | 0.00 | 3.58 | 0.0003 | 1.00 | 1.00 | 1.00 | 0.0008 |
| Fortified wine | 0.00 | 0.00 | -0.21 | 0.8363 | 1.00 | 1.00 | 1.00 | 0.8856 |
| Fried roast potatoes | 0.00 | 0.00 | 0.97 | 0.3305 | 1.00 | 1.00 | 1.00 | 0.4315 |
| Fruit juice | 0.00 | 0.00 | 1.77 | 0.0764 | 1.00 | 1.00 | 1.00 | 0.1197 |
| Full fat yogurt | 0.00 | 0.00 | -6.20 | <0.0001 | 1.00 | 1.00 | 1.00 | <0.0001 |
| Grain dishes added fat | 0.00 | 0.00 | 0.41 | 0.6842 | 1.00 | 1.00 | 1.00 | 0.7657 |
| Green leafy cabbages | 0.00 | 0.00 | 1.28 | 0.1991 | 1.00 | 1.00 | 1.00 | 0.2794 |
| High fat cheese | 0.00 | 0.00 | -6.29 | <0.0001 | 1.00 | 1.00 | 1.00 | <0.0001 |
| Lamb | 0.00 | 0.00 | 2.73 | 0.0064 | 1.00 | 1.00 | 1.00 | 0.0120 |
| Legumes and pulses | 0.00 | 0.00 | -0.66 | 0.5062 | 1.00 | 1.00 | 1.00 | 0.6101 |
| Low fat yogurt | 0.00 | 0.00 | -1.35 | 0.1782 | 1.00 | 1.00 | 1.00 | 0.2578 |
| Low non sugar sugar sweetened beverages | 0.00 | 0.00 | 2.77 | 0.0056 | 1.00 | 1.00 | 1.00 | 0.0108 |
| Mashed potatoes | 0.00 | 0.00 | 2.86 | 0.0043 | 1.00 | 1.00 | 1.00 | 0.0088 |
| Meat substitutes soy | 0.00 | 0.00 | -1.14 | 0.2542 | 1.00 | 0.99 | 1.00 | 0.3366 |
| Meat substitutes vegetarian | 0.00 | 0.00 | -3.21 | 0.0013 | 1.00 | 1.00 | 1.00 | 0.0029 |
| Medium and low fat cheese | 0.00 | 0.00 | -0.11 | 0.9130 | 1.00 | 1.00 | 1.00 | 0.9431 |
| Milk based and powdered drinks | 0.00 | 0.00 | 5.67 | <0.0001 | 1.00 | 1.00 | 1.00 | <0.0001 |
| Milk dairy desserts | 0.00 | 0.00 | 5.32 | <0.0001 | 1.00 | 1.00 | 1.00 | <0.0001 |
| Mixed bread 50 50 brown and seeded | 0.00 | 0.00 | -0.22 | 0.8296 | 1.00 | 1.00 | 1.00 | 0.8856 |
| Muesli | 0.00 | 0.00 | -0.51 | 0.6102 | 1.00 | 1.00 | 1.00 | 0.7081 |
| Nut based spreads | -0.02 | 0.01 | -4.18 | <0.0001 | 0.98 | 0.97 | 0.99 | 0.0001 |
| Oat cereal non sugar | 0.00 | 0.00 | 4.57 | <0.0001 | 1.00 | 1.00 | 1.00 | <0.0001 |
| Oat cereal sugar | 0.00 | 0.00 | -0.16 | 0.8712 | 1.00 | 1.00 | 1.00 | 0.9099 |
| Oily fish | 0.00 | 0.00 | 4.83 | <0.0001 | 1.00 | 1.00 | 1.00 | <0.0001 |

| | | | | | | | | |
|---|---|---|---|---|---|---|---|---|
| Olive oil drizzling dunking | 0.00 | 0.01 | -0.43 | 0.6703 | 1.00 | 0.98 | 1.01 | 0.7657 |
| Other bread | 0.00 | 0.00 | 0.37 | 0.7104 | 1.00 | 1.00 | 1.00 | 0.7856 |
| Other cereal sugar | 0.00 | 0.00 | 3.98 | 0.0001 | 1.00 | 1.00 | 1.01 | 0.0002 |
| Other desserts and cakes and pastries | 0.00 | 0.00 | -3.95 | 0.0001 | 1.00 | 1.00 | 1.00 | 0.0002 |
| Other fruit | 0.00 | 0.00 | 3.93 | 0.0001 | 1.00 | 1.00 | 1.00 | 0.0002 |
| Other meat offal | 0.00 | 0.00 | 0.79 | 0.4289 | 1.00 | 1.00 | 1.00 | 0.5305 |
| Other sweets | 0.00 | 0.00 | 1.16 | 0.2468 | 1.00 | 1.00 | 1.00 | 0.3315 |
| Other vegetables | 0.00 | 0.00 | 0.20 | 0.8385 | 1.00 | 1.00 | 1.00 | 0.8856 |
| Peas and sweetcorn | 0.00 | 0.00 | 1.42 | 0.1568 | 1.00 | 1.00 | 1.00 | 0.2339 |
| Pizza | 0.00 | 0.00 | -4.99 | <0.0001 | 1.00 | 1.00 | 1.00 | <0.0001 |
| Plant based spread lower fat | 0.03 | 0.00 | 13.32 | <0.0001 | 1.03 | 1.02 | 1.03 | <0.0001 |
| Plant based spread normal | 0.02 | 0.00 | 8.82 | <0.0001 | 1.02 | 1.01 | 1.02 | <0.0001 |
| Pork | 0.00 | 0.00 | 4.15 | <0.0001 | 1.00 | 1.00 | 1.00 | 0.0001 |
| Potatoes sweet potatoe | 0.00 | 0.00 | 5.27 | <0.0001 | 1.00 | 1.00 | 1.00 | <0.0001 |
| Poultry | 0.00 | 0.00 | -1.69 | 0.0914 | 1.00 | 1.00 | 1.00 | 0.1409 |
| Processed meat | 0.00 | 0.00 | 6.46 | <0.0001 | 1.00 | 1.00 | 1.00 | <0.0001 |
| Raw salad | 0.00 | 0.00 | -3.62 | 0.0003 | 1.00 | 1.00 | 1.00 | 0.0007 |
| Red wine | 0.00 | 0.00 | 5.85 | <0.0001 | 1.00 | 1.00 | 1.00 | <0.0001 |
| Rice oat milk | 0.00 | 0.00 | -1.24 | 0.2154 | 1.00 | 1.00 | 1.00 | 0.2935 |
| Root vegetables | 0.00 | 0.00 | -1.57 | 0.1159 | 1.00 | 1.00 | 1.00 | 0.1757 |
| Salted nuts and seeds | 0.00 | 0.00 | -1.84 | 0.0659 | 1.00 | 1.00 | 1.00 | 0.1088 |
| Samosa pakora | 0.00 | 0.00 | 0.01 | 0.9924 | 1.00 | 1.00 | 1.00 | 0.9924 |
| Sauces condiments high fat | 0.00 | 0.00 | -6.82 | <0.0001 | 1.00 | 0.99 | 1.00 | <0.0001 |
| Sauces condiments low fat | 0.00 | 0.00 | -2.43 | 0.0151 | 1.00 | 1.00 | 1.00 | 0.0274 |
| Savoury crackers | 0.00 | 0.00 | 3.09 | 0.0020 | 1.00 | 1.00 | 1.01 | 0.0042 |
| Savoury snacks | 0.00 | 0.00 | -5.65 | <0.0001 | 1.00 | 1.00 | 1.00 | <0.0001 |
| Semi skimmed milk | 0.00 | 0.00 | -3.81 | 0.0001 | 1.00 | 1.00 | 1.00 | 0.0004 |
| Shellfish | 0.00 | 0.00 | 0.74 | 0.4565 | 1.00 | 1.00 | 1.00 | 0.5573 |
| Skimmed milk | 0.00 | 0.00 | -0.51 | 0.6089 | 1.00 | 1.00 | 1.00 | 0.7081 |
| Soups | 0.00 | 0.00 | 5.64 | <0.0001 | 1.00 | 1.00 | 1.00 | <0.0001 |
| Soy desserts and yogurt | 0.00 | 0.00 | 0.94 | 0.3466 | 1.00 | 1.00 | 1.00 | 0.4402 |
| Soy milk | 0.00 | 0.00 | -2.22 | 0.0265 | 1.00 | 1.00 | 1.00 | 0.0462 |
| Spirits | 0.01 | 0.00 | 7.78 | <0.0001 | 1.01 | 1.00 | 1.01 | <0.0001 |
| Stewed fruit | 0.00 | 0.00 | 2.66 | 0.0079 | 1.00 | 1.00 | 1.00 | 0.0145 |
| Sugar sweetened drinks | 0.00 | 0.00 | 1.37 | 0.1709 | 1.00 | 1.00 | 1.00 | 0.2510 |
| Sushi | 0.00 | 0.00 | -1.33 | 0.1837 | 1.00 | 1.00 | 1.00 | 0.2616 |
| Tea | 0.00 | 0.00 | 0.52 | 0.6045 | 1.00 | 1.00 | 1.00 | 0.7081 |
| Tea decaffeinated | 0.00 | 0.00 | -5.49 | <0.0001 | 1.00 | 1.00 | 1.00 | <0.0001 |
| Tomatoes | 0.00 | 0.00 | -0.96 | 0.3365 | 1.00 | 1.00 | 1.00 | 0.4333 |
| Unsalted nuts and seeds | 0.00 | 0.00 | -1.80 | 0.0712 | 1.00 | 1.00 | 1.00 | 0.1140 |
| Vegetable dips | -0.01 | 0.00 | -4.49 | <0.0001 | 0.99 | 0.98 | 0.99 | <0.0001 |
| Vegetable side dishes | 0.00 | 0.00 | -0.02 | 0.9857 | 1.00 | 1.00 | 1.00 | 0.9924 |
| Water | 0.00 | 0.00 | -6.16 | <0.0001 | 1.00 | 1.00 | 1.00 | <0.0001 |
| White bread | 0.00 | 0.00 | 3.31 | 0.0009 | 1.00 | 1.00 | 1.00 | 0.0021 |
| White fish and tinned tuna | 0.00 | 0.00 | 0.02 | 0.9827 | 1.00 | 1.00 | 1.00 | 0.9924 |
| White pasta and rice | 0.00 | 0.00 | -11.56 | <0.0001 | 1.00 | 1.00 | 1.00 | <0.0001 |
| White wine | 0.00 | 0.00 | -4.84 | <0.0001 | 1.00 | 1.00 | 1.00 | <0.0001 |
| Whole milk | 0.00 | 0.00 | -4.09 | <0.0001 | 1.00 | 1.00 | 1.00 | 0.0001 |
| Wholemeal bread | 0.00 | 0.00 | -0.79 | 0.4284 | 1.00 | 1.00 | 1.00 | 0.5305 |
| Wholemeal pasta | 0.00 | 0.00 | -3.42 | 0.0006 | 1.00 | 1.00 | 1.00 | 0.0014 |

Overall model performance: $R^2_{McFadden}$: 0.027 $R^2_{Nagelkerke}$: 0.038. AIC: 91702.72. Chi-cuadrado global: 2529.68, df: 93, p = 0.

**Supplementary Table S7.** Physical measures data attending to the presence or not of a cardiovascular event during the study period.

| | No CVD (n=380337) | | CVD (n=65770) | | t | p value | Cohen d | FDR |
|---|---|---|---|---|---|---|---|---|
| **Variable** | **Mean** | **sd** | **Mean** | **sd** | | | | |
| Age at recruitment | 55.73 | 8.09 | 60.75 | 6.72 | -186.93 | < 0.0001 | 0.64 | < 0.0001 |
| Arm fat free mass-left | 2.88 | 0.83 | 3.17 | 0.84 | -88.44 | < 0.0001 | 0.35 | < 0.0001 |
| Arm fat free mass-right | 2.85 | 0.81 | 3.13 | 0.83 | -85.74 | < 0.0001 | 0.34 | < 0.0001 |
| Arm fat mass-left | 1.30 | 0.70 | 1.43 | 0.79 | -41.36 | < 0.0001 | 0.18 | < 0.0001 |
| Arm fat mass-right | 1.22 | 0.62 | 1.34 | 0.70 | -42.31 | < 0.0001 | 0.18 | < 0.0001 |
| Arm fat percentage-left | 30.47 | 10.23 | 30.20 | 10.49 | 6.58 | < 0.0001 | -0.03 | < 0.0001 |
| Arm fat percentage-right | 29.57 | 10.15 | 29.33 | 10.32 | 5.82 | < 0.0001 | -0.02 | < 0.0001 |
| Arm predicted mass-left | 2.70 | 0.79 | 2.98 | 0.80 | -88.85 | < 0.0001 | 0.35 | < 0.0001 |
| Arm predicted mass-right | 2.67 | 0.78 | 2.93 | 0.79 | -86.26 | < 0.0001 | 0.34 | < 0.0001 |
| Basal metabolic rate | 6545.19 | 1346.36 | 6999.67 | 1396.52 | -83.38 | < 0.0001 | 0.34 | < 0.0001 |
| Body fat percentage | 31.43 | 8.56 | 31.56 | 8.48 | -3.99 | 0.0001 | 0.02 | 0.0001 |
| BMI | 27.16 | 4.69 | 28.88 | 5.14 | -87.34 | < 0.0001 | 0.36 | < 0.0001 |
| cIMT | 683.78 | 124.86 | 732.59 | 140.56 | -23.60 | < 0.0001 | 0.39 | < 0.0001 |
| FEV1 | 2.74 | 0.78 | 2.58 | 0.78 | 48.88 | < 0.0001 | -0.20 | < 0.0001 |
| FVC | 3.61 | 1.03 | 3.47 | 1.01 | 35.05 | < 0.0001 | -0.14 | < 0.0001 |
| Heel BMD | 0.54 | 0.14 | 0.55 | 0.15 | -2.46 | 0.0141 | 0.01 | 0.0144 |
| Hip circumference | 103.07 | 9.09 | 105.14 | 9.89 | -54.65 | < 0.0001 | 0.22 | < 0.0001 |
| Impedance of arm-left | 335.59 | 56.60 | 314.29 | 54.68 | 98.67 | < 0.0001 | -0.38 | < 0.0001 |
| Impedance of arm-right | 328.73 | 55.22 | 308.35 | 53.25 | 96.95 | < 0.0001 | -0.37 | < 0.0001 |
| Impedance of leg-left | 249.58 | 35.11 | 238.00 | 37.26 | 79.87 | < 0.0001 | -0.33 | < 0.0001 |
| Impedance of leg-right | 248.79 | 35.47 | 237.16 | 37.38 | 79.93 | < 0.0001 | -0.33 | < 0.0001 |
| Impedance of whole body | 605.50 | 88.16 | 571.04 | 87.69 | 100.04 | < 0.0001 | -0.39 | < 0.0001 |
| Leg fat free mass-left | 8.76 | 1.98 | 9.44 | 2.09 | -84.04 | < 0.0001 | 0.34 | < 0.0001 |
| Leg fat free mass-right | 8.87 | 2.00 | 9.52 | 2.09 | -80.36 | < 0.0001 | 0.33 | < 0.0001 |
| Leg fat mass-left | 4.24 | 1.84 | 4.28 | 2.03 | -5.60 | < 0.0001 | 0.02 | < 0.0001 |
| Leg fat mass-right | 4.30 | 1.87 | 4.36 | 2.07 | -7.73 | < 0.0001 | 0.03 | < 0.0001 |
| Leg fat percentage-left | 32.21 | 10.61 | 30.58 | 10.75 | 38.60 | < 0.0001 | -0.15 | < 0.0001 |
| Leg fat percentage-right | 32.28 | 10.66 | 30.81 | 10.79 | 34.70 | < 0.0001 | -0.14 | < 0.0001 |
| Leg predicted mass-left | 8.29 | 1.88 | 8.94 | 1.98 | -84.78 | < 0.0001 | 0.34 | < 0.0001 |
| Leg predicted mass-right | 8.39 | 1.90 | 9.02 | 1.99 | -81.13 | < 0.0001 | 0.33 | < 0.0001 |
| Arterial Stiffness Index | 9.22 | 3.89 | 9.96 | 4.75 | -23.77 | < 0.0001 | 0.18 | < 0.0001 |
| Seated height | 136.87 | 7.18 | 137.45 | 7.13 | -21.11 | < 0.0001 | 0.08 | < 0.0001 |
| Sitting height | 89.09 | 4.86 | 89.42 | 4.98 | -17.29 | < 0.0001 | 0.07 | < 0.0001 |
| Standing height | 168.26 | 9.27 | 169.39 | 9.28 | -31.38 | < 0.0001 | 0.12 | < 0.0001 |
| Trunk fat free mass | 29.29 | 5.93 | 31.21 | 6.00 | -81.67 | < 0.0001 | 0.32 | < 0.0001 |
| Trunk fat mass | 13.47 | 5.08 | 15.16 | 5.43 | -80.06 | < 0.0001 | 0.33 | < 0.0001 |
| Trunk fat percentage | 30.98 | 8.03 | 32.19 | 7.78 | -39.35 | < 0.0001 | 0.15 | < 0.0001 |
| Trunk predicted mass | 28.08 | 5.76 | 29.95 | 5.81 | -82.24 | < 0.0001 | 0.33 | < 0.0001 |
| Waist circumference | 89.19 | 13.15 | 96.24 | 13.70 | -133.74 | < 0.0001 | 0.53 | < 0.0001 |
| Weight | 77.12 | 15.60 | 83.04 | 16.83 | -91.90 | < 0.0001 | 0.37 | < 0.0001 |
| Whole body fat free mass | 52.62 | 11.38 | 56.44 | 11.59 | -84.27 | < 0.0001 | 0.33 | < 0.0001 |
| Whole body fat mass | 24.54 | 9.41 | 26.57 | 10.17 | -51.54 | < 0.0001 | 0.21 | < 0.0001 |
| Whole body water mass | 38.51 | 8.33 | 41.31 | 8.49 | -84.43 | < 0.0001 | 0.34 | < 0.0001 |

**Supplementary Table S8.** Model 4. StepAIC regression model after debugging the model for physical measures with high multicollinearity (VIF >5).

| Predictor | Estimate | Std Error | Z | p | OR | Lower CI | Upper CI | FDR |
|---|---|---|---|---|---|---|---|---|
| (Intercept) | -2.34 | 0.01 | -369.27 | <0.001 | 0.10 | 0.10 | 0.10 | <0.001 |
| Waist circumference | 0.37 | 0.01 | 28.55 | <0.001 | 1.44 | 1.41 | 1.48 | <0.001 |
| Hip circumference | -0.11 | 0.01 | -10.27 | <0.001 | 0.89 | 0.87 | 0.91 | <0.001 |
| Seated height | -0.04 | 0.01 | -4.96 | <0.001 | 0.96 | 0.95 | 0.98 | <0.001 |
| Weight | 0.11 | 0.03 | 3.79 | 0.0001 | 1.11 | 1.05 | 1.18 | 0.0002 |
| Impedance of leg-left | -0.06 | 0.01 | -6.43 | <0.001 | 0.94 | 0.92 | 0.96 | <0.001 |
| Impedance of arm-right | 0.02 | 0.01 | 1.94 | 0.0523 | 1.02 | 1.00 | 1.04 | 0.0523 |
| Leg fat free mass-left | -0.17 | 0.03 | -6.14 | <0.001 | 0.84 | 0.80 | 0.89 | <0.001 |
| Trunk fat percentage | 0.05 | 0.01 | 4.53 | <0.001 | 1.06 | 1.03 | 1.08 | <0.001 |
| Age at recruitment | 0.42 | 0.01 | 62.31 | <0.001 | 1.52 | 1.50 | 1.54 | <0.001 |
| Sex | 0.42 | 0.01 | 29.08 | <0.001 | 1.52 | 1.48 | 1.56 | <0.001 |
| PWA | 0.30 | 0.01 | 54.09 | <0.001 | 1.36 | 1.34 | 1.37 | <0.001 |
| BMD | -0.02 | 0.00 | -3.07 | 0.0022 | 0.99 | 0.98 | 0.99 | 0.0023 |
| cIMT | 1.29 | 0.01 | 187.84 | <0.001 | 3.64 | 3.60 | 3.69 | <0.001 |
| FEV1 | -0.32 | 0.01 | -45.44 | <0.001 | 0.73 | 0.72 | 0.74 | <0.001 |

Overall model performance: $R^2_{McFadden}$: 0.273. $R^2_{Nagelkerke}$: 0.361. AIC: 271174.90. Chi-cuadrado global: 102033, df: 14, p = 0.

**Supplementary Table S9.** Lifestyle data attending to the presence or not of a cardiovascular event during the study period.

| | No CVD (n=191376) | | CVD (n=22129) | | t | p value | Cohen d | FDR |
|---|---|---|---|---|---|---|---|---|
| Variable | Mean | sd | Mean | sd | | | | |
| Age at recruitment | 52.12 | 6.99 | 56.63 | 6.75 | -93.71 | < 0.0001 | 0.65 | < 0.0001 |
| Children fathered | 1.72 | 1.30 | 1.89 | 1.33 | -14.55 | < 0.0001 | 0.13 | < 0.0001 |
| Distance to job | 12.62 | 79.94 | 14.25 | 108.10 | -2.08 | 0.0371 | 0.02 | < 0.0001 |
| Length of working week | 35.18 | 13.06 | 35.37 | 13.79 | -1.90 | 0.0572 | 0.01 | 0.0009 |
| MET for all activity | 2581.32 | 2677.85 | 2724.60 | 2932.71 | -6.94 | < 0.0001 | 0.05 | 0.8840 |
| MET for moderate | 831.94 | 1153.05 | 914.31 | 1252.40 | -9.34 | < 0.0001 | 0.07 | 0.0103 |
| MET for vigorous | 703.83 | 1169.39 | 695.10 | 1291.48 | 0.96 | 0.3371 | -0.01 | < 0.0001 |
| MET for walking | 1045.55 | 1120.04 | 1115.18 | 1187.24 | -8.31 | < 0.0001 | 0.06 | < 0.0001 |
| Number of medications | 1.64 | 1.94 | 3.06 | 2.81 | -72.95 | < 0.0001 | 0.69 | < 0.0001 |
| Sleep duration | 7.05 | 0.98 | 6.96 | 1.10 | 11.40 | < 0.0001 | -0.09 | < 0.0001 |
| Summed days activity | 10.45 | 4.84 | 10.29 | 5.01 | 4.64 | < 0.0001 | -0.03 | < 0.0001 |
| Summed minutes activity | 125.59 | 101.13 | 130.86 | 111.28 | -6.73 | < 0.0001 | 0.05 | 0.9457 |
| Time driving | -1.68 | 5.24 | -0.86 | 5.10 | -22.70 | < 0.0001 | 0.16 | < 0.0001 |
| Time in current job | 12.12 | 11.38 | 13.52 | 12.43 | -15.94 | < 0.0001 | 0.12 | < 0.0001 |
| Time using computer | -1.43 | 5.13 | -0.81 | 4.73 | -18.24 | < 0.0001 | 0.12 | < 0.0001 |
| Time watching TV | 1.64 | 3.31 | 2.15 | 2.96 | -23.86 | < 0.0001 | 0.16 | < 0.0001 |
| Townsend index | -1.45 | 2.94 | -1.26 | 3.02 | -9.02 | < 0.0001 | 0.07 | < 0.0001 |
| Travelling to job | 4.10 | 2.46 | 4.07 | 2.70 | 1.75 | 0.0794 | -0.01 | 0.1766 |

**Supplementary Table S10.** Model 4. StepAIC regression model after debugging the model for lifestyle variables with high multicollinearity (VIF >5).

| **Predictor** | **Estimate** | **Std Error** | **Z** | **p** | **Odds Ratio** | **Lower CI** | **Upper CI** | **FDR** |
|---|---|---|---|---|---|---|---|---|
| (Intercept) | -2.16 | 0.02 | -93.24 | <0.0001 | 0.12 | 0.11 | 0.12 | <0.0001 |
| Above any recommendation | 0.03 | 0.02 | 1.39 | 0.1631 | 1.03 | 0.99 | 1.06 | 0.2292 |
| Above moderate recomms | 0.03 | 0.02 | 1.82 | 0.0682 | 1.03 | 1.00 | 1.07 | 0.1223 |
| Age at recruitment | 0.53 | 0.02 | 34.28 | <0.0001 | 1.69 | 1.64 | 1.74 | <0.0001 |
| Alcohol intake | 0.05 | 0.02 | 3.01 | 0.0026 | 1.05 | 1.02 | 1.09 | 0.0080 |
| Alcohol intake versus 10y | 0.02 | 0.01 | 1.47 | 0.1408 | 1.02 | 0.99 | 1.05 | 0.2075 |
| Broken bones in 5y | 0.00 | 0.01 | -0.09 | 0.9293 | 1.00 | 0.97 | 1.02 | 0.9475 |
| Cancer | 0.00 | 0.01 | 0.05 | 0.9599 | 1.00 | 0.98 | 1.03 | 0.9599 |
| Children fathered | 0.02 | 0.01 | 1.83 | 0.0666 | 1.02 | 1.00 | 1.05 | 0.1223 |
| Current employ_2 | -0.01 | 0.01 | -0.73 | 0.4644 | 0.99 | 0.96 | 1.02 | 0.5260 |
| Diabetes | -0.02 | 0.01 | -2.48 | 0.0132 | 0.98 | 0.96 | 0.99 | 0.0342 |
| Distance to job | 0.00 | 0.01 | -0.21 | 0.8326 | 1.00 | 0.98 | 1.01 | 0.8659 |
| Ethnic background | 0.03 | 0.02 | 2.13 | 0.0335 | 1.03 | 1.00 | 1.07 | 0.0792 |
| Falls last year | -0.02 | 0.01 | -1.20 | 0.2297 | 0.98 | 0.96 | 1.01 | 0.2986 |
| Had major operations | 0.10 | 0.01 | 8.44 | <0.0001 | 1.11 | 1.08 | 1.13 | <0.0001 |
| Home area | -0.02 | 0.01 | -1.22 | 0.2220 | 0.98 | 0.96 | 1.01 | 0.2959 |
| IPAQ group | -0.01 | 0.02 | -0.41 | 0.6783 | 0.99 | 0.94 | 1.04 | 0.7505 |
| Job at visit | 0.02 | 0.01 | 1.27 | 0.2044 | 1.02 | 0.99 | 1.04 | 0.2798 |
| Job heavy work | -0.01 | 0.02 | -0.73 | 0.4653 | 0.99 | 0.96 | 1.02 | 0.5260 |
| Job mainly standing | 0.05 | 0.02 | 2.94 | 0.0033 | 1.05 | 1.02 | 1.09 | 0.0096 |
| Job shift work | -0.02 | 0.01 | -1.65 | 0.0996 | 0.98 | 0.95 | 1.00 | 0.1570 |
| Length of working week | 0.02 | 0.02 | 1.63 | 0.1028 | 1.03 | 1.00 | 1.06 | 0.1572 |
| Long standing illness | 0.07 | 0.01 | 5.40 | <0.0001 | 1.07 | 1.05 | 1.10 | <0.0001 |
| Mean hand grip | 0.00 | 0.02 | 0.29 | 0.7727 | 1.00 | 0.97 | 1.04 | 0.8200 |
| Medication pain _1 | 0.10 | 0.01 | 6.75 | <0.0001 | 1.10 | 1.07 | 1.13 | <0.0001 |
| Medication pain _2 | -0.04 | 0.01 | -3.18 | 0.0015 | 0.96 | 0.93 | 0.98 | 0.0051 |
| MET for vigorous | 0.03 | 0.01 | 1.93 | 0.0541 | 1.03 | 1.00 | 1.06 | 0.1083 |
| Mineral supplements_1 | -0.05 | 0.02 | -3.36 | 0.0008 | 0.95 | 0.92 | 0.98 | 0.0029 |
| Mineral supplements_2 | -0.06 | 0.02 | -3.53 | 0.0004 | 0.95 | 0.92 | 0.98 | 0.0016 |
| Number of medications | 0.41 | 0.02 | 26.48 | <0.0001 | 1.51 | 1.47 | 1.56 | <0.0001 |
| Other Conditions | 0.05 | 0.01 | 4.21 | <0.0001 | 1.05 | 1.03 | 1.08 | 0.0001 |
| Other medications | 0.04 | 0.01 | 3.03 | 0.0024 | 1.05 | 1.02 | 1.08 | 0.0079 |
| Overall health rating | 0.15 | 0.01 | 10.99 | <0.0001 | 1.17 | 1.14 | 1.20 | <0.0001 |
| Qualifications_1 | 0.00 | 0.01 | 0.34 | 0.7338 | 1.00 | 0.98 | 1.03 | 0.7950 |
| Qualifications_2 | -0.02 | 0.01 | -1.68 | 0.0921 | 0.98 | 0.95 | 1.00 | 0.1497 |
| Sleep duration | -0.02 | 0.01 | -1.80 | 0.0723 | 0.98 | 0.95 | 1.00 | 0.1252 |
| Smoking status | 0.10 | 0.01 | 7.66 | <0.0001 | 1.10 | 1.07 | 1.13 | <0.0001 |
| Stair climbing | -0.01 | 0.01 | -0.86 | 0.3887 | 0.99 | 0.96 | 1.01 | 0.4594 |
| Summed minutes activity | -0.02 | 0.02 | -1.12 | 0.2630 | 0.98 | 0.94 | 1.02 | 0.3258 |
| Time driving | 0.04 | 0.01 | 2.92 | 0.0035 | 1.04 | 1.01 | 1.07 | 0.0097 |
| Time in current job | -0.02 | 0.01 | -1.98 | 0.0478 | 0.98 | 0.96 | 1.00 | 0.1019 |
| Time using computer | 0.01 | 0.01 | 0.88 | 0.3773 | 1.01 | 0.99 | 1.04 | 0.4563 |
| Time watching TV | 0.03 | 0.01 | 1.83 | 0.0665 | 1.03 | 1.00 | 1.05 | 0.1223 |
| Townsend index | 0.03 | 0.01 | 1.98 | 0.0474 | 1.03 | 1.00 | 1.05 | 0.1019 |
| Transport type_1 | -0.02 | 0.02 | -1.12 | 0.2631 | 0.98 | 0.94 | 1.02 | 0.3258 |
| Transport type_2 | -0.02 | 0.01 | -1.46 | 0.1437 | 0.98 | 0.96 | 1.01 | 0.2075 |
| Travelling to job | 0.02 | 0.01 | 1.70 | 0.0886 | 1.02 | 1.00 | 1.04 | 0.1487 |
| Usual walking pace | -0.08 | 0.01 | -5.68 | <0.0001 | 0.92 | 0.90 | 0.95 | <0.0001 |
| Vitamin supplements_1 | -0.06 | 0.02 | -3.69 | 0.0002 | 0.95 | 0.92 | 0.97 | 0.0010 |
| Vitamin supplements_2 | -0.04 | 0.02 | -2.33 | 0.0197 | 0.97 | 0.94 | 0.99 | 0.0488 |
| Walking for pleasure | 0.03 | 0.01 | 1.97 | 0.0490 | 1.03 | 1.00 | 1.05 | 0.1019 |
| Weight compared 1y | 0.05 | 0.01 | 4.38 | <0.0001 | 1.06 | 1.03 | 1.08 | <0.0001 |

Overall model performance: $R^2_{McFadden}$: 0.120. $R^2_{Nagelkerke}$: 0.166. AIC: 44425.84. Chi-cuadrado global: 97881.71, df: 150649, p = 1.

### Supplementary Information 2.

#### Sensitivity analysis for the logistic regression analysis

**BIOMARKERS DATASET**

*Model 1. Raw multivariate logistic regression analysis*

| Predictor | Estimate | Std Error | Z | p |
|---|---|---|---|---|
| (Intercept) | -2.0937 | 0.0920 | -22.7540 | 0.0000 |
| Albumin | 0.0372 | 0.0829 | 0.4492 | 0.6533 |
| Alkaline phosphatase | 0.0042 | 0.0621 | 0.0672 | 0.9464 |
| Alanine aminotransferase | 0.0558 | 0.0975 | 0.5721 | 0.5673 |
| Apolipoprotein A | -0.2589 | 0.0830 | -3.1181 | 0.0018 |
| Apolipoprotein B | -0.1400 | 0.0821 | -1.7066 | 0.0879 |
| Aspartate aminotransferase | -0.0232 | 0.0888 | -0.2612 | 0.7939 |
| Direct bilirubin | 0.1500 | 0.0618 | 2.4274 | 0.0152 |
| Urea | 0.0825 | 0.0833 | 0.9905 | 0.3219 |
| Calcium | -0.0327 | 0.0829 | -0.3946 | 0.6931 |
| Creatinine | 0.0163 | 0.1089 | 0.1495 | 0.8812 |
| C reactive protein | -0.0031 | 0.0538 | -0.0572 | 0.9544 |
| Cystatin C | 0.4018 | 0.0909 | 4.4203 | 0.0000 |
| Gamma glutamyltransferase | 0.1706 | 0.0692 | 2.4645 | 0.0137 |
| Glucose | 0.0273 | 0.0817 | 0.3339 | 0.7384 |
| Glycated haemoglobin HbA1c | 0.2852 | 0.0793 | 3.5964 | 0.0003 |
| IGF 1 | -0.2274 | 0.0749 | -3.0379 | 0.0024 |
| Lipoprotein A | 0.0947 | 0.0644 | 1.4714 | 0.1412 |
| Oestradiol | -0.1989 | 0.1264 | -1.5740 | 0.1155 |
| Phosphate | 0.0452 | 0.0705 | 0.6405 | 0.5219 |
| Rheumatoid factor | 0.0569 | 0.0608 | 0.9357 | 0.3494 |
| SHBG | 0.0476 | 0.0803 | 0.5929 | 0.5532 |
| Testosterone | 0.2511 | 0.0702 | 3.5799 | 0.0003 |
| Total protein | -0.0738 | 0.0761 | -0.9688 | 0.3326 |
| Triglycerides | 0.1051 | 0.0878 | 1.1969 | 0.2314 |
| Urate | 0.1415 | 0.0863 | 1.6410 | 0.1008 |
| Vitamin D | 0.0388 | 0.0669 | 0.5801 | 0.5618 |

Overall model performance: $R^2_{McFadden}$: 0.186. $R^2_{Nagelkerke}$: 0.247. AIC: 1634.27. Chi-cuadrado global: 438639, df: 499580, p = 1.

*Model 2. Multivariate logistic regression analysis including only those variables with significant association in the raw model*

| Predictor | Estimate | Std Error | Z | p |
|---|---|---|---|---|
| (Intercept) | -1.8015 | 0.0055 | -326.7881 | <0.001 |
| Apolipoprotein A | -0.1826 | 0.0057 | -31.8938 | <0.001 |
| Direct bilirubin | 0.0817 | 0.0050 | 16.3850 | <0.001 |
| Cystatin C | 0.4555 | 0.0055 | 82.7981 | <0.001 |
| GGT | 0.0726 | 0.0042 | 17.1532 | <0.001 |
| Glycated haemoglobin-HbA1c | 0.2727 | 0.0044 | 61.7471 | <0.001 |
| IGF-1 | -0.1558 | 0.0053 | -29.4391 | <0.001 |
| Testosterone | 0.1882 | 0.0054 | 35.1727 | <0.001 |

Overall model performance: $R^2_{McFadden}$: 0.082. $R^2_{Nagelkerke}$: 0.120. AIC: 258223.27. Chi-cuadrado global: 182012, df: 187333, p = 1.

*Model 3. StepAIC multivariate logistic regression analysis including all variables (before collinearity analysis).*

| Predictor | Estimate | Std Error | Z | p |
|---|---|---|---|---|
| (Intercept) | -1.8624 | 0.0050 | -370.3394 | <0.0001 |
| Albumin | -0.0570 | 0.0053 | -10.7972 | <0.0001 |
| Alkaline phosphatase | 0.0395 | 0.0042 | 9.3572 | <0.0001 |
| Alanine aminotransferase | -0.1070 | 0.0064 | -16.6726 | <0.0001 |
| Apolipoprotein A | -0.1515 | 0.0050 | -30.3373 | <0.0001 |
| Apolipoprotein B | -0.2350 | 0.0045 | -52.5535 | <0.0001 |
| Aspartate aminotransferase | 0.0545 | 0.0058 | 9.3496 | <0.0001 |
| Direct bilirubin | 0.0550 | 0.0043 | 12.6867 | <0.0001 |
| Urea | 0.1184 | 0.0047 | 25.2694 | <0.0001 |
| Calcium | 0.0562 | 0.0051 | 10.9913 | <0.0001 |
| Creatinine | -0.1917 | 0.0062 | -31.1286 | <0.0001 |
| C reactive protein | 0.0178 | 0.0039 | 4.5535 | <0.0001 |
| Cystatin C | 0.3906 | 0.0057 | 68.0981 | <0.0001 |
| Gamma glutamyltransferase | 0.0713 | 0.0043 | 16.7275 | <0.0001 |
| Glucose | -0.0504 | 0.0047 | -10.6136 | <0.0001 |
| Glycated haemoglobin HbA1c | 0.2696 | 0.0050 | 53.9891 | <0.0001 |
| IGF 1 | -0.1435 | 0.0045 | -32.0954 | <0.0001 |
| Lipoprotein A | 0.1109 | 0.0039 | 28.1872 | <0.0001 |
| Oestradiol | -0.1093 | 0.0083 | -13.1313 | <0.0001 |
| Phosphate | 0.0367 | 0.0043 | 8.5901 | <0.0001 |
| Rheumatoid factor | 0.0282 | 0.0040 | 7.1333 | <0.0001 |
| SHBG | -0.0141 | 0.0056 | -2.5255 | 0.0116 |
| Testosterone | 0.2328 | 0.0051 | 45.4951 | <0.0001 |
| Total protein | -0.0627 | 0.0050 | -12.4501 | <0.0001 |
| Triglycerides | 0.0792 | 0.0044 | 17.9321 | <0.0001 |
| Urate | 0.1484 | 0.0052 | 28.5355 | <0.0001 |

Overall model performance: $R^2_{McFadden}$: 0.094. $R^2_{Nagelkerke}$: 0.136. AIC: 398674.49. Chi-cuadrado global: 41496.81, df: 25, p = 0.

### METABOLOMIC DATASET

*Model 1. Raw multivariate logistic regression analysis*

| Predictor | Estimate | Std Error | Z | p |
|---|---|---|---|---|
| (Intercept) | -1.8214 | 0.007 | -258.9426 | <0.0001 |
| Acetate | -0.0244 | 0.0075 | -3.2439 | 0.0012 |
| Acetoacetate | -0.0147 | 0.0108 | -1.3589 | 0.1742 |
| Acetone | 0.076 | 0.0104 | 7.332 | <0.0001 |
| Alanine | 0.0305 | 0.0074 | 4.1295 | <0.0001 |
| Albumin | -0.1681 | 0.0081 | -20.7646 | <0.0001 |
| Apolipoprotein B/Apolipoprotein A1 | 0.0102 | 0.0498 | 0.2045 | 0.8379 |
| Average Diameter for LDL Particles | -0.1125 | 0.0128 | -8.8089 | <0.0001 |
| Chol./T.Lipids in Medium LDL | -0.0602 | 0.0314 | -1.9169 | 0.0552 |
| Chol./T.Lipids in Very Large HDL | 0.0249 | 0.0266 | 0.9343 | 0.3501 |
| CE in Small HDL | 0.7072 | 0.0859 | 8.2301 | <0.0001 |
| CE in Very Small VLDL | 0.3973 | 0.0438 | 9.0665 | <0.0001 |
| CE/T.Lipids in IDL | -0.3553 | 0.0356 | -9.9769 | <0.0001 |
| CE/T.Lipids in Large LDL | 0.1716 | 0.0298 | 5.7611 | <0.0001 |
| CE/T.Lipids in Very Large HDL | -0.0418 | 0.025 | -1.6697 | 0.095 |
| Citrate | -0.0051 | 0.0078 | -0.6569 | 0.5113 |
| Creatinine | 0.2594 | 0.0077 | 33.7537 | <0.0001 |
| Free Chol. in Small LDL | -0.4934 | 0.0717 | -6.8851 | <0.0001 |
| Free Chol. in Very Large HDL | 0.134 | 0.0469 | 2.8587 | 0.0043 |
| Free Chol./T.Lipids in IDL | -0.0515 | 0.0175 | -2.9476 | 0.0032 |
| Free Chol./T.Lipids in Large HDL | -0.012 | 0.0192 | -0.6218 | 0.5341 |
| Free Chol./T.Lipids in Large VLDL | 0.0912 | 0.0244 | 3.732 | 0.0002 |
| Free Chol./T.Lipids in Small HDL | 0.2031 | 0.036 | 5.6444 | <0.0001 |
| Free Chol./T.Lipids in Very Large VLDL | -0.1519 | 0.0218 | -6.9745 | <0.0001 |
| Glucose | 0.0818 | 0.0061 | 13.4105 | <0.0001 |
| Glutamine | 0.1706 | 0.0077 | 22.0567 | <0.0001 |
| Glycine | -0.1766 | 0.0081 | -21.8128 | <0.0001 |
| Glycoprotein Acetyls | 0.0796 | 0.0096 | 8.3186 | <0.0001 |
| Histidine | -0.093 | 0.008 | -11.6617 | <0.0001 |
| Isoleucine | -0.1545 | 0.009 | -17.1284 | <0.0001 |
| Lactate | 0.0151 | 0.0086 | 1.7606 | 0.0783 |
| Linoleic Acid | -0.0918 | 0.0376 | -2.4381 | 0.0148 |
| Linoleic Acid/Total Fatty Acids | -0.3988 | 0.0244 | -16.3458 | <0.0001 |
| Omega 3/Total Fatty Acids | -0.1811 | 0.0137 | -13.1865 | <0.0001 |
| Omega 6/Omega 3 | -0.0312 | 0.012 | -2.6031 | 0.0092 |
| Phenylalanine | 0.0275 | 0.0076 | 3.5937 | 0.0003 |
| PH in Medium HDL | -0.6745 | 0.0811 | -8.3177 | <0.0001 |
| PH/T.Lipids in Chylomicrons | 0.1309 | 0.0121 | 10.8122 | <0.0001 |
| PH/T.Lipids in IDL | -0.208 | 0.0222 | -9.3473 | <0.0001 |
| PH/T.Lipids in Large HDL | 0.1134 | 0.021 | 5.4018 | <0.0001 |
| PH/T.Lipids in Large LDL | 0.0797 | 0.0298 | 2.6742 | 0.0075 |
| PH/T.Lipids in Large VLDL | -0.1382 | 0.0343 | -4.0284 | 0.0001 |
| PH/T.Lipids in Medium HDL | -0.1763 | 0.0339 | -5.2076 | <0.0001 |

| | | | | |
|---|---|---|---|---|
| PH/T.Lipids in Medium LDL | -0.0267 | 0.0239 | -1.1149 | 0.2649 |
| PH/T.Lipids in Small HDL | 0.1328 | 0.0375 | 3.5389 | 0.0004 |
| PH/T.Lipids in Small LDL | 0.288 | 0.0201 | 14.2989 | <0.0001 |
| PH/T.Lipids in Very Large VLDL | 0.0357 | 0.0234 | 1.5267 | 0.1268 |
| PH/T.Lipids in Very Small VLDL | 0.0733 | 0.0177 | 4.137 | <0.0001 |
| Pyruvate | -0.0364 | 0.0084 | -4.3419 | <0.0001 |
| SFA Total Fatty Acids | -0.3042 | 0.0165 | -18.4057 | <0.0001 |
| Triglycerides in Large HDL | 0.0195 | 0.0337 | 0.5779 | 0.5633 |
| Triglycerides/T.Lipids in Chylomicrons | 0.0091 | 0.011 | 0.825 | 0.4094 |
| Triglycerides/T.Lipids in Very Large HDL | -0.0741 | 0.0143 | -5.1752 | <0.0001 |
| Tyrosine | 0.1271 | 0.0084 | 15.1758 | <0.0001 |

Chol.: cholesterol. PH: phospholipids. T.Lipids: Total Lipids. Overall model performance: $R^2_{McFadden}$: 0.090. $R^2_{Nagelkerke}$: 0.130. AIC: 160354.82. Chi-cuadrado global: 59866.52, df: 49637, p = 0.

*Model 2. Multivariate logistic regression analysis including only those variables with significant association in the raw model*

| Predictor | Estimate | Std Error | Z | p |
|---|---|---|---|---|
| (Intercept) | -1.8242 | 0.0063 | -289.6242 | <0.0001 |
| Acetate | -0.0221 | 0.0065 | -3.4085 | 0.0007 |
| Acetone | 0.0653 | 0.0062 | 10.5333 | <0.0001 |
| Alanine | 0.0411 | 0.0065 | 6.3276 | <0.0001 |
| Albumin | -0.1625 | 0.0070 | -23.0543 | <0.0001 |
| Average Diameter for LDL Particles | -0.1194 | 0.0102 | -11.7446 | <0.0001 |
| CE in Small HDL | 0.7587 | 0.0673 | 11.2741 | <0.0001 |
| CE in Very Small VLDL | 0.4157 | 0.0285 | 14.5639 | <0.0001 |
| CE/T. Lipids in IDL | -0.3722 | 0.0265 | -14.0195 | <0.0001 |
| CE/T. Lipids in Large LDL | 0.1364 | 0.0205 | 6.6484 | <0.0001 |
| Creatinine | 0.2530 | 0.0067 | 37.5351 | <0.0001 |
| Free Chol. in Small LDL | -0.5688 | 0.0582 | -9.7694 | <0.0001 |
| Free Chol. in Very Large HDL | 0.1766 | 0.0364 | 4.8503 | <0.0001 |
| Free Chol./T. Lipids in IDL | -0.0595 | 0.0142 | -4.1981 | <0.0001 |
| Free Chol./T. Lipids in Large VLDL | 0.0722 | 0.0191 | 3.7895 | 0.0002 |
| Free Chol./T. Lipids in Small HDL | 0.2047 | 0.0275 | 7.4520 | <0.0001 |
| Free Chol./T. Lipids in Very Large VLDL | -0.1301 | 0.0141 | -9.2465 | <0.0001 |
| Glucose | 0.0796 | 0.0052 | 15.2017 | <0.0001 |
| Glutamine | 0.1671 | 0.0068 | 24.7161 | <0.0001 |
| Glycine | -0.1861 | 0.0072 | -25.8695 | <0.0001 |
| Glycoprotein Acetyls | 0.0755 | 0.0084 | 8.9386 | <0.0001 |
| Histidine | -0.0902 | 0.0071 | -12.7159 | <0.0001 |
| Isoleucine | -0.1521 | 0.0081 | -18.8892 | <0.0001 |
| Linoleic Acid | -0.0670 | 0.0269 | -2.4890 | 0.0128 |
| Linoleic Acid/Total Fatty Acids | -0.4012 | 0.0196 | -20.4558 | <0.0001 |
| Omega 3/Total Fatty Acids | -0.1735 | 0.0120 | -14.4330 | <0.0001 |
| Omega 6/Omega 3 | -0.0265 | 0.0106 | -2.5071 | 0.0122 |
| Phenylalanine | 0.0270 | 0.0070 | 3.8751 | 0.0001 |
| PL in Medium HDL | -0.7053 | 0.0590 | -11.9532 | <0.0001 |
| PL/T. Lipids in Chylomicrons | 0.1162 | 0.0094 | 12.3102 | <0.0001 |
| PL/T. Lipids in IDL | -0.2048 | 0.0191 | -10.7080 | <0.0001 |
| PL/T. Lipids in Large HDL | 0.1251 | 0.0140 | 8.9172 | <0.0001 |
| PL/T. Lipids in Large LDL | 0.0521 | 0.0221 | 2.3576 | 0.0184 |
| PL/T. Lipids in Large VLDL | -0.0774 | 0.0191 | -4.0549 | 0.0001 |
| PL/T. Lipids in Medium HDL | -0.1334 | 0.0226 | -5.9025 | <0.0001 |
| PL/T. Lipids in Small HDL | 0.1066 | 0.0302 | 3.5360 | 0.0004 |
| PL/T. Lipids in Small LDL | 0.2922 | 0.0157 | 18.5766 | <0.0001 |
| PL/T. Lipids in Very Small VLDL | 0.0670 | 0.0147 | 4.5510 | <0.0001 |
| Pyruvate | -0.0276 | 0.0063 | -4.3665 | <0.0001 |
| SFA Total Fatty Acids | -0.3003 | 0.0139 | -21.6755 | <0.0001 |
| Triglycerides/T. Lipids in Very Large HDL | -0.0542 | 0.0096 | -5.6402 | <0.0001 |
| Tyrosine | 0.1272 | 0.0075 | 17.0131 | <0.0001 |

Overall model performance: $R^2_{McFadden}$: 0.091. $R^2_{Nagelkerke}$: 0.132. AIC: 200227.10. Chi-cuadrado global: 19970.24, df: 41, p = 0.

*Model 3. StepAIC multivariate logistic regression analysis including all variables (before collinearity analysis).*

| Predictor | Estimate | Std Error | Z | p |
|---|---|---|---|---|
| (Intercept) | -1.82 | 0.01 | -289.62 | <0.0001 |
| Acetate | -0.02 | 0.01 | -3.41 | 0.0007 |
| Acetone | 0.07 | 0.01 | 10.53 | <0.0001 |
| Alanine | 0.04 | 0.01 | 6.33 | <0.0001 |
| Albumin | -0.16 | 0.01 | -23.05 | <0.0001 |
| Average Diameter for LDL Particles | -0.12 | 0.01 | -11.74 | <0.0001 |
| CE in Small HDL | 0.76 | 0.07 | 11.27 | <0.0001 |
| CE in Very Small VLDL | 0.42 | 0.03 | 14.56 | <0.0001 |
| CE/T.Lipids in IDL | -0.37 | 0.03 | -14.02 | <0.0001 |
| CE/T.Lipids in Large LDL | 0.14 | 0.02 | 6.65 | <0.0001 |
| Creatinine | 0.25 | 0.01 | 37.54 | <0.0001 |
| Free Chol. in Small LDL | -0.57 | 0.06 | -9.77 | <0.0001 |
| Free Chol. in Very Large HDL | 0.18 | 0.04 | 4.85 | <0.0001 |
| Free Chol./T.Lipids in IDL | -0.06 | 0.01 | -4.20 | <0.0001 |
| Free Chol./T.Lipids in Large VLDL | 0.07 | 0.02 | 3.79 | 0.0002 |
| Free Chol./T.Lipids in Small HDL | 0.20 | 0.03 | 7.45 | <0.0001 |
| Free Chol./T.Lipids in Very Large VLDL | -0.13 | 0.01 | -9.25 | <0.0001 |
| Glucose | 0.08 | 0.01 | 15.20 | <0.0001 |
| Glutamine | 0.17 | 0.01 | 24.72 | <0.0001 |
| Glycine | -0.19 | 0.01 | -25.87 | <0.0001 |
| Glycoprotein Acetyls | 0.08 | 0.01 | 8.94 | <0.0001 |
| Histidine | -0.09 | 0.01 | -12.72 | <0.0001 |
| Isoleucine | -0.15 | 0.01 | -18.89 | <0.0001 |
| Linoleic Acid | -0.07 | 0.03 | -2.49 | 0.0128 |
| Linoleic Acid/T.Fatty Acids | -0.40 | 0.02 | -20.46 | <0.0001 |
| Omega 3/T.Fatty Acids | -0.17 | 0.01 | -14.43 | <0.0001 |
| Omega 6/Omega 3 | -0.03 | 0.01 | -2.51 | 0.0122 |
| Phenylalanine | 0.03 | 0.01 | 3.88 | 0.0001 |
| PL in Medium HDL | -0.71 | 0.06 | -11.95 | <0.0001 |
| PL/T.Lipids in Chylomicrons | 0.12 | 0.01 | 12.31 | <0.0001 |
| PL/T.Lipids in IDL | -0.20 | 0.02 | -10.71 | <0.0001 |
| PL/T.Lipids in Large HDL | 0.13 | 0.01 | 8.92 | <0.0001 |
| PL/T.Lipids in Large LDL | 0.05 | 0.02 | 2.36 | 0.0184 |
| PL/T.Lipids in Large VLDL | -0.08 | 0.02 | -4.05 | 0.0001 |
| PL/T.Lipids in Medium HDL | -0.13 | 0.02 | -5.90 | <0.0001 |
| PL/T.Lipids in Small HDL | 0.11 | 0.03 | 3.54 | 0.0004 |
| PL/T.Lipids in Small LDL | 0.29 | 0.02 | 18.58 | <0.0001 |
| PL/T.Lipids in Very Small VLDL | 0.07 | 0.01 | 4.55 | <0.0001 |
| Pyruvate | -0.03 | 0.01 | -4.37 | <0.0001 |
| SFA T.Fatty Acids | -0.30 | 0.01 | -21.68 | <0.0001 |
| TG/T.Lipids in Very Large HDL | -0.05 | 0.01 | -5.64 | <0.0001 |
| Tyrosine | 0.13 | 0.01 | 17.01 | <0.0001 |

Overall model performance: $R^2_{McFadden}$: 0.091. $R^2_{Nagelkerke}$: 0.132. AIC: AIC: 200227.10. Chi-cuadrado global: 19970.24 df: 41, p = 0.

### DIETARY DATASET

*Model 1. Raw multivariate logistic regression analysis*

| Predictor | Estimate | Std Error | Z | p |
|---|---|---|---|---|
| (Intercept) | -2.0412 | 0.0102 | -200.5382 | 0.0000 |
| Added sugars and preserves | 0.0716 | 0.0094 | 7.6595 | 0.0000 |
| Allium vegetables | 0.0170 | 0.0109 | 1.5546 | 0.1200 |
| Animal fat spread lower fat | 0.0513 | 0.0096 | 5.3402 | 0.0000 |
| Animal fat spread normal | 0.0693 | 0.0124 | 5.5973 | 0.0000 |
| Apples and pears | 0.0241 | 0.0103 | 2.3323 | 0.0197 |
| Beef | 0.0344 | 0.0101 | 3.4030 | 0.0007 |
| Beer and cider | 0.0738 | 0.0091 | 8.1405 | 0.0000 |
| Berries | -0.0187 | 0.0108 | -1.7365 | 0.0825 |
| Biscuit cereal | 0.0656 | 0.0099 | 6.6086 | 0.0000 |
| Biscuits | 0.0309 | 0.0098 | 3.1602 | 0.0016 |
| Bran cereal | 0.0164 | 0.0101 | 1.6203 | 0.1052 |
| Breaded battered chicken | 0.0236 | 0.0092 | 2.5831 | 0.0098 |
| Breaded battered fish | 0.0003 | 0.0098 | 0.0327 | 0.9739 |
| Chocolate confectionery | -0.0694 | 0.0107 | -6.5110 | 0.0000 |
| Citrus | 0.0366 | 0.0099 | 3.7000 | 0.0002 |
| Coffee caffeinated | 0.0065 | 0.0115 | 0.5666 | 0.5710 |
| Coffee decaffeinated | 0.0653 | 0.0098 | 6.6706 | 0.0000 |
| Cream | -0.0174 | 0.0106 | -1.6448 | 0.1000 |
| Dried fruit | 0.0181 | 0.0100 | 1.8081 | 0.0706 |
| Egg and egg dishes | 0.0289 | 0.0097 | 2.9945 | 0.0027 |
| Fortified wine | 0.0043 | 0.0096 | 0.4489 | 0.6535 |
| Fried roast potatoes | 0.0139 | 0.0105 | 1.3237 | 0.1856 |
| Fruit juice | 0.0033 | 0.0099 | 0.3313 | 0.7404 |
| Full fat yogurt | -0.0688 | 0.0111 | -6.2164 | 0.0000 |
| Grain dishes added fat | 0.0044 | 0.0098 | 0.4477 | 0.6544 |
| Green leafy cabbages | 0.0099 | 0.0109 | 0.9111 | 0.3622 |
| High fat cheese | -0.0635 | 0.0108 | -5.8850 | 0.0000 |
| Lamb | 0.0235 | 0.0095 | 2.4646 | 0.0137 |
| Legumes and pulses | -0.0017 | 0.0097 | -0.1715 | 0.8639 |
| Low fat yogurt | -0.0105 | 0.0104 | -1.0075 | 0.3137 |
| Low non sugar sweetened beverages | 0.0252 | 0.0097 | 2.5939 | 0.0095 |
| Mashed potatoes | 0.0226 | 0.0100 | 2.2712 | 0.0231 |
| Meat substitutes soy | -0.0025 | 0.0111 | -0.2258 | 0.8213 |
| Meat substitutes vegetarian | -0.0372 | 0.0116 | -3.1932 | 0.0014 |
| Medium and low-fat cheese | -0.0013 | 0.0099 | -0.1275 | 0.8985 |
| Milk based and powdered drinks | 0.0494 | 0.0093 | 5.3374 | 0.0000 |
| Milk dairy desserts | 0.0409 | 0.0095 | 4.3209 | 0.0000 |
| Mixed bread 50 50 brown and seeded | -0.0064 | 0.0118 | -0.5428 | 0.5873 |
| Muesli | -0.0091 | 0.0113 | -0.7995 | 0.4240 |
| Nut based spreads | -0.0451 | 0.0112 | -4.0154 | 0.0001 |
| Oat cereal non sugar | 0.0437 | 0.0108 | 4.0404 | 0.0001 |
| Oat cereal sugar | 0.0024 | 0.0102 | 0.2399 | 0.8104 |
| Oily fish | 0.0443 | 0.0098 | 4.5241 | 0.0000 |
| Olive oil drizzling dunking | -0.0038 | 0.0109 | -0.3480 | 0.7279 |
| Other bread | 0.0016 | 0.0102 | 0.1586 | 0.8739 |
| Other cereal sugar | 0.0330 | 0.0103 | 3.2042 | 0.0014 |
| Other desserts and cakes and pastries | -0.0302 | 0.0102 | -2.9697 | 0.0030 |
| Other fruit | 0.0371 | 0.0106 | 3.5100 | 0.0004 |
| Other meat | 0.0109 | 0.0093 | 1.1737 | 0.2405 |
| Other sweets | 0.0143 | 0.0095 | 1.5089 | 0.1313 |
| Other vegetables | 0.0019 | 0.0118 | 0.1584 | 0.8741 |
| Peas and sweetcorn | 0.0096 | 0.0098 | 0.9850 | 0.3246 |
| Pizza | -0.0541 | 0.0110 | -4.9212 | 0.0000 |
| Plant based spread lower fat | 0.1272 | 0.0107 | 11.9373 | 0.0000 |
| Plant based spread normal | 0.0931 | 0.0115 | 8.0925 | 0.0000 |
| Pork | 0.0367 | 0.0093 | 3.9407 | 0.0001 |
| Potatoes sweet potatoe | 0.0486 | 0.0103 | 4.7131 | 0.0000 |
| Poultry | -0.0216 | 0.0105 | -2.0519 | 0.0402 |

| | | | | |
|---|---|---|---|---|
| Processed meat | 0.0614 | 0.0097 | 6.3551 | 0.0000 |
| Raw salad | -0.0344 | 0.0117 | -2.9415 | 0.0033 |
| Red wine | 0.0518 | 0.0097 | 5.3203 | 0.0000 |
| Rice oat milk | -0.0125 | 0.0111 | -1.1258 | 0.2602 |
| Root vegetables | -0.0173 | 0.0115 | -1.5029 | 0.1329 |
| Salted nuts and seeds | -0.0198 | 0.0102 | -1.9393 | 0.0525 |
| Samosa pakora | 0.0019 | 0.0102 | 0.1893 | 0.8498 |
| Sauces condiments high fat | -0.0597 | 0.0106 | -5.6534 | 0.0000 |
| Sauces condiments low fat | -0.0270 | 0.0111 | -2.4324 | 0.0150 |
| Savoury crackers | 0.0252 | 0.0102 | 2.4582 | 0.0140 |
| Savoury snacks | -0.0575 | 0.0104 | -5.5450 | 0.0000 |
| Semi skimmed milk | -0.0409 | 0.0135 | -3.0209 | 0.0025 |
| Shellfish | 0.0065 | 0.0100 | 0.6461 | 0.5182 |
| Skimmed milk | 0.0038 | 0.0121 | 0.3100 | 0.7566 |
| Soups | 0.0510 | 0.0095 | 5.3653 | 0.0000 |
| Soy desserts and yogurt | 0.0092 | 0.0096 | 0.9658 | 0.3341 |
| Soy milk | -0.0244 | 0.0112 | -2.1813 | 0.0292 |
| Spirits | 0.0620 | 0.0087 | 7.1053 | 0.0000 |
| Stewed fruit | 0.0193 | 0.0099 | 1.9573 | 0.0503 |
| Sugar sweetened drinks | 0.0146 | 0.0098 | 1.4845 | 0.1377 |
| Sushi | -0.0131 | 0.0113 | -1.1549 | 0.2481 |
| Tea | 0.0037 | 0.0127 | 0.2952 | 0.7678 |
| Tea decaffeinated | -0.0594 | 0.0115 | -5.1559 | 0.0000 |
| Tomatoes | -0.0123 | 0.0113 | -1.0856 | 0.2777 |
| Unsalted nuts and seeds | -0.0141 | 0.0106 | -1.3228 | 0.1859 |
| Vegetable dips | -0.0407 | 0.0115 | -3.5288 | 0.0004 |
| Vegetable side dishes | -0.0003 | 0.0101 | -0.0328 | 0.9739 |
| Water | -0.0587 | 0.0108 | -5.4152 | 0.0000 |
| White bread | 0.0226 | 0.0124 | 1.8191 | 0.0689 |
| White fish and tinned tuna | 0.0011 | 0.0100 | 0.1116 | 0.9111 |
| White pasta and rice | -0.1139 | 0.0116 | -9.7943 | 0.0000 |
| White wine | -0.0502 | 0.0111 | -4.5368 | 0.0000 |
| Whole milk | -0.0388 | 0.0110 | -3.5269 | 0.0004 |
| Wholemeal bread | -0.0138 | 0.0123 | -1.1224 | 0.2617 |
| Wholemeal pasta | -0.0372 | 0.0110 | -3.3776 | 0.0007 |

Overall model performance: $R^2_{McFadden}$: 0.027. $R^2_{Nagelkerke}$: 0.038. AIC: 73378.15. Chi-cuadrado global: 20854.25, df: 25423, p = 1.

*Model 2. Multivariate logistic regression analysis including only those variables with significant association in the raw model*

| Predictor | Estimate | Std Error | Z | p |
|---|---|---|---|---|
| (Intercept) | -2.0395 | 0.0091 | -224.3738 | 0.0000 |
| Added sugars and preserves | 0.0728 | 0.0083 | 8.7533 | 0.0000 |
| Animal fat spread lower fat | 0.0479 | 0.0082 | 5.8673 | 0.0000 |
| Animal fat spread normal | 0.0701 | 0.0094 | 7.4374 | 0.0000 |
| Apples and pears | 0.0280 | 0.0090 | 3.0949 | 0.0020 |
| Beef | 0.0371 | 0.0088 | 4.2183 | 0.0000 |
| Beer and cider | 0.0697 | 0.0080 | 8.7068 | 0.0000 |
| Biscuit cereal | 0.0650 | 0.0084 | 7.7704 | 0.0000 |
| Biscuits | 0.0390 | 0.0086 | 4.5240 | 0.0000 |
| Breaded battered chicken | 0.0248 | 0.0081 | 3.0450 | 0.0023 |
| Chocolate confectionery | -0.0657 | 0.0094 | -6.9613 | 0.0000 |
| Citrus | 0.0254 | 0.0089 | 2.8649 | 0.0042 |
| Coffee decaffeinated | 0.0576 | 0.0081 | 7.1397 | 0.0000 |
| Egg and egg dishes | 0.0294 | 0.0085 | 3.4493 | 0.0006 |
| Full fat yogurt | -0.0620 | 0.0098 | -6.3294 | 0.0000 |
| High fat cheese | -0.0608 | 0.0095 | -6.3776 | 0.0000 |
| Lamb | 0.0251 | 0.0084 | 2.9813 | 0.0029 |
| Low non sugar sugar sweetened beverages | 0.0259 | 0.0086 | 3.0208 | 0.0025 |
| Mashed potatoes | 0.0261 | 0.0085 | 3.0659 | 0.0022 |
| Meat substitutes vegetarian | -0.0347 | 0.0103 | -3.3738 | 0.0007 |
| Milk based and powdered drinks | 0.0464 | 0.0082 | 5.6818 | 0.0000 |
| Milk dairy desserts | 0.0493 | 0.0083 | 5.9046 | 0.0000 |
| Nut based spreads | -0.0447 | 0.0099 | -4.4945 | 0.0000 |
| Oat cereal non sugar | 0.0398 | 0.0089 | 4.4523 | 0.0000 |
| Oily fish | 0.0401 | 0.0087 | 4.6068 | 0.0000 |
| Other cereal sugar | 0.0369 | 0.0087 | 4.2200 | 0.0000 |
| Other desserts and cakes and pastries | -0.0345 | 0.0090 | -3.8328 | 0.0001 |
| Other fruit | 0.0336 | 0.0091 | 3.6917 | 0.0002 |
| Pizza | -0.0487 | 0.0097 | -5.0328 | 0.0000 |
| Plant based spread lower fat | 0.1316 | 0.0080 | 16.4628 | 0.0000 |
| Plant based spread normal | 0.0963 | 0.0086 | 11.2387 | 0.0000 |
| Pork | 0.0374 | 0.0083 | 4.5270 | 0.0000 |
| Potatoes sweet potatoe | 0.0465 | 0.0086 | 5.3888 | 0.0000 |
| Poultry | -0.0147 | 0.0092 | -1.6038 | 0.1088 |
| Processed meat | 0.0598 | 0.0085 | 7.0415 | 0.0000 |
| Raw salad | -0.0436 | 0.0097 | -4.4992 | 0.0000 |
| Red wine | 0.0495 | 0.0086 | 5.7541 | 0.0000 |
| Sauces condiments high fat | -0.0645 | 0.0092 | -7.0242 | 0.0000 |
| Sauces condiments low fat | -0.0263 | 0.0098 | -2.6773 | 0.0074 |
| Savoury crackers | 0.0250 | 0.0090 | 2.7885 | 0.0053 |
| Savoury snacks | -0.0494 | 0.0091 | -5.4009 | 0.0000 |
| Semi skimmed milk | -0.0398 | 0.0094 | -4.2126 | 0.0000 |
| Soups | 0.0477 | 0.0085 | 5.6270 | 0.0000 |
| Soy milk | -0.0208 | 0.0096 | -2.1731 | 0.0298 |
| Spirits | 0.0614 | 0.0078 | 7.9111 | 0.0000 |
| Tea decaffeinated | -0.0609 | 0.0096 | -6.3617 | 0.0000 |
| Vegetable dips | -0.0503 | 0.0103 | -4.8983 | 0.0000 |
| Water | -0.0635 | 0.0094 | -6.7801 | 0.0000 |
| White pasta and rice | -0.1228 | 0.0101 | -12.0967 | 0.0000 |
| White wine | -0.0488 | 0.0098 | -5.0045 | 0.0000 |
| Whole milk | -0.0367 | 0.0094 | -3.9037 | 0.0001 |
| Wholemeal pasta | -0.0374 | 0.0096 | -3.8877 | 0.0001 |

Overall model performance: $R^2_{McFadden}$: 0.026. $R^2_{Nagelkerke}$: 0.037. AIC: 91696.58. Chi-cuadrado global: 2451.82, df: 51, p = 0.

*Model 3. StepAIC multivariate logistic regression analysis including all variables (before collinearity analysis).*

| Predictor | Estimate | Std Error | Z | p |
|---|---|---|---|---|
| (Intercept) | -2.0395 | 0.0091 | -224.3738 | 0.0000 |
| Added sugars and preserves | 0.0728 | 0.0083 | 8.7533 | 0.0000 |
| Animal fat spread lower fat | 0.0479 | 0.0082 | 5.8673 | 0.0000 |
| Animal fat spread normal | 0.0701 | 0.0094 | 7.4374 | 0.0000 |
| Apples and pears | 0.0280 | 0.0090 | 3.0949 | 0.0020 |
| Beef | 0.0371 | 0.0088 | 4.2183 | 0.0000 |
| Beer and cider | 0.0697 | 0.0080 | 8.7068 | 0.0000 |
| Biscuit cereal | 0.0650 | 0.0084 | 7.7704 | 0.0000 |
| Biscuits | 0.0390 | 0.0086 | 4.5240 | 0.0000 |
| Breaded battered chicken | 0.0248 | 0.0081 | 3.0450 | 0.0023 |
| Chocolate confectionery | -0.0657 | 0.0094 | -6.9613 | 0.0000 |
| Citrus | 0.0254 | 0.0089 | 2.8649 | 0.0042 |
| Coffee decaffeinated | 0.0576 | 0.0081 | 7.1397 | 0.0000 |
| Egg and egg dishes | 0.0294 | 0.0085 | 3.4493 | 0.0006 |
| Full fat yogurt | -0.0620 | 0.0098 | -6.3294 | 0.0000 |
| High fat cheese | -0.0608 | 0.0095 | -6.3776 | 0.0000 |
| Lamb | 0.0251 | 0.0084 | 2.9813 | 0.0029 |
| Low non sugar sugar sweetened beverages | 0.0259 | 0.0086 | 3.0208 | 0.0025 |
| Mashed potatoes | 0.0261 | 0.0085 | 3.0659 | 0.0022 |
| Meat substitutes vegetarian | -0.0347 | 0.0103 | -3.3738 | 0.0007 |
| Milk based and powdered drinks | 0.0464 | 0.0082 | 5.6818 | 0.0000 |
| Milk dairy desserts | 0.0493 | 0.0083 | 5.9046 | 0.0000 |
| Nut based spreads | -0.0447 | 0.0099 | -4.4945 | 0.0000 |
| Oat cereal non sugar | 0.0398 | 0.0089 | 4.4523 | 0.0000 |
| Oily fish | 0.0401 | 0.0087 | 4.6068 | 0.0000 |
| Other cereal sugar | 0.0369 | 0.0087 | 4.2200 | 0.0000 |
| Other desserts and cakes and pastries | -0.0345 | 0.0090 | -3.8328 | 0.0001 |
| Other fruit | 0.0336 | 0.0091 | 3.6917 | 0.0002 |
| Pizza | -0.0487 | 0.0097 | -5.0328 | 0.0000 |
| Plant based spread lower fat | 0.1316 | 0.0080 | 16.4628 | 0.0000 |
| Plant based spread normal | 0.0963 | 0.0086 | 11.2387 | 0.0000 |
| Pork | 0.0374 | 0.0083 | 4.5270 | 0.0000 |
| Potatoes sweet potatoe | 0.0465 | 0.0086 | 5.3888 | 0.0000 |
| Poultry | -0.0147 | 0.0092 | -1.6038 | 0.1088 |
| Processed meat | 0.0598 | 0.0085 | 7.0415 | 0.0000 |
| Raw salad | -0.0436 | 0.0097 | -4.4992 | 0.0000 |
| Red wine | 0.0495 | 0.0086 | 5.7541 | 0.0000 |
| Sauces condiments high fat | -0.0645 | 0.0092 | -7.0242 | 0.0000 |
| Sauces condiments low fat | -0.0263 | 0.0098 | -2.6773 | 0.0074 |
| Savoury crackers | 0.0250 | 0.0090 | 2.7885 | 0.0053 |
| Savoury snacks | -0.0494 | 0.0091 | -5.4009 | 0.0000 |
| Semi skimmed milk | -0.0398 | 0.0094 | -4.2126 | 0.0000 |
| Soups | 0.0477 | 0.0085 | 5.6270 | 0.0000 |
| Soy milk | -0.0208 | 0.0096 | -2.1731 | 0.0298 |
| Spirits | 0.0614 | 0.0078 | 7.9111 | 0.0000 |
| Tea decaffeinated | -0.0609 | 0.0096 | -6.3617 | 0.0000 |
| Vegetable dips | -0.0503 | 0.0103 | -4.8983 | 0.0000 |
| Water | -0.0635 | 0.0094 | -6.7801 | 0.0000 |
| White pasta and rice | -0.1228 | 0.0101 | -12.0967 | 0.0000 |
| White wine | -0.0488 | 0.0098 | -5.0045 | 0.0000 |
| Whole milk | -0.0367 | 0.0094 | -3.9037 | 0.0001 |
| Wholemeal pasta | -0.0374 | 0.0096 | -3.8877 | 0.0001 |

Overall model performance: $R^2_{McFadden}$: 0.026. $R^2_{Nagelkerke}$: 0.037. AIC: 91696.58. Chi-cuadrado global: 2451.82, df: 51, p = 0.

*Model 1. Raw multivariate logistic regression analysis*

| Predictor | Estimate | Std Error | Z | p |
|---|---|---|---|---|
| (Intercept) | -2.3411 | 0.0064 | -368.4642 | <0.0001 |
| Waist circumference | 0.3247 | 0.0131 | 24.7218 | <0.0001 |
| Hip circumference | -0.1392 | 0.0116 | -11.9862 | <0.0001 |
| Standing height | -0.1526 | 0.0339 | -4.4960 | <0.0001 |
| Seated height | 0.0295 | 0.0081 | 3.6632 | 0.0002 |
| Body mass index BMI | 0.2115 | 0.0583 | 3.6243 | 0.0003 |
| Weight | 0.0244 | 0.0776 | 0.3145 | 0.7532 |
| Impedance of leg-left | 0.0109 | 0.0172 | 0.6323 | 0.5272 |
| Impedance of arm-right | 0.0669 | 0.0148 | 4.5367 | <0.0001 |
| Leg fat free mass-left | -0.0156 | 0.0378 | -0.4120 | 0.6803 |
| Trunk fat percentage | -0.0063 | 0.0413 | -0.1520 | 0.8792 |
| Age at recruitment | 0.4682 | 0.0076 | 61.6842 | <0.0001 |
| Sex | 0.3890 | 0.0282 | 13.7988 | <0.0001 |
| PWA | 0.3114 | 0.0057 | 54.9929 | <0.0001 |
| BMD | -0.0183 | 0.0049 | -3.7437 | 0.0002 |
| cIMT | 1.2918 | 0.0069 | 187.7607 | <0.0001 |
| FEV1 | -0.2871 | 0.0072 | -39.9061 | <0.0001 |
| Body fat percentage | -0.0685 | 0.0672 | -1.0190 | 0.3082 |
| Whole body fat free mass | 0.0580 | 0.0487 | 1.1910 | 0.2337 |
| Whole body impedance | 0.0791 | 0.0273 | 2.8959 | 0.0038 |

Overall model performance: $R^2_{McFadden}$: 0.274. $R^2_{Nagelkerke}$: 0.362. AIC: 270793.73. Chi-cuadrado global: 102386.2, df: 15, p = 0.

*Model 2. Multivariate logistic regression analysis including only those variables with significant association in the raw model*

| Predictor | Estimate | Std Error | Z | p |
|---|---|---|---|---|
| (Intercept) | -2.3406 | 0.0063 | -368.7915 | <0.0001 |
| Waist circumference | 0.3209 | 0.0128 | 25.1016 | <0.0001 |
| Hip circumference | -0.1409 | 0.0114 | -12.3948 | <0.0001 |
| Standing height | -0.1212 | 0.0111 | -10.9097 | <0.0001 |
| Seated height | 0.0298 | 0.0081 | 3.6933 | 0.0002 |
| Body mass index BMI | 0.1780 | 0.0160 | 11.1473 | <0.0001 |
| Impedance of arm right | 0.0593 | 0.0130 | 4.5517 | <0.0001 |
| Age at recruitment | 0.4567 | 0.0063 | 72.1867 | <0.0001 |
| Sex | 0.4255 | 0.0108 | 39.4647 | <0.0001 |
| PWA | 0.3108 | 0.0057 | 54.9215 | <0.0001 |
| BMD | -0.0180 | 0.0049 | -3.6864 | 0.0002 |
| cIMT | 1.2918 | 0.0069 | 187.7870 | <0.0001 |
| FEV1 | -0.2870 | 0.0072 | -39.9545 | <0.0001 |
| Whole body impedance | 0.0344 | 0.0133 | 2.5855 | 0.0097 |

Overall model performance: $R^2_{McFadden}$: 0.274. $R^2_{Nagelkerke}$: 0.362. AIC: 23140.4217. Chi-cuadrado global: 102368.1, df: 13, p = 0.

*Model 3. StepAIC multivariate logistic regression analysis including all variables (before collinearity analysis).*

| Predictor | Estimate | Std Error | Z | p |
|---|---|---|---|---|
| (Intercept) | -2.3409 | 0.0063 | -368.6854 | <0.0001 |
| Waist circumference | 0.3261 | 0.0131 | 24.9739 | <0.0001 |
| Hip circumference | -0.1395 | 0.0116 | -12.0410 | <0.0001 |
| Standing height | -0.1445 | 0.0207 | -6.9727 | <0.0001 |
| Seated height | 0.0299 | 0.0081 | 3.7095 | 0.0002 |
| Body mass index (BMI) | 0.2229 | 0.0300 | 7.4202 | <0.0001 |
| Impedance of arm -right | 0.0582 | 0.0131 | 4.4460 | <0.0001 |
| Age at recruitment | 0.4688 | 0.0070 | 67.2798 | <0.0001 |
| Sex | 0.3849 | 0.0146 | 26.3167 | <0.0001 |
| PWA | 0.3115 | 0.0057 | 55.0201 | <0.0001 |
| BMD | -0.0181 | 0.0049 | -3.7024 | 0.0002 |
| cIMT | 1.2917 | 0.0069 | 187.7651 | <0.0001 |
| FEV1 | -0.2871 | 0.0072 | -39.9448 | <0.0001 |
| Body fat percentage | -0.0732 | 0.0256 | -2.8536 | 0.0043 |
| Whole body fat free mass | 0.0524 | 0.0334 | 1.5688 | 0.1167 |
| Whole body impedance | 0.0962 | 0.0200 | 4.8020 | <0.0001 |

Overall model performance: $R^2_{McFadden}$: 0.074. $R^2_{Nagelkerke}$: 0.097. AIC: 22547.6352. Chi-cuadrado global: 1791.906, df: 11, p = 0.

### LIFESTYLE DATASET

*Model 1. Raw multivariate logistic regression analysis*

| Predictor | Estimate | SE | Z | p |
|---|---|---|---|---|
| Sex | 0.9880 | 0.1848 | 5.3461 | 0.0000 |
| Age at recruitment | 0.1007 | 0.0131 | 7.6780 | 0.0000 |
| Number of days/week walked | 0.1657 | 0.1272 | 1.3033 | 0.1925 |
| Number of days/week of moderate physical activity | 0.1486 | 0.1303 | 1.1409 | 0.2539 |
| Duration of moderate activity | -0.0018 | 0.0021 | -0.9000 | 0.3681 |
| Number of days/week of vigorous physical activity | 0.0448 | 0.1207 | 0.3713 | 0.7104 |
| Usual walking pace | -0.2250 | 0.1305 | -1.7250 | 0.0845 |
| Frequency of friend/family visits | -0.0482 | 0.0695 | -0.6941 | 0.4876 |
| Time spend outdoors in summer | -0.0414 | 0.0437 | -0.9474 | 0.3434 |
| Time spent outdoors in winter | 0.0053 | 0.0538 | 0.0986 | 0.9215 |
| Time spent watching television (TV) | -0.0027 | 0.0553 | -0.0487 | 0.9612 |
| Time spent using computer | 0.0212 | 0.0448 | 0.4727 | 0.6364 |
| Length of mobile phone use | 0.0464 | 0.1047 | 0.4428 | 0.6579 |
| Weekly usage of mobile phone in last 3 months | 0.0008 | 0.0539 | 0.0156 | 0.9876 |
| Sleep duration | 0.0006 | 0.0720 | 0.0079 | 0.9937 |
| Getting up in morning | 0.0120 | 0.1128 | 0.1059 | 0.9156 |
| Morning/evening person (chronotype) | 0.0779 | 0.0886 | 0.8787 | 0.3796 |
| Nap during day | 0.1632 | 0.1229 | 1.3287 | 0.1840 |
| Sleeplessness / insomnia | 0.0385 | 0.1006 | 0.3825 | 0.7021 |
| Snoring | 0.0582 | 0.1423 | 0.4090 | 0.6825 |
| Daytime dozing / sleeping (narcolepsy) | 0.0179 | 0.1477 | 0.1210 | 0.9037 |
| Major dietary changes in the last 5 years | 0.0511 | 0.0775 | 0.6601 | 0.5092 |
| Overall health rating | 0.4516 | 0.1116 | 4.0480 | 0.0001 |
| Plays computer games | 0.1302 | 0.1359 | 0.9581 | 0.3380 |
| Use of sun/uv protection | -0.0913 | 0.0814 | -1.1219 | 0.2619 |
| Leisure/social activities | -0.1281 | 0.0607 | -2.1095 | 0.0349 |
| Types of transport used (excluding work) | -0.0468 | 0.1091 | -0.4287 | 0.6682 |
| Types of physical activity in last 4 weeks | -0.0042 | 0.0455 | -0.0922 | 0.9265 |
| Smoking status | 0.3834 | 0.1426 | 2.6898 | 0.0071 |
| Alcohol drinker status | -0.0847 | 0.2179 | -0.3887 | 0.6975 |
| Ever smoked | -0.4395 | 0.2069 | -2.1238 | 0.0337 |
| IPAQ activity group | 0.1699 | 0.1945 | 0.8735 | 0.3824 |
| Summed days activity | -0.0877 | 0.1173 | -0.7483 | 0.4543 |
| Summed minutes activity | 0.0030 | 0.0026 | 1.1617 | 0.2454 |
| Above moderate/vigorous recommendation | -0.2153 | 0.2247 | -0.9581 | 0.3380 |
| Above moderate/vigorous/walking recommendation | -0.2885 | 0.2891 | -0.9977 | 0.3184 |
| MET minutes per week for walking | 0.0000 | 0.0002 | 0.2675 | 0.7891 |
| MET minutes per week for vigorous activity | 0.0002 | 0.0002 | 1.3253 | 0.1851 |
| Summed MET minutes per week for all activity | -0.0002 | 0.0002 | -1.3269 | 0.1845 |
| Townsend deprivation index at recruitment | -0.0197 | 0.0307 | -0.6415 | 0.5212 |
| Type of accommodation lived in | 0.0708 | 0.3840 | 0.1843 | 0.8538 |
| Own or rent accommodation lived in | 0.0684 | 0.1028 | 0.6658 | 0.5056 |
| Length of time at current address | 0.0025 | 0.0070 | 0.3642 | 0.7157 |
| Number in household | -0.0359 | 0.0789 | -0.4551 | 0.6490 |
| Number of vehicles in household | 0.0605 | 0.0917 | 0.6597 | 0.5094 |
| Average total household income before tax | -0.0581 | 0.0866 | -0.6714 | 0.5019 |
| Time employed in main current job | 0.0023 | 0.0061 | 0.3822 | 0.7023 |
| Length of working week for main job | 0.0004 | 0.0065 | 0.0561 | 0.9552 |
| Frequency of travelling from home to job workplace | 0.0600 | 0.0339 | 1.7665 | 0.0773 |
| Job involves mainly walking or standing | 0.1034 | 0.0851 | 1.2156 | 0.2241 |
| Job involves heavy manual or physical work | -0.2230 | 0.1082 | -2.0617 | 0.0392 |
| Job involves shift work | 0.1542 | 0.0733 | 2.1034 | 0.0354 |
| Age completed full time education | -0.0146 | 0.0273 | -0.5356 | 0.5923 |
| Private healthcare | -0.0620 | 0.0947 | -0.6544 | 0.5128 |
| Qualifications (primary) | 0.0601 | 0.0553 | 1.0874 | 0.2769 |
| Gas or solid-fuel cooking/heating | -0.1118 | 0.1680 | -0.6654 | 0.5058 |
| Heating type(s) in home | 0.1074 | 0.1550 | 0.6933 | 0.4881 |
| How are people in household related to participant | 0.0891 | 0.0857 | 1.0402 | 0.2982 |
| Ethnic background | 0.0000 | 0.0001 | 0.3500 | 0.7263 |

Overall model performance: $R^2_{McFadden}$: 0.118. $R^2_{Nagelkerke}$: 0.163. AIC:35591.38. Chi-cuadrado global: 106716.2, df: 163203 p = 1.

*Model 2. Multivariate logistic regression analysis including only those variables with significant association in the raw model*

| Predictor | Estimate | SE | Z | p |
|---|---|---|---|---|
| Sex | 1.0176 | 0.0551 | 18.4826 | 0.0000 |
| Age at recruitment | 0.0860 | 0.0037 | 23.2050 | 0.0000 |
| Overall health rating | 0.5496 | 0.0367 | 14.9748 | 0.0000 |
| Leisure/social activities | -0.0143 | 0.0197 | -0.7270 | 0.4672 |
| Smoking status | 0.2121 | 0.0508 | 4.1775 | 0.0000 |
| Ever smoked | -0.1345 | 0.0706 | -1.9041 | 0.0569 |
| Job involves heavy manual or physical work | 0.0018 | 0.0325 | 0.0556 | 0.9557 |
| Job involves shift work | 0.0649 | 0.0320 | 2.0298 | 0.0424 |

Overall model performance: $R^2_{McFadden}$: 0.129. $R^2_{Nagelkerke}$: 0.167. AIC: 85109.54. Chi-cuadrado global: 57142.01, df: 60109, p = 1.

*Model 3. StepAIC multivariate logistic regression analysis including all variables (before collinearity analysis).*

| Predictor | Estimate | Std Error | Z | p |
|---|---|---|---|---|
| (Intercept) | -8.0861 | 0.2587 | -31.2514 | 0.0000 |
| Sex | 0.8939 | 0.0304 | 29.4047 | 0.0000 |
| Age at recruitment | 0.0854 | 0.0023 | 36.3886 | 0.0000 |
| Number of days/week walked 10+ minutes | 0.0498 | 0.0164 | 3.0350 | 0.0024 |
| Number of days/week of moderate physical activity 10+ minutes | 0.0387 | 0.0158 | 2.4446 | 0.0145 |
| Number of days/week of vigorous physical activity 10+ minutes | 0.0553 | 0.0163 | 3.3972 | 0.0007 |
| Usual walking pace | -0.2342 | 0.0247 | -9.4919 | 0.0000 |
| Frequency of friend/family visits | -0.0221 | 0.0123 | -1.7962 | 0.0725 |
| Time spent using computer | 0.0237 | 0.0097 | 2.4591 | 0.0139 |
| Weekly usage of mobile phone in last 3 months | 0.0276 | 0.0123 | 2.2470 | 0.0246 |
| Getting up in morning | 0.0438 | 0.0191 | 2.2984 | 0.0215 |
| Nap during day | 0.0706 | 0.0239 | 2.9534 | 0.0031 |
| Sleeplessness / insomnia | 0.0443 | 0.0196 | 2.2609 | 0.0238 |
| Daytime dozing / sleeping (narcolepsy) | 0.0804 | 0.0273 | 2.9457 | 0.0032 |
| Major dietary changes in the last 5 years | 0.0478 | 0.0160 | 2.9946 | 0.0027 |
| Overall health rating | 0.5566 | 0.0213 | 26.0829 | 0.0000 |
| Leisure/social activities | -0.0305 | 0.0125 | -2.4482 | 0.0144 |
| Types of physical activity in last 4 weeks | -0.0118 | 0.0065 | -1.8328 | 0.0668 |
| Smoking status | 0.1201 | 0.0210 | 5.7114 | 0.0000 |
| Alcohol drinker status | -0.1258 | 0.0308 | -4.0828 | 0.0000 |
| Summed days activity | -0.0441 | 0.0141 | -3.1293 | 0.0018 |
| Townsend deprivation index at recruitment | 0.0117 | 0.0054 | 2.1716 | 0.0299 |
| Own or rent accommodation lived in | 0.0610 | 0.0180 | 3.3817 | 0.0007 |
| Number of vehicles in household | 0.0370 | 0.0186 | 1.9832 | 0.0473 |
| Average total household income before tax | -0.0587 | 0.0148 | -3.9647 | 0.0001 |
| Age completed full time education | -0.0146 | 0.0073 | -1.9951 | 0.0460 |
| Qualifications (primary) | 0.0330 | 0.0097 | 3.4147 | 0.0006 |
| Heating type(s) in home | -0.0847 | 0.0270 | -3.1317 | 0.0017 |
| Ethnic background | 0.0001 | 0.0000 | 2.6498 | 0.0081 |

Overall model performance: $R^2_{McFadden}$: 0.145. $R^2_{Nagelkerke}$: 0.189. AIC: 121622.53. Chi-cuadrado global: 20665.02, df: 41, p = 0.
